# Differential effects of Nordic and Vegetarian diets on lipid metabolism, gut microbiome and cardiometabolic risk factors: A multi-omic perspective from a randomized clinical intervention trial

**DOI:** 10.1101/2025.11.03.25337885

**Authors:** Hanna Huber, Alina Schieren, Anna Donkers, Aakash Mantri, Waldemar Seel, Birgit Stoffel-Wagner, Martin Coenen, Leonie Weinhold, Matthias Schmid, Peter Krawitz, Bolette Hartmann, Jens J. Holst, Jacqueline Leidner, Tal Pacht, Lorenzo Bonaguro, Mohamed Yaghmour, Christoph Thiele, Markus M. Nöthen, Peter Stehle, Marie-Christine Simon

**Affiliations:** Nutrition and Microbiota, Institute of Nutrition and Food Science, University of Bonn; 53115 Bonn, Germany; Translational Dementia Research, German Center of Neurodegenerative Diseases (DZNE), 53127 Bonn, Germany; Institute for Genomic Statistics and Bioinformatics, University of Bonn, 53127 Bonn, Germany; Central Laboratory, Institute of Clinical Chemistry and Clinical Pharmacology, University Hospital Bonn, 53127 Bonn, Germany; Clinical Study Core Unit, Study Center Bonn, Institute of Clinical Chemistry and Clinical Pharmacology, University Hospital Bonn, 53127 Bonn, Germany; Institute of Medical Biometry, Informatics and Epidemiology, University Hospital Bonn, 53127 Bonn, Germany; Biomedical Sciences and Novo Nordisk Foundation Center for Basic Metabolic Research, University of Copenhagen, 2200 København, Denmark; Systems Medicine, German Center for Neurodegenerative Diseases (DZNE), 53127 Bonn, Germany; Genomics and Immunoregulation, LIMES Life and Medical Science Institute, University of Bonn, 53115 Bonn, Germany; LIMES Life and Medical Science Institute, University of Bonn, 53115 Bonn, Germany; Institute of Human Genetics, University of Bonn & University Hospital Bonn, 53127 Bonn, Germany; Nutritional Physiology, Institute of Nutrition and Food Science, University of Bonn; 53115 Bonn, Germany

**Keywords:** lipid metabolism, glucagon-like peptide 1, gut-brain axis, obesity, dietary intervention, vegetarian diet, Nordic diet, gut microbiome, genetic predisposition, cardiometabolic risk factors, personalized nutrition, multi-omics, peripheral immune composition, high-risk population

## Abstract

**Background:** Beneficial effects of diets with predominance of plant-based foods as fruits, vegetables, whole grain and plant-protein and less animal-based foods, or so-called “*plant-based*” diets, on cardiometabolic risk have been reported. We aimed to examine the effects of two distinct plant-based diets on intermediate cardiometabolic risk factors, particularly lipid metabolism, while also considering the impact of the gut microbiome, genetic predisposition, and immune status on the metabolic response to a dietary change.

**Methods:** In this randomized, controlled dietary intervention trial, 120 obese adults (59 ± 1 years, 70 females) consumed an isoenergetic Nordic (ND) or a lacto-ovo vegetarian diet (VD) or maintained their habitual diet (control group) for six weeks. At baseline and after the end of the trial, in-depth metabolic characterization was conducted, including measurement of incretins such as glucagon-like peptide 1 (GLP-1), postprandial lipids with lipidomic profiling, and microbiome analysis. Genetic makeup and peripheral immune system composition were characterized at baseline.

**Results:** ND intervention beneficially altered lipid metabolism up to 15%. The largest changes were observed in participants with high genetic predisposition for hyperlipidemia, while lipid metabolism remained stable upon VD. The changes observed were associated with specific microbial signatures and pathways. GLP-1 levels remained stable during the study period.

**Conclusion:** The metabolic response to a dietary change in obese adults is linked to the individual genetic risk, baseline microbiome composition, and immune phenotype, pointing towards a personalized nutritional approach in preventing cardiometabolic diseases.

## INTRODUCTION

The prevalence of non-communicable diseases, particularly cardiometabolic diseases such as cardiovascular diseases (CVD), stroke and diabetes mellitus, and their major risk factors is increasing worldwide and have been framed as global emergency in both developed and developing countries, especially due to their long duration and slow progression^1,2^. One major risk factor for this development is the adherence to a ‘Western-type’ dietary pattern: the consumption of high caloric and energy dense food easily leads to hyperenergetic nutrition associated with long-term metabolic alterations. Furthermore, a concurrent decline in energy expenditure due to an increasingly sedentary lifestyle amplifies the positive energy balance, resulting in weight gain and impaired blood lipid profile, which has been associated with a range of cardiometabolic diseases^3,4^. Fasting hyperlipidemia, a hallmark of metabolic syndrome and CVD, is characterized by both elevated plasma triglyceride-rich lipoproteins and non-high-density lipoprotein (HDL)-cholesterol and a reduction in HDL cholesterol concentrations^5–7^. Postprandial lipemia, particularly postprandial hypertriglyceridemia, has also emerged as an important cardiovascular risk factor and is considered an early marker of atherosclerosis^8–11^. In recent years, it has additionally been discovered that a Western-type dietary pattern might induce alterations in the fecal microbial composition^12^, a readily accessible indicator of the gut microbiome, which in turn also has been linked to various cardiometabolic diseases such as diabetes mellitus, obesity, and CVD^13–15^. Moreover, the adaptive immune system is associated with atherosclerosis and CVD risk due to a sustained and chronic inflammatory state^16,17^. Many epidemiological studies suggested that adherence to a (lifelong) plant-based diet with high amounts of fibers may contribute to minimizing the incidence of metabolic syndrome traits such as visceral fat distribution, (pre)hypertension, dyslipidemia, and/or elevated C-reactive protein (CRP) levels, unfavorable alterations in gastrointestinal hormone secretion as glucagon-like peptide 1 (GLP-1) and dysregulation of the lipidome, decreasing the overall risk of developing cardiometabolic diseases^18–23^. According to the World Health Organization (WHO), plant-based diets constitute a diverse range of dietary patterns that emphasize foods derived from plant sources coupled with lower consumption or exclusion of animal products. So called Vegetarian diets (VD) form a subset of plant-based diets, which may exclude the consumption of some or all forms of animal foods^24^. Thus, observational studies indicate, that adherence to a lacto-ovo VD (with plant-derived foods, milk[products] and eggs) or a so called ‘Nordic’ diet (ND, with typical food items locally available in the Scandinavian area, such as berries, root vegetables and cabbage, wholegrain oats and rye, boiled potatoes and fatty sea fish) was associated with favorable blood lipid profile, glucose control, and body composition^25–30^ as well as lower overall mortality^31,32^. Moreover, growing evidence shows that dietary and lifestyle interventions, including VD and ND interventions, have the potential to be broadly efficacious strategies to improve metabolic regulation and treat and/or delay cardiometabolic diseases and thus, reduce health care costs^33,34^.

Since comparative intervention studies contrasting differently composed plant-based diets in the same population are lacking, a ‘ranking’ of food patterns with respect to quality and quantity of desirable metabolic effects cannot be established. Thus, it remains unclear which plant-based diet (Nordic, lacto-ovo vegetarian or even vegan) might be the most promising to beneficially influence biochemical and clinical variables within a relatively short time interval. Furthermore, the personalized response upon a diet is different, thus an in-depth assessment of individual factors influencing cardiometabolic health, such as the lipidome, genetics, the peripheral immune system, and the gut microbiome, will provide further insights into the mechanisms of dietary interventions for the prevention and management of cardiometabolic diseases.

This randomized, controlled dietary intervention trial investigates the metabolic response to two different plant-based dietary patterns—both high in fruits, vegetables, unsaturated fatty acids, and dietary fiber, and low in meat, sweets, sugar-sweetened beverages, and fast food— in individuals with a risk phenotype for cardiometabolic diseases. The specific aim of this 6-week parallel-arm intervention trial in 120 adults is to assess the effects of a ND and a VD intervention vs. a habitual Western-type control diet (HD) on fasting and meal-stimulated lipid metabolism and gastrointestinal hormone secretion, with the ultimate goal of enhancing cardiometabolic health. Moreover, the impact of the gut microbiome composition, the genetic predisposition, and the immunological signature were considered. This multi-layer, integrative strategy enhances the sensitivity to detect minor but biologically relevant changes and provides mechanistic insights into individual variability in response to dietary interventions regarding emerging evidence of personalized nutritional strategies.

## RESULTS

### Participants

A total of 151 participants were screened; 120 participants (50 men and 70 women) met the inclusion criteria, presenting at least one metabolic syndrome trait (including visceral fat distribution, (pre)hypertension, dyslipidemia, and a proinflammatory state) and were enrolled in the study between November 2018 and April 2020. Participants in the ND and VD study groups followed menu plans comprising food items characteristic of the respective dietary pattern (**Supplementary Table S1**) and matching their individual energy needs (total energy expenditure, TEE). Participants in the control group maintained their habitual diet (HD) and physical activity throughout the trial. A total of 112 participants completed the six weeks dietary intervention study, and their respective per-protocol data were included in the analysis (**Supplementary Figure S1**). Dropouts (7.5%; HD, n = 1 [2.5%]; ND, n = 3 [7.3%]; VD, n = 5 [12.5%], respectively) occurred due to personal reasons (n = 2, 1.7%), health reasons (n = 2, 2.5%) and the coronavirus disease 2019 (COVID-19) pandemic (due to COVID-19 infection or quarantine regulations; n = 4, 3.3%). The baseline characteristics of the participants at study entry are listed in **Table 1**. All participants were overweight (53%) or obese (47%), had a visceral fat distribution indicated by an elevated waist circumference (WC; men > 94cm and women > 80cm), and at least one further metabolic syndrome trait. On group level, the participants had elevated total-cholesterol (> 5.2 mmol/l) and LDL-cholesterol (> 3.4 mmol/l) levels, hypertension stage 1 and a proinflammatory state (hsCRP > 3.0 mg/l). Participants were characterized by an elevated calculated 10-year CVD risk (Framingham Risk Score (FRS) > 10%)^35^. The daily energy intake before study recruitment was distributed as covered by 42.6 ± 0.5 E% fat (16.5 ± 0.2 E% saturated fatty acids) and 37.4 ± 0.5 E% carbohydrates (6.0 ± 0.2 E% monosaccharides and 11.3 ± 0.3 E% disaccharides, 21.0 g/d / 1.7 ± 0.0 E% dietary fiber) reflecting a typical Western-type dietary behavior (data from food frequency questionnaire (FFQ), **Supplementary Table S2**).

**Table 1:** Baseline characteristics^a^.

|  | Total (n = 120) | HD (n = 39) | ND (n = 41) | VD (n = 40) |
| --- | --- | --- | --- | --- |
| Sex, m/f | 50 / 70 | 20 / 19 | 15 / 27 | 15 / 24 |
| Age, y | 59.7 ± 0.7 | 59.3 ± 1.2 | 59.9 ± 1.1 | 60.0 ± 1.2 |
| BMI, kg/m<sup>2</sup> | 31.1 ± 0.3 | 30.9 ± 0.5 | 31.0 ± 0.5 | 31.5 ± 0.6 |
| Waist circumference, cm | 106.2 ± 0.9 | 105.8 ± 1.7 | 105.0 ± 1.4 | 108.0 ± 1.5 |
| Hip circumference, cm | 111.2 ± 0.9 | 109.9 ± 1.5 | 110.4 ± 1.5 | 113.5 ± 1.6 |
| Waist-to-height-ratio | 0.6 ± 0.00 | 0.6 ± 0.0 | 0.6 ± 0.0 | 0.6 ± 0.0 |
| BP systolic, mmHg | 137.4 ± 1.6 | 136.1 ± 2.6 | 136.4 ± 2.5 | 139.8 ± 3.0 |
| BP diastolic, mmHg | 87.9 ± 1.0 | 86.6 ± 1.6 | 88.3 ± 1.7 | 88.8 ± 1.8 |
| Heart rate, min<sup>-1</sup> | 68.4 ± 1.0 | 68.6 ± 1.9 | 69.3 ± 1.6 | 67.2 ± 1.7 |
| REE, kcal/d | 1720.1 ± 27.0 | 1811.7 ± 47.3 | 1722.2 ± 44.6 | 1628.6 ± 45.3 |
| Body fat, % | 41.4 ± 0.7 | 38.9 ± 1.2 | 41.2 ± 1.2 | 44.1 ± 1.3 |
| Creatinine, µmol/l | 70.8 ± 1.5 | 69.4 ± 2.1 | 71.1 ± 2.8 | 71.8 ± 2.6 |
| Urea, mmol/l | 5.0 ± 0.1 | 5.0 ± 0.2 | 5.3 ± 0.2 | 4.8 ± 0.2 |
| Bilirubin, µmol/l | 9.5 ± 0.4 | 8.9 ± 0.7 | 9.6 ± 0.6 | 9.9 ± 0.7 |
| Uric acid, µmol/l | 321.8 ± 5.9 | 324.4 ± 10.5 | 323.8 ± 9.8 | 317.1 ± 10.5 |
| GGT, U/l | 31.8 ± 2.1 | 28.4 ± 1.8 | 33.5 ± 3.7 | 33.5 ± 4.7 |
| ALT, U/l | 29.0 ± 1.1 | 28.3 ± 1.8 | 28.7 ± 1.7 | 30.1 ± 2.1 |
| AST, U/l | 27.3 ± 0.7 | 26.5 ± 1.2 | 28.1 ± 1.5 | 27.2 ± 1.2 |
| Serum triglycerides, mmol/l | 1.5 ± 0.1 | 1.4 ± 0.1 | 1.6 ± 0.1 | 1.6 ± 0.1 |
| Serum total cholesterol, mmol/l | 5.5 ± 0.1 | 5.1 ± 0.2 | 5.9 ± 0.2 | 5.6 ± 0.2 |
| Serum HDL cholesterol, mmol/l | 1.5 ± 0.0 | 1.50 ± 0.1 | 1.5 ± 0.1 | 1.5 ± 0.1 |
| Serum LDL cholesterol, mmol/l | 3.8 ± 0.1 | 3.5 ± 0.2 | 4.1 ± 0.2 | 3.9 ± 0.1 |
| Plasma glucose, mmol/l | 5.2 ± 0.1 | 5.2 ± 0.1 | 5.2 ± 0.1 | 5.3 ± 0.1 |
| HOMA | 3.1 ± 0.2 | 3.2 ± 0.4 | 2.9 ± 0.2 | 3.1 ± 0.3 |
| HbA1c, % | 5.4 ± 0.0 | 5.5 ± 0.1 | 5.4 ± 0.1 | 5.4 ± 0.1 |
| Serum hsCRP, mg/l | 3.3 ± 0.3 | 3.4 ± 0.5 | 2.9 ± 0.4 | 3.5 ± 0.8 |
| Serum insulin, pmol/l | 92.7 ± 4.6 | 95.5 ± 10.4 | 89.6 ± 6.8 | 93.1 ± 6.7 |
| Plasma GLP-1, pmol/l | 4.4 ± 0.2 | 4.2 ± 0.4 | 4.8 ± 0.3 | 4.2 ± 0.4 |
| MSSS | -3.24 ± 0.04 | -3.34 ± 0.07 | -3.24 ± 0.62 | -3.16 ± 0.06 |
| CMRI score | 0.05 ± 0.29 | -0.74 ± 0.53 | -0.14 ± 0.40 | 1.02 ± 0.54 |
| FHS risk score | 12.4 ± 0.4 | 12.2 ± 0.6 | 12.6 ± 0.6 | 12.2 ± 0.7 |

<sup>a</sup> Data are shown as mean ± SEM; n = 120
ALT, Alanine aminotransferase; AST, Aspartate aminotransferase; BMI, Body mass index; BP, Blood pressure; CMRI, Cardiometabolic risk; f, Female; FHS, Framingham heart study; GGT, Gamma glutamyl transferase; GLP-1; Glucagon-like peptide 1; HD, habitual diet group; HDL, High-density lipoprotein; HOMA, Homeostasis model assessment; hsCRP, High-sensitivity C-reactive protein; LDL, Low-density lipoprotein; m, Male; MSSS, Metabolic syndrome severity score; ND, Nordic diet group; REE, Resting energy expenditure; VD, Vegetarian diet group.

### Validated compliance markers confirmed good adherence to the study diets

To assess the compliance with the study diets, a short version of the FFQ and several blood biomarkers specifically related to the consumption of key components of the specific diets (**Supplementary Table S1**) and anthropometric measures at baseline and after six weeks have been monitored.

The short-form FFQ revealed intervention diet-specific changes in the consumption frequency of key food groups at the end of the study period. Briefly, in the ND group, an increase in ND-specific key foods was observed, e.g., mean change ± SD; rapeseed oil +0.7 ± 0.1 portions/d, p-value from linear mixed models compared to baseline and HD, p < 0.001; berries +0.6 ± 0.1 portions/d, p < 0.001; nuts +1.1 ± 0.5 portions/d, p < 0.05; fish +1.2 ± 0.5 portions/d, p = 0.001. In the VD group, it became evident that meat (products) and fish had been eliminated from the diet at endline, e.g., mean change ± SD; pork, beef or lamb -0.4 ± 0.0 portions/d, p-value from linear mixed models compared to baseline and HD, p = 0.232; poultry and chicken -0.3 ± 0.0 portions/d, p < 0.001; cold cuts and processed meat -0.4 ± 0.1 portions/d, p < 0.001; **Supplementary Table S3**). Moreover, consumption frequencies of food items specific to our VD intervention were increased at the end of the study compared to baseline, e.g., mean change ± SD; olive oil +0.7 ± 0.2 portions/d, p-value from linear mixed models compared to baseline and HD, diet x visit interaction, p < 0.001; whole grain wheat +1.8 ± 0.7 portions/d, p = 0.004. In both the ND and VD group, the consumption of foods associated with a Western-style diet was decreased after 6 weeks, highlighting a shift to a healthier diet; e.g., mean change ± SD; cold cuts and processed meat -0.8 ± 0.4 portions/d in the ND group, -0.4 ± 0.1 portions/d in the VD group, p-value from linear mixed models compared to baseline and HD, p < 0.001; sweets -0.9 ± 0.5 portions/d in the ND group; -0.7 ± 0.5 portions/d in the VD group, p = 0.503; sugar sweetened beverages -0.1 ± 0.0 portions/d in the ND group; -0.1 ± 0.0 portions/d in the VD group, p = 0.646, however, this did not reach statistical significance. In the HD groups, no to small changes in consumption frequencies from baseline to the end of the study were observed (**Supplementary Table S3**).

Next, we evaluated the adherence to the study diets using food or food-group specific blood and anthropometric biomarkers. Compared to HD, after six weeks, participants in the ND group had a decline in plasma phospholipid oleic acid (C18:1n9c; β = -77.90 ± 29.84, p = 0.008) suggestive of a decreased olive oil consumption and an increase in plasma phospholipid eicosapentaenoic acid (EPA, C20:5n3c; β = 33.60 ± 6.56, p < 0.001) and docosahexaenoic acid (DHA, C22:6n3c; β = 49.70 ± 8.78, p < 0.001), reflecting an increased fish consumption. Participants in the VD group had a slight increase in plasma phospholipid oleic acid (β = 7.66 ± 30.28, p = 0.008), and a decrease in EPA (β = -6.56 ± 7.96, p < 0.001) and DHA (C22:6n3c; β = -10.41 ± 8.93, p < 0.001), reflecting an increased oleic acid and a decreased fish consumption. The concentrations of plasma phospholipid myristic acid (C14:0) and α-linoleic acid (C18:3n3c), surrogate markers of dairy lipids, showed no group differences. α-Tocopherol decreased after both diet interventions (ND: β = -1791.26 ± 586.39, VD: β = -56.85 ± 601.40, p = 0.003). No significant group-differences in plasma concentrations of retinol, β-carotene and vitamin C, reflective of vegetable and fruits, were found at the end of the trial (all p > 0.05, p-value from linear mixed models compared to baseline and HD; **Table 2**). Furthermore, consistent with the isoenergetic study design, there were no significant changes in body weight in any group before and after the intervention. Moreover, no significant changes in body composition and waist and hip circumference occurred in any group (all p > 0.05, p-value from linear mixed models compared to baseline and HD; **Supplementary Table S4**). Overall, the objective evaluation—alterations in fatty acid composition of the plasma phospholipids linked to the consumption of diet-specific food items combined with body weight stability and altered dietary behavior as shown by the short-form FFQ—confirmed good participant compliance throughout the study period.

**Table 2:**
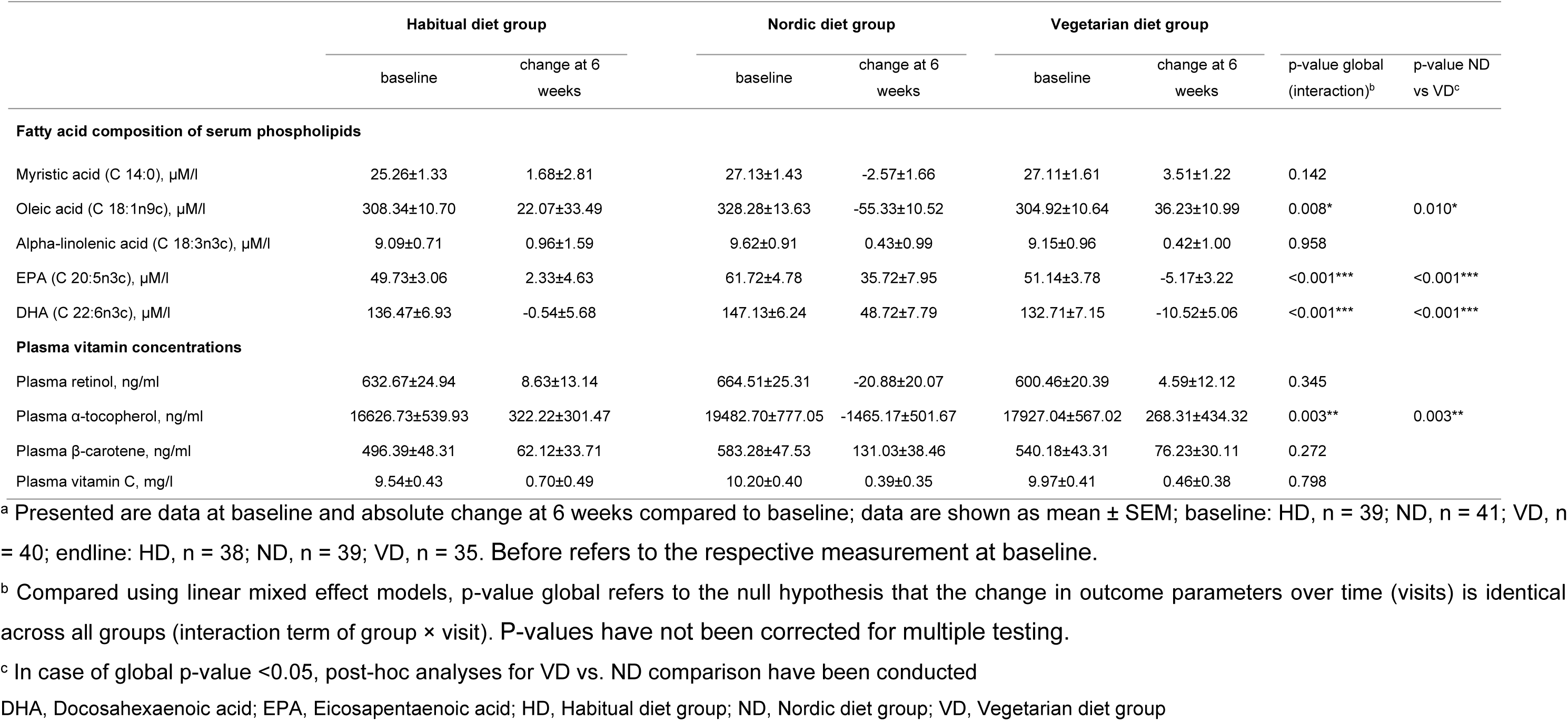
Compliance biomarkers before and after the dietary intervention^a^.

### Nordic Diet beneficially altered lipid metabolism

To examine the effect of the ND and VD on lipid metabolism, we measured fasting and meal-stimulated lipid profile during MMTT before and after the 6-weeks dietary intervention.

Remarkably, ND intervention lead to strong reductions in total cholesterol (ND: β = -0.40 ± 0.12, VD: β = 0.11 ± 0.12, diet x visit interaction, p = 0.001), LDL cholesterol (ND: β = -0.22 ± 0.04, VD: β = 0.02 ± 0.04, diet x visit interaction, p = 0.007) and fasting triglyceride levels (ND: β = -0.25 ± 0.10, VD: β = 0.20 ± 0.10, diet x visit interaction, p < 0.001), while this effect was not observed after VD intervention. Fasting values of serum HDL cholesterol, serum NEFA, postprandial AUC (diet x visit interaction, all p > 0.05) and time course of serum triglyceride and serum NEFA levels during MMTT remained stable (diet x visit x time interaction, all p > 0.05; **Figure 1, Supplementary Table S5, Supplementary Table S6**).

**Figure 1:**
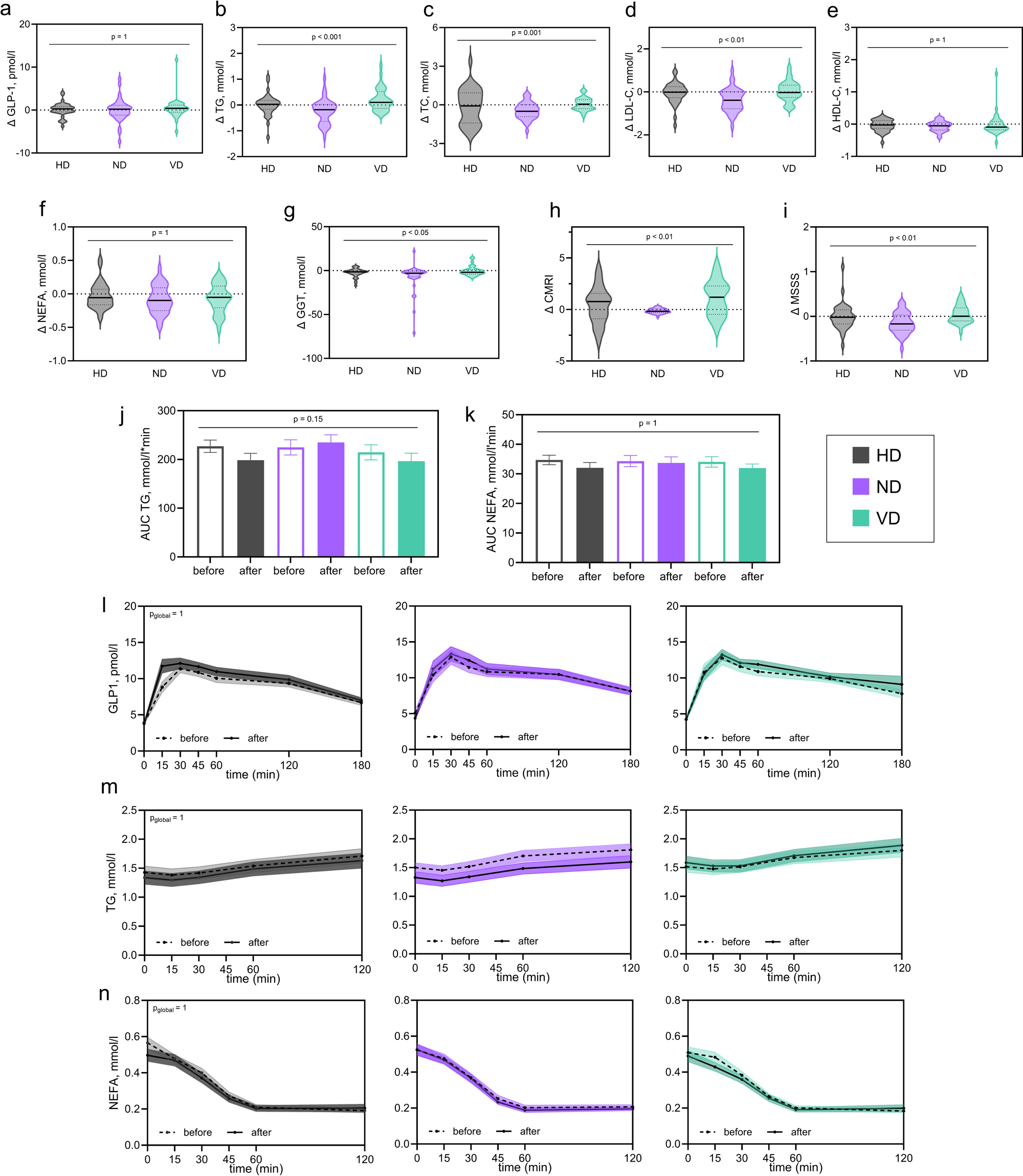
Longitudinal changes in parameters of lipid metabolism, gut hormones, liver enzymes and cardiometabolic risk indices at baseline and after six weeks in participants with metabolic syndrome traits Displayed are the absolute changes in fasting **(a)** plasma GLP-1, **(b)** serum triglycerides, **(c)** plasma total cholesterol, **(d)** plasma LDL cholesterol, **(e)** plasma HDL cholesterol, **(f)** fasting serum NEFAs, **(g)** fasting serum GGT and the absolute change in **(h)** CMRI and **(i)** MSSS scores after 6 weeks. In **(j)**, the AUC of serum triglycerides during MMTT, in **(k)**, the AUC of serum NEFAs during MMTT, in **(l)**, the postprandial time course of plasma GLP-1, in **(m)**, the serum triglycerides and in **(n)**, the serum NEFA concentrations during MMTT at baseline and after six weeks is presented In a-i, violin plots of the absolute change at endline compared to baseline are presented; the violin plot presents the median (solid line) and quartiles (dashed lines). In j-n, data is presented as mean ± SEM (shaded area). N = 120; p-value global refers to the null hypothesis that the change in outcome parameters over time (visits) is identical across all groups. For postprandial time course (l-n), p_global_ represents the visit × diet effect for specific time points. P-values have been corrected for multiple testing by Bonferroni-Holm correction. HD group is plotted in grey, ND group is plotted in purple, VD group is plotted in green. Biomarkers at the beginning of the study are presented in dashed lines, while biomarkers at the end of the study are presented in solid lines AUC, area under the curve; CMRI, global cardiometabolic risk index; GLP-1, gucagon like peptide 1; GGT, γ-glutamyl transferase; HD, habitual diet group; HDL-C, high-density lipoprotein cholesterol; LDL-C, low-density lipoprotein cholesterol; MMTT, mixed meal tolerance test; MSSS, metabolic syndrome severity score; NEFA, non-esterified fatty acids; ND, Nordic diet group; TC, total cholesterol; TG, triglycerides; VD, vegetarian diet group

To examine these effects on lipid metabolism in more detail, we investigated the lipidome before and after intervention. Shotgun lipidomics analysis of baseline and end-of trial plasma samples identified 321 lipid species in total, belonging to 18 lipid classes. After 6 weeks of dietary intervention, the concentrations of 11 lipid classes were changed differently between the three diet groups (all p < 0.05), with most classes showing a decrease after ND. Four classes (lysophosphatidylcholines (LPCs), ether-linked lysophosphatidylcholines (LPC-Os) and ether-linked phosphatidylcholines (PC-Os) and ether-linked phosphatidylethanol-amines (PE-Os)) were reduced after VD, while phosphatidylglycerols (PGs) and phosphatidylethanolamines (PEs) were increased after VD (**Figure 2 a, Supplementary Table S7 a**). The normalized average of double bonds increased for six classes after ND (triglycerides (TAGs), PEs and PE-Os, sphingomyelins (SMs), PGs and phosphatidylcholines (PCs)) while it decreased after VD (all p < 0.009) (**Figure 2 b, Supplementary Table S7 a**). The normalized average chain length increased in the same six classes after ND and decreased in ceramides (Cers) and dihexosylceramides (DiHexCers). After VD, normalized average chain length increased slightly in PE-Os and Cers and decreased in five classes (TAGs, SMs, PGs, PEs, PCs) (all p < 0.05) (**Figure 2 c, Supplementary Table S7 a**). The share of very long TAGs (> 58 C-atoms) of all TAGs increased after ND while remaining stable in HD and VD (p < 0.001) (**Figure 2 d**). For the analysis of changes in lipid species, 240 lipid species from 17 lipid classes were included. Of those, 108 species from 15 lipid classes changed differently between the three diets, including 30 TAG species, 26 PCs and 13 PEs (**Figure 2 e, Supplementary Table S7 b**). TAG species with a higher number of double bonds and chain length mainly increased after ND (all p < 0.02) while there was a decrease of those with shorter chains and fewer double bonds, as well as of most species from PC (except for PC (42:4) and PC (42:6)), PE and other classes. After VD, the opposite effect was observed, apart from a few single species (e.g. PC-O (34:1), PC-O (36:3), CE (22:2), PE-O (38:5) and PE-O (40:8) which decreased after both diets. In the HD group, most species showed little to no change except for an increase in few species like PG (32:1), PG (34:2) and a reduction in e.g. CE (18:4) which remained stable after ND and VD diet (**Supplementary Table S7 b**). These results suggest that ND beneficially affect membrane fluidity, cell signaling, and metabolic processes and lead to a reduction in cardiometabolic risk by improving interims cardiometabolic risk factors, while less beneficial effects where observed after VD.

**Figure 2:**
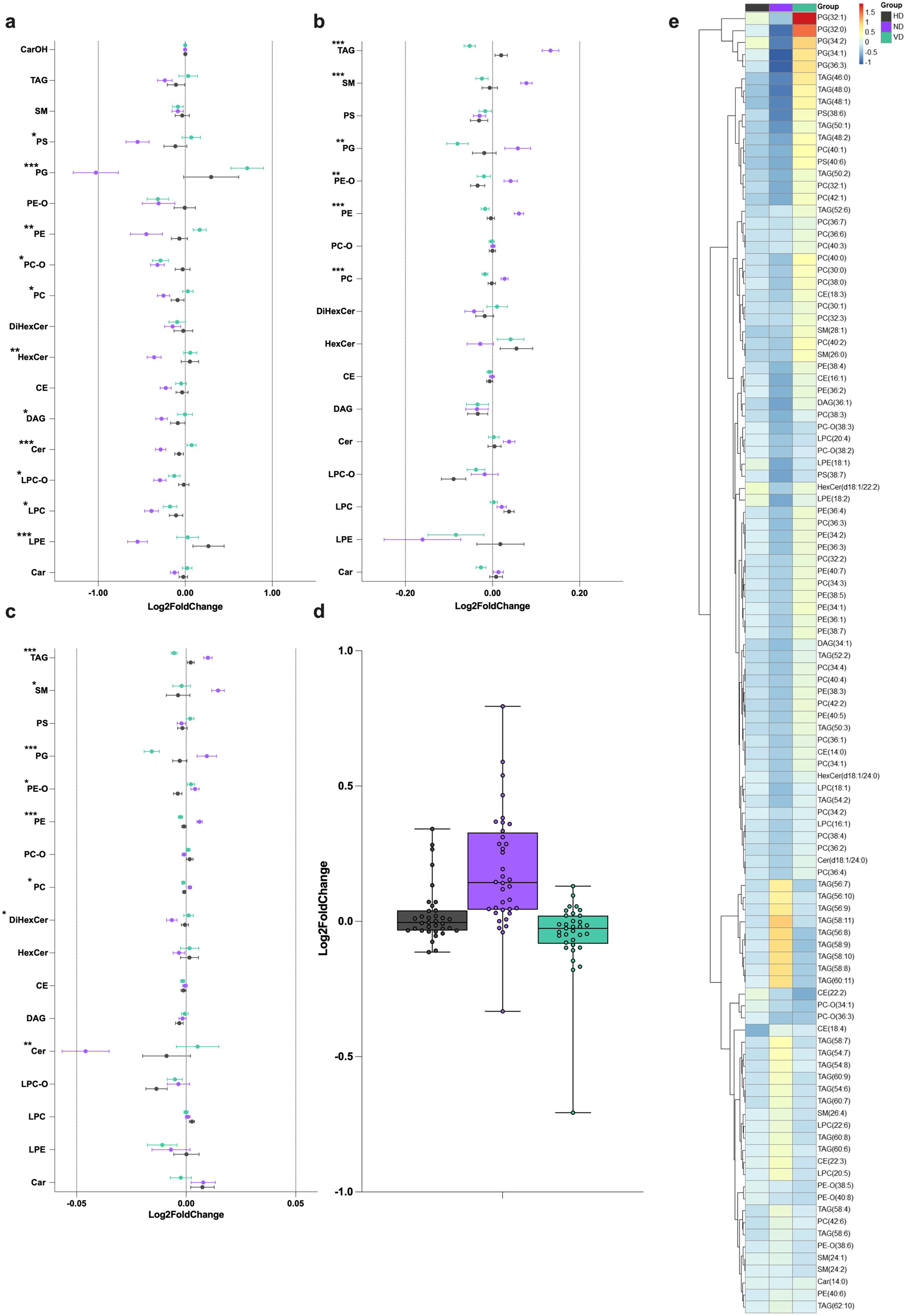
Plasma lipidomic profile before and after dietary intervention in participants with metabolic syndrome traits Displayed are the mean log2 fold-changes with SEM in **(a)** 18 lipid classes, **(b)** normalized average number of double bonds per lipid class for 17^1^ lipid classes, **(c)** normalized acyl chain length per lipid class for 17^1^ lipid classes, (**d**) the share of TAG species with more than 58 C-atoms of all TAGs and **(e)** all significant changes of lipid species among intervention groups; colors represent direction of change; blue: decrease; red: increase. HD group is displayed in grey, ND group is displayed in purple, VD group is displayed in green Group differences in biomarker changes at the end of the study compared to baseline were assessed by linear (mixed-effects) models or zero-inflated gaussian models for variables with zero-inflation. Significance is indicated as follows: *p < 0.05, **p < 0.01, ***p < 0.001 after FDR correction ^1^for the CarOH class, the normalized average number of double bonds and acyl chain length could not be calculated as the concentration was ∼0 pmol/2µl Car, acylcarnitines; CE, cholesteryl esters; Cer, ceramides; DAG, diacylglycerols; DiHexCer, dihexosylceramides; FDR, false discovery rate; HD, habitual diet group; HexCer, hexosylceramides; LPC, lysophosphatidylcholines; LPC-O, ether-linked lysophosphatidylcholines; LPE, lysophosphatidylethanolamines; ND, Nordic diet group; PC, phosphatidylcholines; PC-O, ether-linked phosphatidylcholines; PE, phosphatidylethanolamines; PE-O, ether-linked phosphatidylethanolamine; PG, phosphatidylglycerols; PS, phosphatidylserines; SEM, standard error of the mean; SM, sphingomyelins; TAG, triacylglycerols; VD, vegetarian diet group

### Nordic Diet improved liver enzyme levels

Serum GGT concentrations decreased by 15 % after ND intervention (β = -4.58 ± 2.20, diet x visit interaction, p = 0.048, **Figure 1, Supplementary Table S6**), and remained stable upon VD. Also, ALT and AST remained stable in ND or VD group (both p > 0.05). Consistent with the results on lipid metabolism, the ND resulted in beneficial change in liver enzyme levels, indicating improved liver function and reduced oxidative stress, while VD stabilized liver enzymes.

### Improved cardiometabolic risk scores in the ND group

To investigate the effect of a ND and VD intervention on the severity of metabolic syndrome traits and interims cardiometabolic risk, corresponding risk scores were calculated at baseline and at the end of the trial. After 6 weeks, the Metabolic Syndrome Severity Score (MSSS) and Cardiometabolic Risk Index (CMRI) were significantly improved in the ND group, but showed opposite effects upon VD (MSSS: ND: β=-0.118, VD: β=0.072, diet x visit interaction, p = 0.001 CMRI: ND: β=-0.798, VD: β=0.710, diet x visit interaction, p = 0.002, **Figure 1, Supplementary Table S6**).

Furthermore, diet-related modifications in the lipidomic profile were associated with improvements in cardiometabolic risk scores. We found significant correlations for the adjusted change in 61 lipidomic variables with the change in the MSSS and CMRI scores. Especially lipids that were decreased by ND, exhibited a positive correlation with the scores. This included TAGs with a shorter chain length and fewer double bonds, such as TAG(46:0), TAG(48:0), TAG(48:1), and PC with few double bonds, such as PC(40:0), PC(40:1), PC(42:1). Accordingly, total TAG chain length, increased only by ND, showed an inverse correlation with MSSS and CMRI (**Supplementary Table S8**). Adding to our previous findings, these results suggest extended metabolic benefits of the ND intervention modulated by an alteration of the quality and functionality of different lipid species, lowering the risk of chronic metabolic disorders. Thus, the two different health-promoting study diets showed specific effects on metabolism.

### The Nordic and Vegetarian diet induced taxonomic changes in gut microbial signature, while GLP-1 levels remain stable after 6 weeks

To assess the effect of our ND and VD intervention on the secretion of gut hormones, we measured fasting and meal-stimulated GLP-1 levels. After 6-week ND and VD intervention, fasting and postprandial GLP-1 levels during Mixed meal tolerance tests (MMTT) remained stable in all groups (diet × visit × time interaction, p = 1; **Figure 1**, **Supplementary Table S5**).

To investigate the effect of the ND and VD dietary interventions on gut microbial signature, 16S rRNA amplicon sequencing was performed from fecal samples collected at baseline and at the end of the trial. In addition, to analyze the effect of the dietary change at a greater microbiome resolution and identify putative functional genes, whole genome sequencing (WGS) was performed in a random subgroup (n = 34). At baseline, the three most common bacterial genera in the population identified by 16S sequencing were *Blautia*, *Phocaeicola* and *Bacteroides*, possibly reflecting the typical Western-type diet^36^. Both 16S and WGS analyses revealed no significant difference in overall microbiome composition between the three diet groups at baseline [beta diversity; p = 0.77 in 16S sample (generalized UniFrac dissimilarity); p = 0.56 in WGS sample (Aitchison distance)]. Of the viruses classified using WGS and Kraken, over 99% belonged to the phylum *Uroviricota*. Here, two of the most abundant virus genera were *Carjivirus* and *Junduvirus;* bacteriophages which are known to be hosted by various *Bacteroides* spp^37^. This suggests that the abundance of phages is closely related to the abundance of specific bacteria. However, the viral microbiome generally still showed a considerably higher variance between the participants than the bacterial microbiome, which may mask the induced effects of the dietary intervention.

Thus, alpha diversity (Richness and Shannon Index; **Figures 3 a + b; Supplementary Table S9**) and beta diversity (generalized UniFrac distances; **Figures 3 c + d**) in the 16S samples remained stable in all groups (all p > 0.05), which was also confirmed by the WGS data which was available from a random subgroup (n = 34; diet × visit interaction; Richness: p = 0.99, and Shannon index: p = 0.92; **Supplementary Table S9**). Additionally, WGS analysis showed an overall diverse but stable microbiome composition independent of the three diets at the end of the trial (p = 0.94). Furthermore, differential abundance analysis revealed stable bacteria and virus species or MetaCyc pathways after the intervention (all p > 0.05 after FDR correction; **Supplementary File S1**). For the WGS subsample, the baseline characteristics are shown in **Supplementary Table S10,** the anthropometric and metabolic characteristics at study entry and after six weeks are reported in **Supplementary Table S11**. In line, fecal short chain fatty acids (SCFA), measured to provide further insights into the functional activity of the gut microbiome, remained stable after ND and VD compared to HD group (**Supplementary Table S12**).

**Figure 3:**
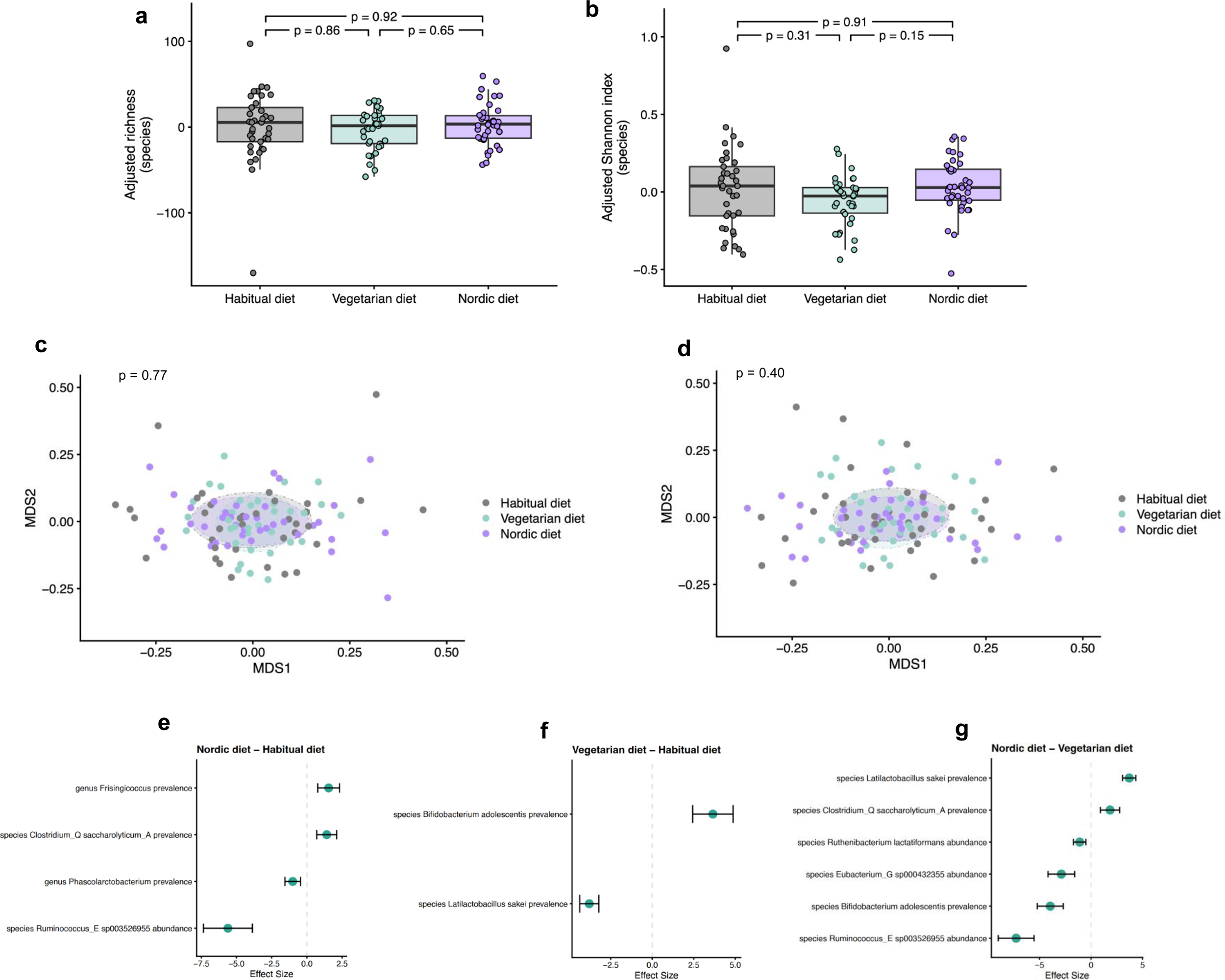
Changes in bacterial microbiome composition and taxonomy before and after dietary intervention in participants with metabolic syndrome traits Shown are the changes in **(a, b)** alpha diversity, **(c, d)** beta diversity, and **(e, f, g)** taxonomic profile after VD and ND dietary intervention and in the HD group **(a)** Change in species richness from baseline to end-of-trial in HD, VD and ND intervention. Shown are the residuals of the adjusted richness values for the three diet groups before adjustment of the diet group variable **(b)** Change in species Shannon index from baseline to end-of-trial in HD, VD and ND intervention. Shown are the residuals of the adjusted Shannon index values for the three diet groups before adjustment of the diet group variable **(c)** Generalized UniFrac dissimilarities of the baseline samples represented in two dimensions by non-metric multidimensional scaling. Each point represents one sample. P-values from permutational ANOVA test **(d)** Generalized UniFrac dissimilarities of the end-of-trial samples represented in two dimensions by non-metric multidimensional scaling. Each point represents one sample. P-values from permutational ANOVA test **(e-g)** Effect sizes of the diet group variable for the taxa whose change in prevalence or abundance from baseline to end-of-trial differed significantly between the respective diet groups. A positive effect size means that prevalence/abundance of the taxon increased from baseline to end-of-trial in the first mentioned group compared to the second mentioned group. Whiskers indicate the 95% confidence intervals of the corresponding effect size ANOVA, analysis of Variance; AUC, area under the curve; FDR, false discovery rate; HD, habitual diet group; MMTT, mixed meal tolerance test; ND, Nordic diet group; VD, vegetarian diet group

After 6 weeks of ND and VD, several significant taxonomic changes compared to HD were observed (**Figures 3 e - g**). Relative abundance/ prevalence at the genus level of individual samples before and after dietary intervention are shown in **Supplementary Figure S2**. After ND intervention, the abundance of *Ruminococcus_E* sp003526955 was lower and the prevalence of *Clostridium_Q saccharolyticum_A* was higher compared to VD and HD group. *Bifidobacterium adolescentis, Eubacterium_G* sp000432355 and *Ruthenibacterium lactatiformans* were less and *Latilactobacillus sakei* was more prevalent/ abundant after ND than after VD intervention (all p < 0.05). At genus level, *Frisingicoccus* was more prevalent in ND and *Phascolarctobacterium* was less prevalent in ND than in HD (all p < 0.05). In the VD group, the prevalence of *Latilactobacillus sakei* and *Bifidobacterium adolescentis* was increased compared to HD group.

Based on the observed beneficial effects of the ND intervention on lipid metabolism, we further investigated the abundance and metabolic impact of specific bacteria that have been associated with lipid metabolism previously^38^. Thus, the *Oscillibacter* genus could be identified in 83.8% of the WGS samples. At species level, five different species were identified, with the *Oscillibacter* species ER4 being the most abundant. In line with recent findings, we found several negative correlations of *Oscillibacter* abundance and triglycerides, total cholesterol levels and multiple lipidomic variables, e.g. Ceramides, Diacylglycerols, and several short-chain TAG (<58 C atoms), and a positive correlation with CER double bounds. Even after multiple testing correction, the negative association with the Cer class and LPC-O(20:5) remained significant for *Oscillibacter* genus (spearman’s rho = 0.59 and -0.57, p = 0.05) and *Oscillibacter* species ER4 (spearman’s rho = -0.61 and -0.60, p = 0.023).

Our results show that the gut microbiome responds distinctively to dietary changes, even in a diverse microbiome at baseline. This may affect the gut microbiome’s structure and function, and host metabolism, especially related to lipid metabolism.

### Diet-induced metabolic changes are linked to genetic risk scores and microbiome signatures

Next, we aimed to identify key molecular drivers and discriminative patterns from our multi-omics data to differentiate the metabolic effects of the ND and VD. We performed a supervised N-integration sparse discriminant analysis using DIABLO^39^ that integrated longitudinal change in metabolic parameters and fecal SCFAs, and baseline genetic risk for hyperlipidemia into a single analysis to identify correlations between ND and VD phenotypes (**Supplementary Table S13, Supplemental File 2**).

Strong interactions of genetic risk score, the lipid metabolism, and the baseline metagenomics clearly distinguished ND and VD groups (balanced error rate (BER) = 0.115, AUC = 0.796, p = 0.028). The final multi-omic signature consisted of 27 bacterial species at baseline, change in levels of fecal lactic and pyruvic acid, three PGS (for total, LDL and HDL cholesterol), nine metabolic parameters (changes of total and LDL cholesterol, triglycerides (fasting and postprandial AUC), NEFA (fasting and postprandial AUC), GGT, ALT and AST) and 22 lipid species (13 TAGs, 5 PCs, 2 CEs, 1 DAG, 1 LPC, **Figure 4 a**). There was a strong positive correlation between baseline abundance of *Sutterella wadsworthensis* with HDL-PGS and changes in ALT, and an inverse correlation with changes in lactic acid and lipid species (e.g., TAG (56:7), TAG (56:8), TAG (54:7) and TAG (54:6)). Changes in total and LDL cholesterol were also negatively correlated with TC- and LDL-PGS (r ≥ -0.62) and changes in lipid species (e.g. TAG (56:7) and TAG (56:8)).

**Figure 4:**
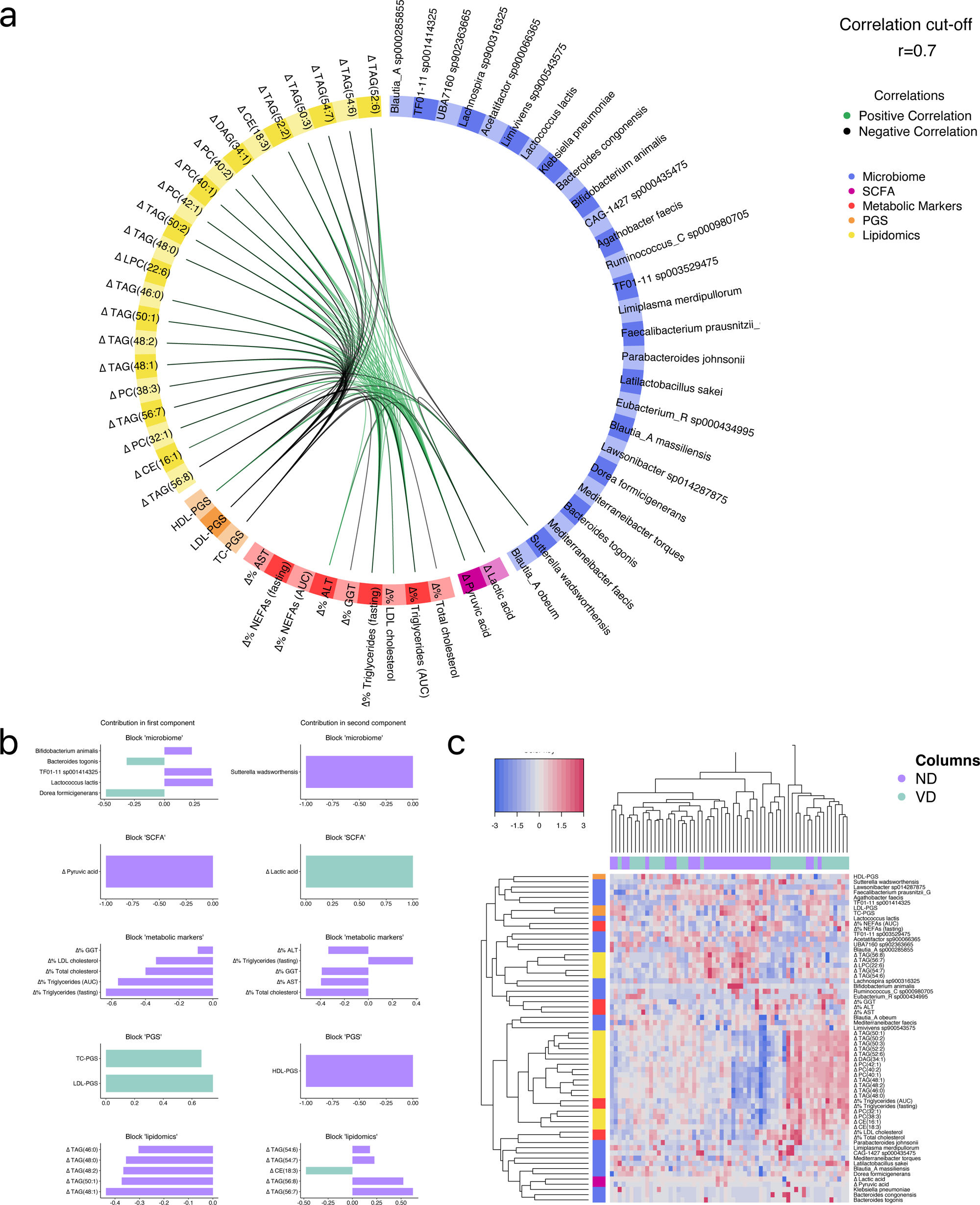
Multi-omics signature of ND vs. VD participants including baseline 16S-derived metagenomics The integrative multivariate analysis identified a multi-omics signature associated with features that discriminate participants in the ND from participants in the HD. **(a)** Circos plot shows correlations (cutoff r > 0.7) between different multi-omic layers: change in metabolic markers (red); baseline PGS (orange), change in lipidomics (yellow), baseline 16S derived metagenomics (dark blue), and change in fecal SCFA (pink) along component one and two, derived from the DIABLO model using sPLS-DA. Intra-block correlations are not presented. Positive correlations are illustrated in green, negative correlations in black. **(b)** The loading plots display the contribution (loading weight) of each feature selected from both components per Omics in an increase of importance from the bottom up. Up to top five contributors are displayed in the loading plot. ND group is displayed in purple, VD group is displayed in green. **(c)** Clustered image maps of component 1 & 2 features. ND group is displayed in purple, VD group is displayed in green. Contribution of change in metabolic markers is shown in red; of PGS in orange, of change in lipidomics in yellow, of 16S derived baseline metagenomics in dark blue, of change in fecal SCFA in pink. Increases in biomarkers are illustrated in red, decreases in blue ALT, alanine aminotransferase; AST, aspartate aminotransferase; AUC, area under the curve; CE, cholesteryl esters; DAG, diacylglycerols; DIABLO, data integration analysis for biomarker discovery using latent components; GGT, γ-glutamyl transferase; HDL, high-density lipoprotein cholesterol; LDL, low-density lipoprotein cholesterol, LPC, lysophosphatidylcholines; ND, Nordic diet group; NEFA, non-esterified fatty acids; PC, phosphatidylcholines; PGS, polygenic risk score; SCFAs, short chain fatty acids; TAG, triacylglycerols; TC; total-cholesterol; TG, _log_triglycerides

LDL-PGS, *Dorea formicigenerans*, and changes in pyruvic acid, were identified as key contributors in discriminating ND and VD group, exhibiting strong correlations with changes in fasting triglycerides and TAGs with shorter chain length and fewer double bonds, e.g., TAG(48:0), TAG(48:1), TAG(50:1). Additional variability between ND and VD groups could be explained by HDL-PGS, *Sutterella wadsworthensis* and changes in lactic acid, total cholesterol and TAG (56:7) (**Figure 4 b**). The features identified by the model were further examined by the clustered image map (**Figure 4 c**) which showed that shorter TAGs showed a higher increase in VD, while longer TAGs increased in ND.

In a further, exploratory approach, we additionally fitted two DIABLO models with WGS data to determine whether changes in microbial pathways (model 2, **Figure 5 a + b**) or baseline abundance of bacterial and viral species at a greater resolution (model 3, **Figure 5 c + d**) distinguish ND and VD and how they may relate to clinical outcomes. In both models, ND and VD groups were well classified (model 2: BER = 0.225, AUC = 0.849, p = 0.019; model 3: BER = 0.225, AUC = 0.854, p = 0.020). Two pathways (LPSSYN-PWY: superpathway of lipopolysaccharide biosynthesis and GLUDEG-I-PWY: GABA shunt), changes in fecal lactic and butyric acid, 37 lipid species (16 TAGs, 5 PCs, 4 LPCs, 4 SMs, 2 DAGs, 2 CEs, 1 PE-O, 1 Cer, 1 HexCer, 1 LPE), six metabolic parameters (changes of total, LDL and HDL cholesterol, triglycerides (postprandial AUC), ALT and AST) and all four PGS were selected. Changes in LPSSYN-PWY were highly negatively correlated with changes in postprandial triglycerides and changes in lipid species (e.g. TAG (48:0), TAG (48:1), TAG (48:2) and TAG (50:1)). Positive correlations for the change in this pathway were evident for the TG-PGS, and changes in lipid species (e.g. TAG (58:8), TAG (58:9) and TAG (58:10)). Changes in GLUDEG-I-PWY showed strong positive correlations with changes in total and LDL cholesterol, changes in ALT and negative associations with LDL-PGS, changes in butyric acid and some lipid species (e.g. SM (24:0), SM (24:1) and SM (24:2)) (**Figure 5 a**), suggesting an involvement in diet-induced alterations in host metabolism.

**Figure 5:**
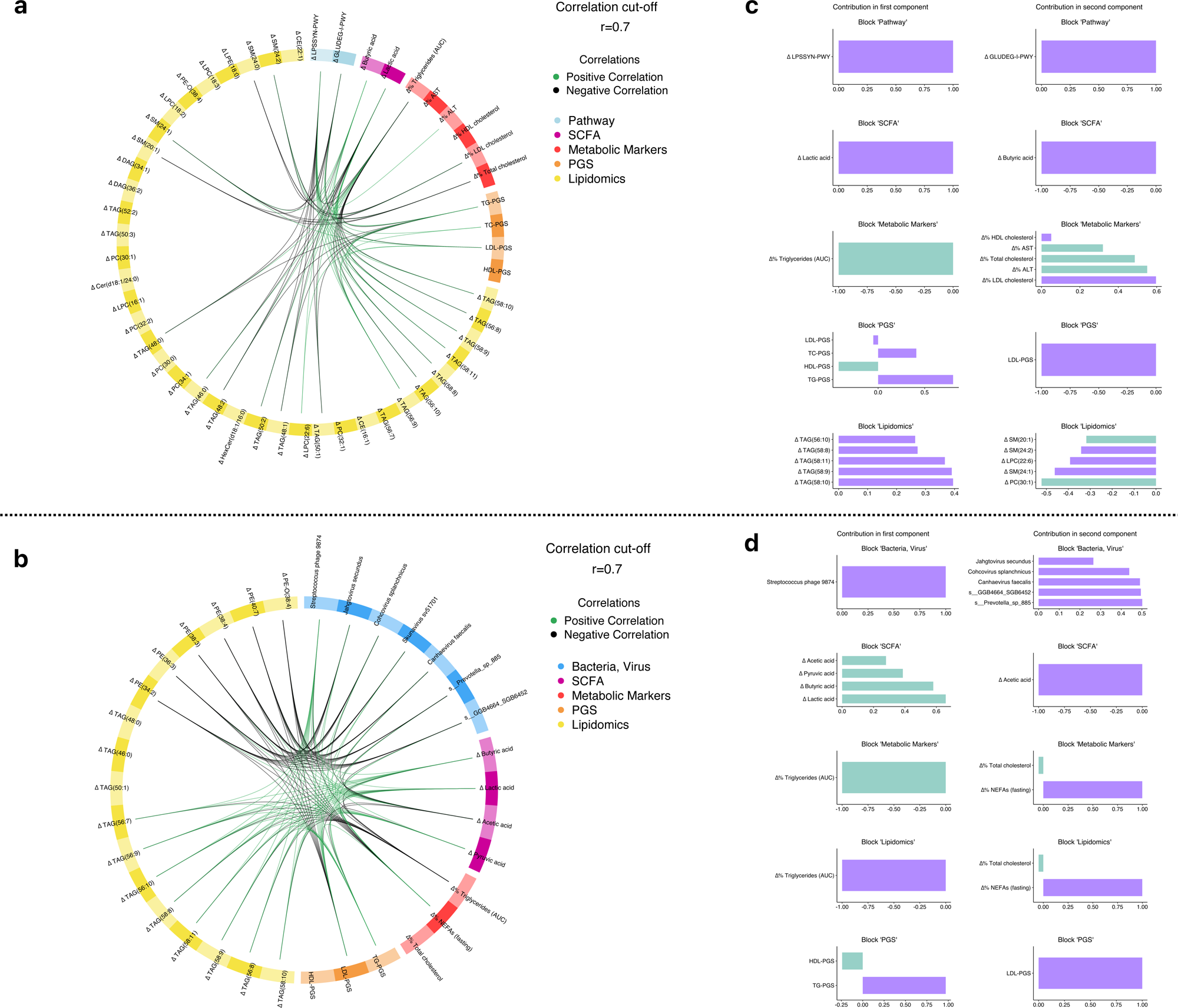
Multi-omics signature of ND vs. VD participants including WGS-derived pathway and microbial data The integrative multivariate analysis identified a multi-omics signature associated with features that discriminate participants in the ND from participants in the VD. Circos plot illustrating the correlations (cutoff r > 0.7) between different multi-omic layers: change in metabolic markers (red); baseline PGS (orange), change in lipidomics (yellow), change in SCFA (pink) and **(a)** change in WGS-derived pathways (light blue) (model 2) and **(b)** baseline WGS-derived microbes and viruses (medium blue) (model 3) along component one and two, derived from the DIABLO model using sPLS-DA. Intra-block correlations are not presented. Positive correlations are illustrated in green, negative correlations in black. The loading plots for **(c)** model 2 and **(d)** model 3 display the contribution (loading weight) of each feature selected from both components per Omics in an increase of importance from the bottom up. Up to top five contributors are displayed in the loading plot. ND group is displayed in purple, VD group is displayed in green. ALT, alanine aminotransferase; AST, aspartate aminotransferase; AUC, area under the curve; CE, cholesteryl esters; Cer, ceramides; DAG, diacylglycerols; DIABLO, data integration analysis for biomarker discovery using latent components; HDL, high-density lipoprotein cholesterol; HexCer, hexosylceramides; LDL, low-density lipoprotein cholesterol, LPC, lysophosphatidylcholines; LPE, lysophosphatidylethanolamines; NEFA, non-esterified fatty acids; PC, phosphatidylcholines; PC-O, ether-linked phosphatidylcholines; PE, phosphatidylethanolamines; PE-O, ether-linked phosphatidylethanolamine; PGS, polygenic risk score; PWY, pathway; SCFAs, short chain fatty acids; SM, sphingomyelins; TAG, triacylglycerols; TC; total-cholesterol; TG, _log_triglycerides

When including the baseline microbiome and viriome in the model (model 3), two bacteria and five viruses, three metabolic parameters (postprandial triglycerides, fasting NEFA and total cholesterol), 17 lipid species (11 TAGs, 5 PEs, 1 PE-O), all four SCFAs and three PGS (LDL-PGS, HDL-PGS, TG-PGS) were selected. *Streptococcus* phage 9874 showed a strong negative correlation with the change in postprandial triglycerides, while being positively associated with changes in all SCFA, changes in lipid species (e.g. TAG (58:8), TAG (58:10), TAG (58:11), TAG (56:8) and TAG (56:9)), and TG-PGS. *Prevotella* sp885, an unclassified species of *Staphylococcaceae* (SGB6452) and *Cohcovirus splanchnicus* correlated inversely with changes in acetic acid, and PE and PE-O lipid species, and positively with changes in fasting NEFA and LDL-PGS (**Figure 5 b**).

All models demonstrated a high discriminative ability (all AUCs ≥ 0.8) and revealed a distinct multi-omic signature consisting of individual bacterial/viral strains present before the dietary intervention, shifts in microbial pathways and metabolites, and polygenic risk scores associated with diet-specific changes in lipid metabolism. These findings suggest a relevant impact of the basal gut microbiome and the genetic predisposition in the individual responsiveness to a dietary change.

### The genetic predisposition for hyperlipidemia has a decisive impact in the response to dietary change

As the polygenic risk scores for hyperlipidemia had a significant contribution in all DIABLO models distinguishing ND and VD, we conducted a detailed analysis of the characteristics and diet-induced metabolic changes within the genetic risk groups.

First, patients with the highest PGS in all 4 PGS categories had the highest baseline mean concentration of total cholesterol, LDL cholesterol and triglycerides compared to individuals with moderate or low genetic risk. Moreover, lipidomics analysis revealed significant differences in lipid classes such as phosphatidylethanolamines (PE), triacylglycerols (TAG), and diacylglycerols (DAG) and specific lipid species in the three groups were found - pairwise post hoc comparisons indicated that the high-risk group had elevated concentrations compared to both the moderate- and low-risk groups (**Supplementary Figure S3**). Next, we investigated the diet-induced changes in lipid metabolism across the PGS groups. As there were significant alterations only after ND intervention, we conducted this analysis in the ND group. The subgroup analyses showed a significant reduction in total and LDL cholesterol levels after ND intervention in participants with the highest LDL-PGS score compared to participants with low and moderate LDL-PGS scores (β = -0.79 ± 0.21, p = 0.014 and β = -0.68 ± 0.15, p = 0.005, respectively). Remarkably, in the high-risk group, total cholesterol levels decreased by 10%, LDL cholesterol levels by 15%. For fasting triglyceride levels, a significant reduction after ND intervention was observed in the moderate LDL-PGS group (β = -0.31 ± 0.10, p = 0.031; **Figure 6**, **Supplementary Table S14)**.

**Figure 6:**
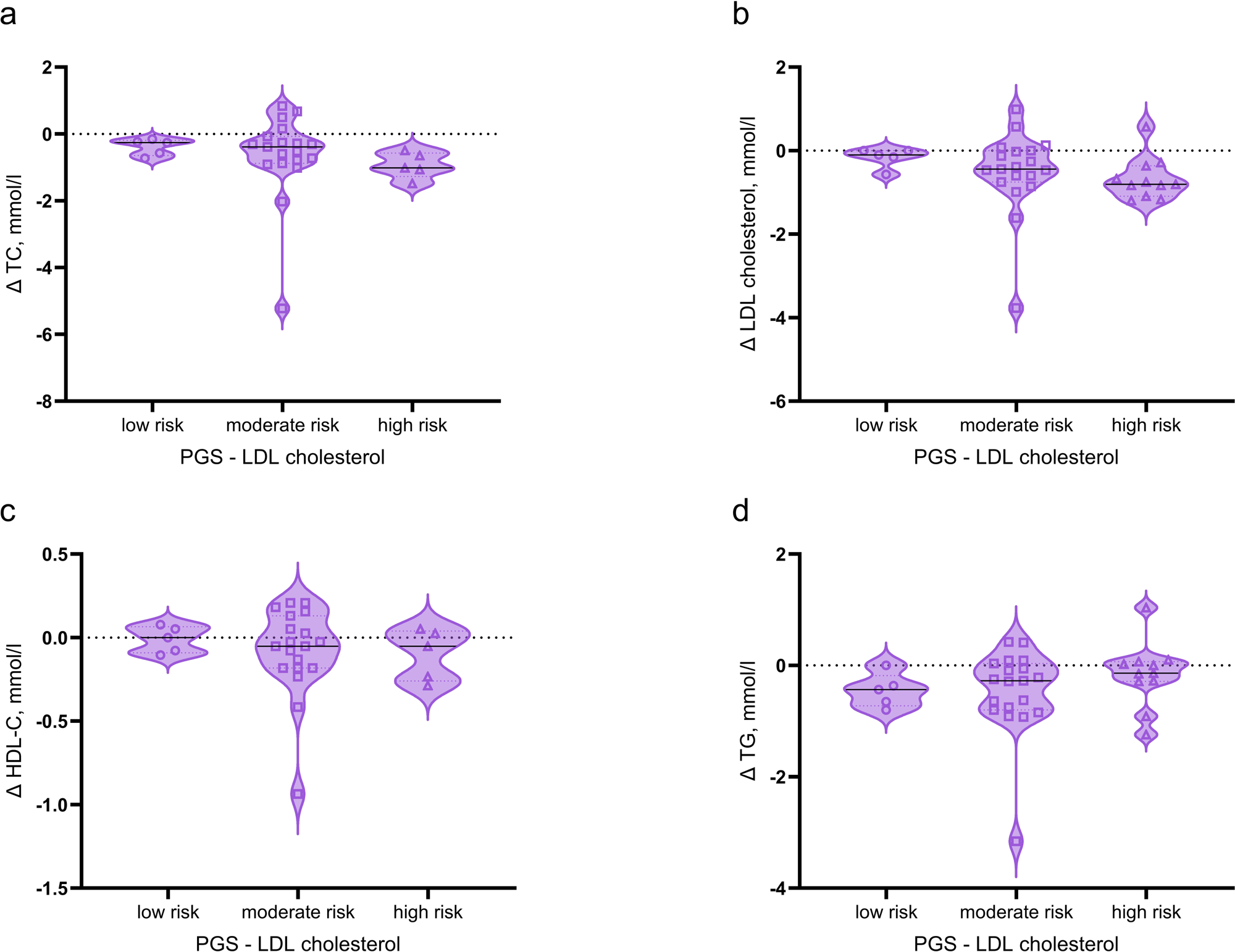
ND-induced improvements in blood lipid profile depending on the genetic predisposition Shown are the absolute changes in concentrations of total **(a)**, LDL **(b),** HDL cholesterol **(c)** and **(d)** triglyceride levels in low vs. moderate vs. high LDL-PGS groups before and after intervention Only participants in ND group. Violin plots of the absolute change at endline compared to baseline. Violin plots show the distribution of individual data points (dots), with the quartile boundaries indicated by dashed lines, the median by a solid line, and the violin extending from the minimum to the maximum observed values. HDL, high-density lipoprotein cholesterol, LDL, low-density lipoprotein cholesterol, ND, Nordic diet group, PGS, polygenic risk score; TC, total cholesterol; TG, triglycerides

To further characterize the phenotype of high vs. low genetic risk individuals by considering the gut microbiome and peripheral immune signature, immuno-phenotyping has been performed. Interestingly, the microbiome of individuals with low genetic risk for hypercholesterolemia (LDL PGS) showed a lower microbial richness compared to individuals with moderate and high genetic risk (150.7 vs. 142.6 vs. 142.5 in high-, moderate- on low-risk individuals, p = 0.002). However, the baseline abundance of the most common bacterial genera and the bacterial α-diversity (Shannon index) were similar across low-, moderate- and high-risk groups in all four PGS categories (all p > 0.05). Next, we quantified the frequencies of the major immune cell types in a subgroup of 20 male participants [60.1 ± 7 years; number of individuals with high/ moderate/ low PGS: TC-PGS (3/13/4), LDL-PGS (4/11/5), HDL-PGS (5/11/4), TG-PGS (2/15/3)] with multicolor flow cytometry (MCFC). Analysis of the main immune cell populations revealed frequencies within the expected range for this population (**Supplementary Figure S4**). High TC-PGS and HDL-PGS groups tendentially showed an increase in granulocyte to lymphocyte (GLR) ratio^40^ compared low TC-PGS and HDL-PGS groups, pointing towards a more inflammatory immune state (**Figure 7 a**). Concomitantly, in individuals with high HDL-PGS, CD8 to CD4 T cell ratio^41^ (**Figure 7 b**), a marker of senescence of the adaptive immune response, was elevated. Additionally, a decrease in circulating natural killer (NK) cells was observed in high TC-PGS and HDL-PGS groups, which can be an indicator of an underlying inflammatory state (**Figure 7 c**). In contrast, high TG-PGS and LDL-PGS groups were mainly characterized by a decrease of eosinophils (**Figure 7 d**).

**Figure 7:**
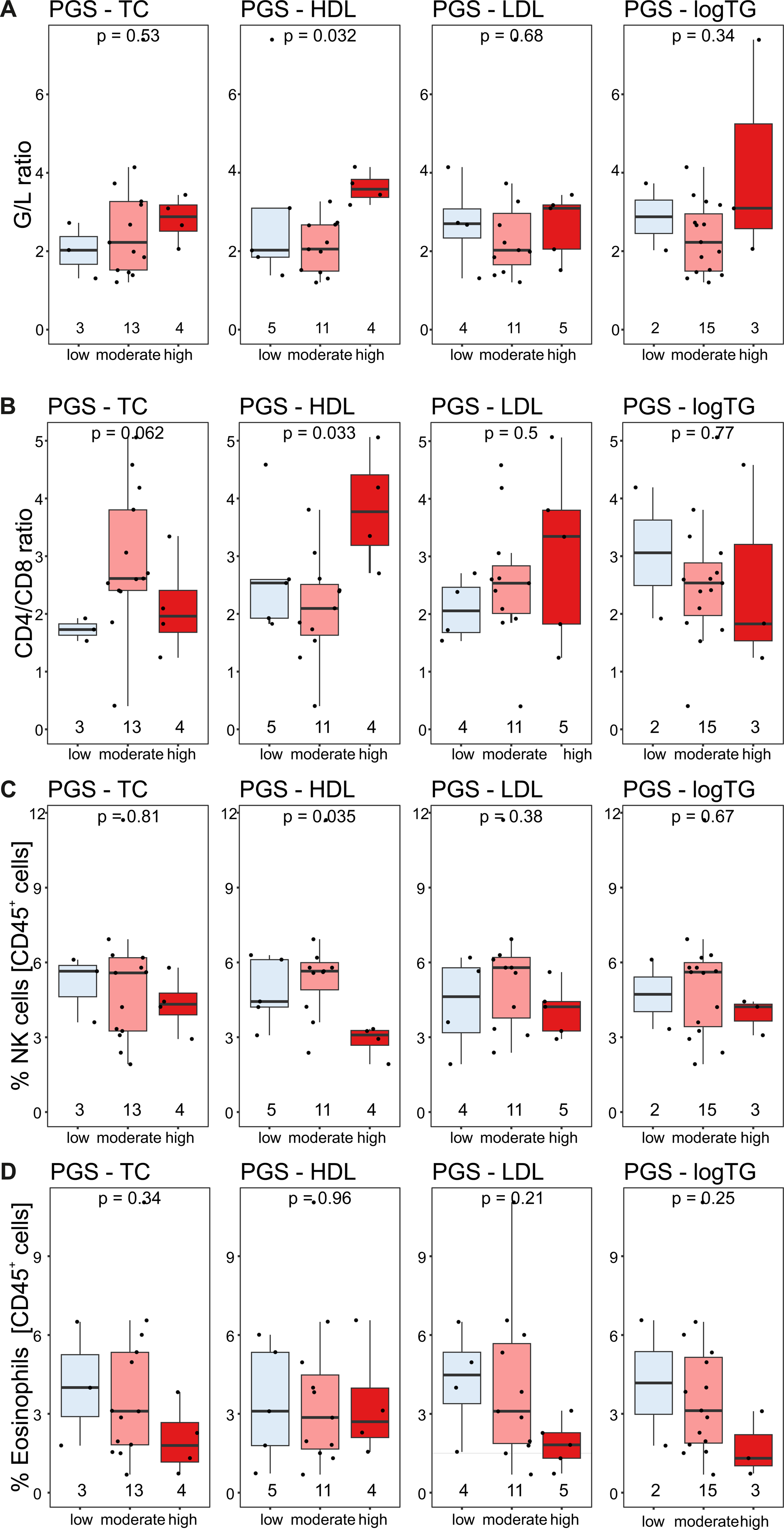
Peripheral immune system composition at baseline in individuals with high vs. low genetic risk for hyperlipidemia Boxplots for **(a)**, G/L ratio, **(b)**, CD4/CD8 T cell ratio, **(c)**, NK cell frequency and **(d)**, eosinophil frequency for each PGS grouping. Dots represent each sample distributed between the “low”, “moderate” and “high” PGS groupings for TC, HDL cholesterol, LDL cholesterol and logTG. The number of samples per group are given at the bottom of the graph; exact p value is shown G/L ratio, granulocyte to lymphocyte ratio; HDL, high-density lipoprotein cholesterol, LDL, low-density lipoprotein cholesterol; logTG, log-transformed triglycerides; PGS, polygenic risk score; TC, total cholesterol

These findings suggest that individuals with a high genetic risk for hyperlipidemia may also exhibit a distinct gut microbiome and peripheral immune signature, which may influence their individual responsiveness to dietary interventions.

## DISCUSSION

In this randomized, three-arm dietary intervention trial in obese adults, we investigated the effects of an isoenergetic 6-weeks Nordic, lacto-ovo vegetarian, or Western-type diet on lipid metabolism and gut microbiome composition. It was shown that through both, the ND and VD intervention, the initial Western-type dietary behavior shifted towards a more beneficial dietary behavior, including an increased consumption of vegetables, wholegrain products, nuts and seeds and a decreased consumption of sweets, sugar-sweetened beverages, and fast food. The Nordic diet, but not the lacto-ovo vegetarian study diet, were shown to beneficially alter lipid metabolism and, thus, reduce interims cardiometabolic risk factors and metabolic syndrome severity, especially in individuals with high-risk genetic profile for hyperlipidemia. However, in both groups, several relevant changes in plasma lipidome were observed post-intervention, including modifications of carbon chain length and degree of desaturation, which highly influence the biophysical properties of lipids. Deep phenotyping of genetic high-risk individuals revealed a reduced microbial richness, and an inflammatory immune state compared to lower-risk individuals. Several diet-induced microbial changes in the abundance of specific taxa were observed. Furthermore, integrative multivariate multi-omic analysis, appropriately reflecting the complexity of the gut microbiome, identified that the basal gut microbiome and viriome signature and function contribute to the individual responsiveness to the dietary change.

The Nordic diet intervention, which was rich in berries, root vegetables and cabbage, wholegrain oats, rye, and fatty sea fish, improved the blood cholesterol and triglyceride levels subsequently resulting in a reduction in metabolic risk as shown by a significant decrease in metabolic syndrome severity and cardio-metabolic risk scores. These beneficial effects were not observed in the participants consuming our lacto-ovo vegetarian study diet, which was rich in whole-grain wheat products, vegetables, seeds and legumes. This contrasts with two meta-analyses of RCTs where VD effectively lowered total and LDL cholesterol levels^42,43^. However, in some studies, stronger weight loss occurred in the VD intervention groups compared to the control groups, and vegan diets were included in this analyses, which may explain the discrepancy with our results. Moreover, their subgroup analysis indicated that the lipid-lowering effect of VD was less evident in obese participants than in normal-weight participants^43^.

Several studies have reported that replacing dietary saturated fatty acids (SFA) by mono- (MUFA) or polyunsaturated fatty acids (PUFA) and increasing fiber and phytosterol intake significantly improved blood lipids, especially cholesterol metabolism^44–47^. Although both VD and ND are rich in MUFA, PUFA, fiber and phytosterols, while limiting the consumption of (processed) meat, sweets and sugar-sweetened beverages compared to a Western-style diet, and are therefore considered health-promoting dietary patterns, there are major differences in diet-specific key components that may have led to the different impact on cardio-metabolic risk factors. For example, while the VD was based on olive oil, ND primarily used rapeseed oil consequently resulting in a different fatty acid composition upon both diets. In line with our findings, a recent meta-analysis of 27 intervention trials revealed that rapeseed oil alone may reduce total cholesterol, LDL cholesterol, and triglyceride levels more effectively than olive oil^48^. The only moderate lipid-lowering effect of olive oil compared to other plant oils was attributed to its fatty acid composition dominated by mono-unsaturated fatty acids (69% oleic acid, 13% SFA, 8% LA, 1% ALA^49^). In early studies, oleic acid was reported to be inferior to polyunsaturated fatty acids (PUFA) in lowering total and LDL cholesterol levels^50^. In contrast, in our study, the specific fatty acid profile of the ND, including the use of rapeseed oil and the replacement of saturated fat with polyunsaturated fat also from fatty fish, nuts, and vegetables (both n-6 and n-3), probably led to favorable effects on non-HDL cholesterol and triglyceride profiles^51–53^. Especially since it has been shown that n-3 PUFAs reduced plasma lipid levels by inhibiting very low-density lipoproteins and triacylglycerol synthesis in the liver, increasing fatty acid oxidation, and promoting the synthesis of membrane phospholipids^54–56^.

Moreover, in contrast to the ND, where low-fat milk and dairy products were consumed, the VD group consumed regular-fat milk and dairy products. While, compared to low-fat dairy, regular-fat dairy is richer in SFA, which have traditionally been associated with increased LDL cholesterol levels and CVD risk ^57–59^, we speculated that this might have contributed to the lack of cholesterol-lowering effects of the VD intervention. However, there is emerging evidence from recent clinical and observational studies challenging this assumption, reporting that the consumption of high-fat dairy foods may not adversely affect cholesterol levels or cardiovascular health^60–62^. E.g., a 4-weeks head-to-head intervention study in adults with metabolic syndrome found that a diet rich in full-fat dairy had no effects on fasting lipid profile compared with diets limited in dairy products or rich in low-fat dairy. The authors hypothesized that dairy fat, when consumed as part of complex whole foods, does not adversely affect classic CVD risk factors^63^, supporting the opinion that investigating whole foods (i.e., the food matrix) as opposed to single nutrients might be important^64,65^. Additionally, prospective cohort studies using substitutional modeling showed that the replacement of meat with dairy products as a source of SFA was associated with a lower CVD risk^66–69^, which might be relevant for our dietary switch from HD to VD.

Apart from this, other key components of the ND intervention, as berries^70,71^ rich in anthocyanin, oats^72^ rich in oat fiber, including β-glucan, and bioactive compounds^73–75^ and apples rich in proanthocyanidin^76^ were reported to exhibit cholesterol-lowering effects in humans and animal models. Since we did not observe cholesterol-lowering effects in the VD group, we concluded that these specific compounds of ND are main contributors to the beneficial properties on lipid metabolism observed in the ND group. This conclusion is supported by the results of a previous ND intervention study, where a 10-week ND intervention trial in hypercholesterolaemic adults achieved a reduction in total and LDL cholesterol of 24% and 31%, respectively^77^. Considering that these results were achieved during a hypoenergetic ND intervention accompanied by a reduction in body weight, the lipid-lowering effects of our isoenergetic ND intervention of 10% and 15% in high-risk individuals, respectively, are even more remarkable. In contrast to other ND intervention trials^77,78^, we found a reduction in fasting triglyceride concentrations, which emphasizes the significant potential of ND in the prevention of cardiometabolic events, especially in combination with the observed favorable effects on cholesterol fractions. However, also food items included in our isocaloric VD intervention, such as barley^79^, legumes^80^, flaxseeds^81^, sesame^82^ and mushrooms^83^ showed lipid-lowering properties in dietary intervention trials with a duration of four weeks to six months – yet, these interventions were accompanied by weight loss, the daily intake of these foods may have been higher than in our study phase, since these studies have primarily examined isolated foods, or were of significantly longer duration, which might explain the lack of lipid-lowering effects of our VD intervention.

Apart from the encouraging findings from routine clinical chemistry assessment, we also observed several favorable ND-induced changes in the plasma lipidomic profile, including modifications of carbon chain length and degree of desaturation of several lipid classes, both of which impact the biophysical properties of lipids^84^. ND induced a reduction in most lipid classes, while there was a significant increase in the concentration of long-chain TAGs and LPCs species, and in the number of double bonds and chain length of several lipid classes including TAG, PE, PE-O, PC, PG and SM. A longer acyl chain and a higher number of double bonds have been inversely linked to CVD risk, especially when observed in TAG lipids^85^. The observed alterations in the lipidome are in line with earlier findings from two 12-week trials with ND key foods^86,87^ and a cohort study considering fatty fish consumption^88^ and might be attributed to PUFAs from ND key lipid sources as fish, nuts, and rapeseed oil, which was also suggested by an increase in EPA (C20:5n3c) and DHA (C22:6n3c) levels in the phospholipids. This hypothesis is also supported by an earlier intervention study, where a 10-weeks supplementation of DHA-rich microalgae oil induced alterations of acyl chain composition of PE and PC shifting towards long and highly unsaturated species, which has the potential to alter membrane biophysics, such as PUFA-derived inflammation-related lipid messenger profiles^89^. Apart from that, the reductions observed in numerous lipid species have been associated with health benefits in epidemiological and intervention studies. The reduction in monounsaturated and saturated PC species might point into the direction of a reduced mortality risk^90^, while a reduction in CER species, which are mainly transported by LDL fractions^91^, could exert a positive impact on insulin resistance^92^. Moreover, the ND-induced reduction in PE species was more pronounced than in PC species, resulting in an increased PC/PE ratio compared to pre-intervention, which might have implications on parameters related to the metabolic syndrome as lipid profile and insulin resistance^93^. Remarkably, it has been reported that PC/PE ratio is lower in mice with chronic liver diseases than healthy controls, which is in line with our findings of ND-induced improvements in liver enzyme levels^94^.

In contrast, we observed even an increase in triglyceride levels from clinical routine analysis after VD intervention. However, these findings align with a lipidome study, where a 4-weeks VD resulted in higher levels of 11 triglycerides, mainly of long-chain and polyunsaturated triglyceride species^95^. Nevertheless, in our lipidomics analysis, no significant increase in TAG species was observed, suggesting that the composition of individual TAG species may not have changed. This is supported by the findings of a very recent 4–week intervention with a plant-based portfolio diet, where no significant differences compared to a control diet were observed^96^. These findings highlight the relevance of considering the quality of fat, especially triglycerides and its proportions of saturated, monounsaturated, and polyunsaturated fatty acyl content provided by the VD to appreciate its impact on blood triglyceride levels. However, the VD intervention induced modifications of carbon chain length and degree of desaturation; the normalized average of double bonds decreased for six lipid classes and the normalized average chain length decreased for five classes. Some of the PE species that increased to a slight but significant extent after VD have been linked to an increased mortality risk^90^. We therefore conclude that the characteristic fat sources and fatty acid signature of ND (fatty fish, rapeseed oil, and nuts) and VD (olive oil, dairy products, and seeds) significantly affect the lipidomic profile as well as lipid metabolism and, thus, cardiovascular risk factors. We believe that the analyzes of the diet-specific impact on the lipidome contributes to a comprehensive assessment of the metabolic health status going beyond conventional clinical measures, which might be inevitably for personalized nutrition and medicine^97^.

Furthermore, we also investigated the effect of our ND and VD interventions on gut hormone secretion, gut microbiome and viriome composition and function. In contrast to previous studies reporting a responsiveness of in GLP-1 secretion upon dietary manipulation^22,79,98–100^, including results from a 12-week intervention trial with a calorie-restricted Okinawa-based ND^101^ and MIND diet^102^, as comprehensively summarized in a recent review^98^, no changes in fasting and postprandial GLP-1 levels after ND and VD intervention were observed. In overweight adults, fasting and postprandial GLP-1 secretion was increased following a 6-week barely intervention, however, the amount of barely consumed in the intervention group might have been significantly higher than in our VD group and the intervention was accompanied by weight loss^79^. Moreover, a very recent 8-week intervention with full-fat yoghurt, which was a substantial part of our VD intervention, resulted in increased fasting, but not postprandial GLP-1 levels^103^. It might be speculated that diet-induced weight loss has a major contribution to the GLP-1 alterations previously reported, and that the duration of our interventions was too short to manifest sustainable, dynamic responses upon isocaloric dietary intake. However, in a very recent RCT, a 4-week low-energy diet failed to increase fasting and postprandial GLP-1 levels after an oral glucose tolerance test^104^. Moreover, macronutrient composition of the diet and the metabolic conditions of the participants seem to play an important role in diet-induced changes in GLP-1 secretion^98^, which makes a direct comparison of the studies difficult.

There were no significant changes in alpha and beta diversity of the gut microbiota at the end of the trial, which is in line with earlier short-term VD and ND trials^105,106^. However, as also reported in these two earlier studies, we identified several specific bacterial strains that were responsive to our dietary intervention. These diet-induced alterations might be of metabolic relevance, as it has been emphasized that even single bacterial strains could exert a decisive impact on cholesterol metabolism^107^. Although our study does not establish mechanistic causality, previous evidence suggests that baseline microbiome composition may be relevant for inter-individual differences in response to dietary intervention. Previous animal and cross-sectional studies revealed a close relationship between the human gut microbiome and lipid metabolism. It was reported that the gut microbiota regulated host cholesterol homeostasis in mice^108^ and contributed substantially to individual variations in circulating HDL cholesterol levels in a Dutch cohort^109^. Additionally, microbial diversity^110^ and several bacterial taxa (e.g., *Enterobacteriaceae* and *Streptococcus spp*.^15,109,111^) have been associated with blood lipid levels in epidemiological studies. Nevertheless, recent studies have suggested that key ND foods, such as berries^112,113^, oats^114^, and fatty fish^115^, have the potential to affect gut microbiome composition. On a taxonomic level, after ND intervention, the abundance of *Ruminococcus_E* species, which has been described as obesity-associated species in humans^116^, was lower compared to baseline and VD/ HD groups. In the VD group, pronounced changes occurred in prevalence of *Bifidobacterium adolescentis* (up-) and *Lactobacillus sakei* (down regulated). The increase in *Bifidobacterium adolescentis* might be linked to health benefits, as *Bifidobacterium adolescentis* has previously been reported to be associated with microbiota recovery after antibiotic-induced dysbiosis^117^. However, *Lactobacillus sakei* was shown to have anti-obesity effects^118^ and to increase the production of anti-inflammatory cytokines in mice^119,120^. We speculate that the observed shifts in microbe taxonomy after VD and ND intervention possibly imply that the initial ‘Westernized’ microbiome of our participants evolved towards a health-promoting microbiota profile with benefits for host metabolism, e.g., via increasing the abundance of beneficial microbes including SCFAs producers. The absence of alterations in SCFAs concentrations following the dietary intervention does not contradict this assumption, as similar findings have been reported in other studies where fecal SCFA concentrations remained unchanged or showed minor fluctuations, despite an increase in the abundance of SCFAs-producing strains^121,122^. Assuming that the amount of fecal SCFAs only accounts for 5–10% of SCFAs produced by the microbiome^123^, and active enterocytes metabolize SCFAs directly^124,125^, this may result in a different concentration of SCFAs in the systemic circulation.

Our multi-omics model suggested a selective role of the basal microbiome signature in modulating the response to the VD and ND interventions and thus, interims cardiometabolic risk factors. In total, 27 basal 16S microbial features were relevant in metabolically discriminating VD and ND groups post-intervention. In the ND group, basal *Lactococcus lactis*, *Lachnospiraceae TF01-11* sp001414325 and *Bifidobacterium animals* abundance showed a strong negative correlation with changes in GGT, non-HDL cholesterol fractions, fasting and postprandial triglycerides and short-chain TAGs (<53 C atoms), which is in line with previous literature. *Lactococcus lactis* was reported to lower total and LDL-cholesterol levels in healthy adults^126^. *Lachnospiraceae* TF01-11 was identified as a healthy diet-enriched species and associated with lower triglyceride levels in a large cross-sectional study^126^. In the VD-group, *Dorea formicigenerans, and Bacteroides togonis* abundance positively correlated with changes in GGT, non-HDL cholesterol, fasting and postprandial triglycerides and short-chain TAGs (<53 C atoms)*. Dorea formicigenerans* has been suggested as non-beneficial microbial signature with enrichments associated with prediabetes^127^, non-response to bariatric surgery^128^ and chronic inflammation in humans^129,130^. *Bifidobacterium animalis* was also clearly linked to metabolic biomarkers, as a supplementation has shown to improve anthropometric adiposity in obese adults^131^ and lipid metabolism in Caenorhabditis elegans ^132^. *Bacteroides togonis* might have possible implications in metabolism of complex carbohydrates and plant-based foods and might play a relevant role in gut homeostasis^133^.

Adding on the results from our first multi-omic model including the 16S gut microbial data, in our second multi-omic model in total seven WGS-derived bacteria/ viruses and two microbial pathways were attributed to the change in clinical outcomes and the discrimination of ND and VD post-intervention. In the ND group, relative changes in the LPSSYN-PWY: superpathway of lipopolysaccharide (LPS) biosynthesis, *Streptococcus* phage 9874*, Jahgtovirus secundus* and *Skunavirus* sv51701 inversely correlated with changes in postprandial triglycerides and shorter-chain TAGs (≤ 48 C-atoms), while being positively associated with changes in longer-chain TAGs (≥ 56 C-atoms). These were the first findings directly linking these viral species to host lipid metabolism in obese adults. LPS produced by gram-negative bacteria such as *Escherichia coli* and *Salmonella* may play a crucial role in bacteria-host interactions by modulating responses by the host immune system^134^. In an *Escherichia coli* culture, LPSSYN-PWY has been shown to be downregulated when exposed to stress^135^. Increased abundance of LPSSYN-PWY could hint to a decreased oxidative stress exposure after ND intervention. Changes in GLUDEG-I-PWY: GABA shunt pathway, whose function is the production and conservation of the supply of GABA in the central nervous system, showed strong positive correlations with changes in total and LDL cholesterol, and ALT, while *Prevotella* sp 885, *Clostridiaceae* GGB4664 SGB6452, *Cohcovirus splanchnicus* and *Canhaevirus faecalis* correlated negatively with the change in total cholesterol, and positively with NEFAs, postprandial triglycerides, PEs and short-chain TAGs. Interestingly, a decreased abundance of *Prevotella* sp 885 has recently been reported as relevant factor along the gut-kidney axis involved in renal impairment^136^. It is a remarkable finding that more viral than bacterial species were selected by our model, suggesting a distinct contribution of viruses in distinguishing the two diet groups. These included *Jahgtovirus secundus*, *Cohcovirus splanchnicus* and *Canhaevirus faecalis*, belonging to the order of CrAss-like bacteriophages, which might be reduced in intestinal and neurological disorders^137,138^. Moreover, under stress conditions, which may include dietary changes, these phages can switch to a lytic cycle, thereby exerting greater control over the bacterial microbiome compared to a stable state^139^. Thus, the phageome may be another determinant in the relationship between the gut microbiome and the host response to nutritional modifications. It might be speculated that the functional properties of the microbes, in addition to their prevalence, have a very specific and modulatory impact on changes in lipid metabolism. Overall, it became evident that additional research is needed to comprehensively grasp the observed connections between the response to a dietary intervention and distinct microbial and viral pathways and signatures, including microbial metabolites. Furthermore, for the first time, we identified a specific pattern of the gut viriome related to the metabolic response to a dietary intervention. Moreover, although our study did not profile fecal sterols or ismA genes and therefore cannot assess microbial cholesterol-to-coprostanol conversion, prior work identifies gut bacteria harboring IsmA that convert cholesterol to coprostanol and links this pathway to lipid profiles^140^; diet can modulate this conversion in a context-dependent manner. Our findings should therefore be viewed as hypothesis-generating, and future studies incorporating fecal sterol measurements and ismA quantification are warranted.

Besides the influence of the microbiome and viriome on the change in clinical markers, a strong influence of the genetic predisposition became evident in all analyses. Our analyses revealed a modifying impact of genetic predisposition on the response to the dietary intervention. Specifically, the most significant reduction in total and LDL cholesterol levels was observed in participants exhibiting the highest metabolic risk or strongest genetic predisposition (high PGS group). In all multi-omic models, mainly all PGS (total/ LDL/ HDL cholesterol and triglycerides- PGS) emerged as one of the strongest contributing factors associated with the changes in lipid metabolism. Similarly, in the early 1990s, light was shed on the effect of common gene variants on diet response; the response to a dietary riboflavin administration on blood pressure had only been observed in individuals homozygous for a specific polymorphism^141^. Furthermore, an earlier showcase study reported that the strongest EPA- and DHA-induced blood triglyceride-lowering effect was associated with the presence of a specific APOE genotype^142^. However, until now, evidence for gene-based dietary recommendations is lacking^143^. Therefore, we consider our findings that individuals at high genetic risk and with metabolic syndrome traits particularly benefit from an ND intervention, an important addition to the current research, especially in combination with the results of our multi-omics analysis. Given that high-risk individuals presented a decreased microbial richness compared to low- and medium-risk individuals, further research is needed in order to characterize the microbial signature in dependence of the genetic predisposition.

Our analyses of the peripheral immune composition suggested that individuals with high genetic risk for hypercholesterolemia are more likely to present an inflammatory state than low-risk individuals. Even if this did not reach statistical significance, presumably due to the small samples size, there was a clear trend for an immunological signature characterized by a higher granulocyte and lower T cell frequency as well as a decrease in eosinophils in individuals with high genetic risk scores compared to lower-risk individuals. These immunological signatures can likely be attributed to an increased inflammatory state and an exhaustion of the adaptive immune response. Our analysis of baseline immune cell frequencies provided first additional insights into the underlying immunological differences of this cohort, potentially contributing to a deeper understanding of the observed responses to dietary changes. As recent studies have provided proof of concept that targeting inflammation may limit the occurrence of cardiovascular events^17,144^, investigating the potential of our plant-based dietary interventions to selectively modulate immunological signatures for participants benefit would be of great value. However, our post-hoc analysis of the cholesterol levels in ND individuals pre- and post-intervention resulted in a small and uneven sample size, highlighting the need for larger intervention studies investigating the impact of genetic predisposition on the success of a dietary intervention.

### Strengths and weaknesses

A strength of this randomized, controlled, three-arm dietary intervention study is the isoenergetic design that allowed assessing ND- and VD-induced effects on clinical parameters without confounding weight-loss. Furthermore, an objective evaluation of blood and anthropometric biomarkers confirmed the compliance of the participants with respect to their test diets. Consequently, we are confident that any differences in the analyzed biomarkers and the gut microbiome composition observed after end of the trial were diet-specific effects. Currently, there are no validated dietary biomarker profiles capable of reliably distinguishing the specific dietary pattern adhered to by an individual, which is why dietary biomarkers of single nutrients or of individual foods or food groups are commonly used to assess compliance with dietary patterns^145^. Capturing the complexity of dietary patterns likely requires a panel of multiple dietary biomarkers rather than individual markers, and future work should seek to validate novel dietary biomarkers indicative of specific dietary patterns and their characteristics. Another strength of our study is that we explored the influence of diet not only on fasting concentrations of lipid fractions but also on postprandial triglycerides following a standardized test meal. This comprehensive approach enhances the estimation of individual cardiometabolic risk. Our study provides evidence that ND effectively lowers the serum concentrations of total cholesterol, LDL cholesterol, fasting triglycerides, and lipid species associated with cardiometabolic events, suggesting that ND might be a useful non-pharmaceutical tool for managing dyslipidemia, especially hypercholesterolemia and hypertriglyceridemia, and may be even superior to other treatment options that may have side effects. Another unique aspect of the present study is the direct comparison of two health-promoting dietary patterns, the Nordic and the lacto-ovo vegetarian diet, in the same population. Thus, our study design allowed an evaluation of the respective diet-induced effects on known risk factors for cardiovascular events beyond conventional clinical markers. This present study provides novel data on the influence of specific gut microbiome strains and genetic profile on a dietary challenge with a ND and VD intervention, thereby explaining the inconsistent results from earlier intervention studies in individuals with metabolic syndrome traits. Finally, our multi-omic approach allowed us to elucidate major systemic changes induced by diet explaining the underlying mechanisms. Since our study protocol comprised six weeks of intervention, conclusions regarding longer-term effects cannot be drawn and require further research effort.

## Conclusion

Through the three-arm study design comparing two distinct dietary interventions—both characterized by a high intake of fruits, vegetables, unsaturated fatty acids, secondary plant compounds and dietary fiber, as well as a low intake of (processed) meat, sweets, sugar-sweetened beverages, and fast food—we demonstrate that dietary composition leads to significant and clinically relevant differences in metabolic responses, and that these changes are associated with both the gut microbiome and genetic predisposition.

The Nordic diet was identified as an effective strategy to reduce interims cardiometabolic risk factors in obese adults with metabolic syndrome traits while individual genetic predisposition, along with specific basal and diet-altered gut microbiome signatures and pathways, emerged as key contributors of these observed effects. This novel data provide insights into the gut microbiome and lipidomic profile during dietary change and highlights important differences in microbial assembly associated with improvement in health status and metabolic responsiveness. Moreover, our results highlight that high-resolution, multi-layer omics analyses allow comprehensive characterization and potential stratification of participants for targeted prevention strategies aiming to prevent cardiometabolic diseases and contribute substantially to the concept that dietary interventions offer a broadly effective approach to enhance cardiometabolic health. Further investigation into the specific bacterial strains linked to lipid metabolism may uncover potential targets for precision microbiome-based therapies in hyperlipidemia. Long-term studies are needed to assess the sustainability of diet-induced metabolic improvements, to explore the broader implications of microbiome shifts on other aspects of health, and to achieve precision nutrition recommendations aiming to reduce cardiometabolic risk.

## Data Availability

All data produced in the present study are available upon reasonable request to the authors.

https://zenodo.org/records/10869143

http://www.ncbi.nlm.nih.gov/bioproject/1093259

## ACKNOWLEDGEMENTS

The authors would like to thank the study participants for their time and effort in participating in the trial. We thank all lab technicians of the Departments Nutrition Physiology and Nutrition and Microbiota, University Bonn and the Central Laboratory, University Hospital Bonn and the medical staff for their excellent assistance during the study and clinical microbiomics for the analysis of the 16-S derived metagenomic data.

## AUTHOR CONTRIBUTIONS

H.H., P.S. and M.C.S designed the study. H.H. and A.S. conducted the clinical trial. A.S., W.S., B.S.W., B. H., J.J.H., L.B, T.P., M.Y and C.T. performed the laboratory analysis; H.H., A.S., A.M., A.D., J.L., L.B. and L.W. performed the statistical analysis. M.C.S., P.S., M.C., P.K. and M.S. supervised the trial and laboratory and data analysis. H.H. prepared the first draft of the manuscript, which was subsequently finalized in close collaboration with A.S., A.M., A.D., W.S., M.C.S. and P.S. All authors provided substantial content contributions and edited the manuscript. H.H., A.S., A.M., A.D, J.L and L.B. created and edited the figures. All authors have read and approved the final manuscript.

## METHODS

### Study design

We conducted a randomized controlled dietary intervention trial with a parallel design over six weeks. After all participants provided written informed consent, their eligibility for study participation was determined at a screening visit. Subsequently, participants were randomly allocated (list randomization through the Institute of Medical Biometry, Informatics and Epidemiology, University Hospital Bonn, Germany) to one of the three study groups following an isoenergetic lacto-ovo VD, ND or a control group, which maintained their habitual Western-type diet (HD).

Anthropometric measurements, recording of REE by indirect calorimetry, deep metabolic characterization (including an MMTT), and collection of fecal samples were performed at baseline and at the end of the trial. At baseline, the habitual nutrition of all participants was surveyed using a food frequency questionnaire (FFQ) and whole blood samples for genetic analyses were collected. At the end of the trial, the participants were interviewed regarding adverse events and changes in their quality of life. To assess compliance with the study diets, participants continuously recorded their dietary intake (food diary), and several blood biomarkers were measured at baseline and after six weeks.

The study protocol was conducted according to the Declaration of Helsinki, approved by the local ethics committee, and registered in the German Clinical Trials Register (DRKS; http://www.germanctr.de and http://www.drks.de) under the identifier DRKS00015861.

### Participants

Overweight (BMI: 27–29.9 kg/m^2^) or obese (BMI: 30–39.9 kg/m^2^) participants (men and women aged 45–70 years) showing at least one metabolic syndrome trait (visceral fat distribution [waist circumference ≥80 cm for women, ≥94 cm for men], prehypertension [≥ 120–139 mmHg systolic and/ or ≥ 80–89 mmHg diastolic] or grade 1 hypertension [≥ 140–159 mmHg systolic and/or ≥ 90–99 mmHg diastolic], dyslipidemia [serum triglycerides ≥ 1.7 mmol/l and/ or HDL cholesterol ≤ 1.3 mmol/l for women and ≤ 1.0 mmol/l for men] and/ or a proinflammatory state [serum hsCRP ≥ 2.0 mg/l]) were recruited at the Institute for Nutrition and Food Sciences, University Bonn, through advertisement in local newspapers, on the radio, or via flyers. The exclusion criteria were as follows: smoking; diabetes mellitus type 2; chronic liver, kidney, gastrointestinal, thyroid, or inflammatory diseases; prior cardiovascular events; acute infections; recent surgeries; pregnancy or breastfeeding; fish allergy; actively practiced vegetarianism or veganism; antibiotic therapy four weeks before baseline; alcohol or drug abuse; chronic intake of dietary supplements (i.e., n-3 fatty acids, vitamin E, and magnesium); and participation in a weight loss program or in other clinical studies.

### Study diets

ND and VD study diets were designed considering characteristic food items (key components) and individual energy needs (see below). For the ND, characteristic key foods have been defined in the literature^146^, which have been included in the design of the ND in this study. A lacto-ovo vegetarian diet is primarily characterized by the exclusion of meat and fish. To avoid overlap between the dietary patterns, the key foods of the ND were not included in the VD. Instead, alternative foods from the same category, e.g., olive oil instead of rapeseed oil, regular-fat dairy products instead of low-fat dairy products, whole-grain wheat instead of whole-grain rye, pseudo cereals instead of oats etc., were used for recipe development (**Supplementary Table S1**). A variety of recipes for each meal of the day was composed and handed over in written form to the intervention group participants before starting the study period. Participants in the control group were asked to follow their habitual diet and to maintain their body weight during the entire study period. There were no specific guidelines with respect to energy intake, number of meals per day, and food choice. All participants were advised to maintain their usual physical activity during the study phase.

Using the German food and nutrient database (Bundeslebensmittelschlüssel 3.01; Max Rubner Institut, Karlsruhe, Germany: software EBISpro [Hohenheim, Germany]), the recipes for daily menus in the two intervention groups covered seven energy classes starting with 1400 kcal/d (class 0) and increasing stepwise by 400 kcal/d (max. deviation +/- 2%). All participants were individually assigned to an energy class according to their individual TEE measured during the baseline visit (REE x 1.5 for physical activity). For example, participants with daily TEE in the range of 1200–1600 kcal received a study diet that provided 1400 kcal/d. In each energy class, daily energy intake was divided into five regular meals: breakfast and dinner provided 25% of the daily intake each, lunch 40%, and two in-between meals 5% each. In both diet groups and in each energy class, ≥ 13 recipes with identical energy contents were available for breakfast, ≥ 14 for lunch, and ≥ 14 for dinner. To ensure an easy integration of the study diet into individual daily life, it was possible to eat ‘lunch’ in the evening and ‘dinner’ at lunchtime. The participants were asked to prepare their meals according to detailed recipes. In addition to the meals, the participants could choose two out of ≥ 18 available in-between meals (snacks) per day, including alcoholic beverages, (dried) fruits, dairy products, nuts, sweets, and salty snacks. If participants reported food intolerance or aversion to single food items, the respective products were replaced in the menu plan by study nutritionists with alternatives following the respective dietary pattern prerequisites. The selection of recipes and any deviations from the recipe plan were documented in a food diary during the intervention phase. Moreover, the participants received advice and assistance once a week from registered dietitians via telephone. If participants preferred *face-to-face* meetings, they were invited to meet the study personnel at the study site at the institute. Importantly, the participants were instructed to measure their body weight in light clothes twice a week and report any weight gain or loss immediately to the study personnel. If persistent fluctuations occurred (plus or minus 5% of body weight), the menu plans were increased or decreased by one energy class. A more detailed description of the study diets, including a list of the recipes provided in the ND and VD group, is provided in the **Supplemental Note** in the Supplemental Material section.

Participants in the control group completed two open 3-day dietary protocols: one within week 3 and the second within week 6. Evaluation of the protocols confirmed that the habitual diet reflected a typical Western-type food pattern characterized by high proportions of processed foods, red meat, sugar-sweetened beverages, and sweets, and a low proportion of fruit and vegetables.

### Compliance

Compliance with the study diets was assessed by filling out a short-form questionnaire to review the food choice (frequency of consuming the key food items [**Supplementary Table S1,** and see “dietary assessment section” below]) at baseline and after six weeks. Furthermore, weekly telephone calls and *face-to-face* contacts during visits to the study center and continuous recording of dietary intake (6-week food diary) were conducted to create accountability and engagement and ensure compliance with the study diets. Moreover, several blood biomarkers at baseline and after six weeks were monitored that reflect the key components of the respective study diets: plasma concentrations of retinol, α-tocopherol, β-carotene, and vitamin C reflecting vegetable and fruit intake^147^; serum phospholipid myristic acid (C 14:0) as a marker of dairy lipids^148,149^; α-linolenic acid (ALA; C18:3n3c) in serum phospholipids reflecting vegetable and plant foods intake^150^ (including flaxseeds, walnuts, vegetable oils); eicosapentaenoic acid (EPA; C20:5n3c), and docosahexaenoic acid (DHA; C22:6n3c) as a measure of fatty fish intake^151^; and oleic acid (C18:1n9c) reflecting the intake of olive oil^152^. Constant body weight was ranked as a sign of compliance with the study diets matched with the individual energy requirements. Data evaluation was based on group comparisons: participants in the ND group were expected to have higher blood concentrations of vitamins and ALA than participants in the HD group, and higher concentrations of EPA and DHA than the participants in the HD and the VD group; participants in the VD group were expected to have higher blood concentrations of vitamins and α-linolenic acid than the participants in the HD group, and higher concentrations of oleic acid and myristic acid than the participants in the ND group.

### MMTT

Fasting participants were invited to ingest 300 ml of a protein chocolate drink within three minutes (carbohydrates, 35.4 g (19 g sugar); protein, 25.3 g; fat, 7.6 g; Boost High Protein, Nestlé Health Science, Vevey, Switzerland). Venous blood samples were collected via a venous catheter before ingestion and at 15, 30, 45, 60, and 120 min postprandially to analyze the time course of serum NEFA and serum triglyceride concentrations.

### Measurements

#### Anthropometrics

Body weight and composition (fat mass and fat-free mass) were measured by air displacement plethysmography using the BodPod body composition system (Cosmed, Fridolfing, Germany). Body height was determined to the nearest 0.1 cm using a stadiometer (seca scale 704, seca GmbH and Co. KG, Hamburg, Germany). Waist circumference (WC) was determined midway between the lowest rib and iliac crest at maximal exhalation to the nearest 0.1 cm. The hip circumference was measured at the level of the greater trochanter to the nearest 0.1 cm. Subsequently, the waist-to-height ratio was calculated.

### Energy expenditure

Indirect calorimetry was used to monitor the total whole-body energy expenditure and substrate oxidation rates (QuarkRMR Device, Cosmed, Fridolfing, Germany). REE was multiplied by 1.5 (physical activity level, PAL) to calculate TEE.

### Dietary assessment

Habitual dietary intake at baseline was assessed using a validated, semi-quantitative FFQ comprising 149 food items. Participants reported their intake frequency across 10 categories (food groups), ranging from “I don’t eat it” to “5 times per day or more” from the previous year. Additional pictures of portion sizes supported participants to estimate their consumed quantity/ serving amounts^153^.

Change in consumed food groups upon dietary intervention and in the control group was assessed using an un-validated, semi-quantitative short-form FFQ comprising 29 food items. Here, we extracted the food groups from the full-length FFQ that matched the key-foods of the HD, ND and VD, amended key items that are not included in the full-length FFQ (e.g., berries or low-fat dairy product) and assessed their consumption frequency at baseline and after 6 weeks.

### Blood pressure and heart rate

During the visits, blood pressure and heart rate were measured twice after a 30-minute rest period in the sitting position using a semiautomatic blood pressure measurement device (Boso Carat professional, Bosch and Son GmBH and Co. KG, Jungingen, Germany). The mean arterial pressure (MAP), reflecting the average arterial pressure during one cardiac cycle, was calculated as (systolic blood pressure + 2 × diastolic blood pressure)/3.

### Assessment of metabolic syndrome severity and cardiometabolic risk

To assess the metabolic syndrome severity, a metabolic syndrome severity score (MSSS), which accounts for sex and race/ethnicity, was calculated as reported previously^154^: Non-Hispanic White males = −5.4559 + 0.0125 * WC − 0.0251 * HDL cholesterol + 0.0047 * systolic blood pressure + 0.8244 * ln(triglycerides) + 0.0106 * glucose Non-Hispanic White females = −7.2591 + 0.0254 * WC − 0.0120 * HDL cholesterol + 0.0075 * systolic blood pressure + 0.5800 * ln(triglycerides) + 0.0203 * glucose

The absolute 10-year CVD risk was calculated at baseline according to the Framingham risk score (FHS) incorporating age, sex, smoking habits, total and HDL cholesterol, and systolic blood pressure. Individuals with FRS scores of <10% are considered to have a low 10-year CVD risk, while those with FRS scores of 10% −20% and >20% are considered to have intermediate and high 10-year CVD risks, respectively^35^. To check for possible diet-induced effects on intermediate cardiometabolic risk, a global cardiometabolic risk index (CMRI) was calculated as an integrated measure of the cardiometabolic risk profile based on percentage body fat, WC, MAP, plasma triglycerides, plasma glucose, total cholesterol and TNF-α, using the following formula: Z-score = (value−mean) / standard deviation for each parameter. Z-scores were summed up to calculate the CMRI score^155^.

### Laboratory analysis

#### Blood sample collection/ compliance and metabolic marker analysis

Blood samples were collected in tubes containing ethylenediaminetetraacetic acid (EDTA), fluoride, or coagulation activator (Sarstedt, Nümbrecht, Germany). After 15 min centrifugation at 3000g and 8°C, plasma and serum supernatants were obtained and immediately frozen in cryovials at -80°C. Fasting glucose and insulin levels, clinical biochemistry, and hematological parameters were measured within 4h after blood sampling. For genetic analysis, whole blood samples were immediately frozen at -80°C.

Glucose plasma concentration was measured by the enzymatic reference method with hexokinase and VIS photometry (Cobas 8000 modular analyzer series, Roche Diagnostics, Basel, Switzerland). Hemoglobin A1c (HbA1c) was measured by HPLC and ion-exchange method (D-100™, BioRad, Solna, Sweden). Serum insulin concentration was determined using a chemiluminescent immunometric assay (Cobas 8000 modular analyzer series, Roche Diagnostics, Basel, Switzerland). Serum concentration of high-sensitive C-reactive protein (hsCRP) was measured using a turbidimetric immunoassay (Cobas 8000 modular analyzer series, Roche Diagnostics, Basel, Switzerland). Serum concentrations of triglycerides, total cholesterol, HDL-cholesterol, LDL-cholesterol, alanine transaminase (ALT), aspartate transaminase (AST), GGT, creatinine, uric acid, urea and total bilirubin were determined using VIS photometry (Cobas 8000 modular analyzer series, Roche Diagnostics, Basel, Switzerland) at the Central Laboratory of the Institute of Clinical Chemistry and Clinical Pharmacology, University Hospital Bonn. For serum triglycerides, the postprandial concentration during MMTT was measured at 15, 30, 60, and 120 min after drink ingestion. Method specifications are available online (https://www.ukbonn.de/ikckp/zentrallabor/leistungsverzeichnis/).

Fasting and postprandial plasma concentrations of GLP-1 at 15, 30, 45, 60, 120 and 180 min after MMTT ingestion, as primary outcome, were measured at the Department of Biomedical Sciences, University of Copenhagen, Denmark, in duplicate using a total GLP-1 NL-ELISA (10-1278-01; Mercodia, Uppsala, Sweden) according to manufacturer’s instructions^156^.

Fasting and postprandial serum concentrations of NEFA were analyzed using an in vitro enzymatic colorimetric assay (NEFA-HR, Wako Diagnostics, Mountain View, USA). Postprandial concentration was measured at 15, 30, 45, 60, and 120 min after MMTT ingestion.

At baseline and after six weeks, fatty acid composition of serum phospholipids was measured in duplicate by gas chromatography (model 3900; Varian Inc., Palo Alto, CA, USA; flame ionization detector) as described elsewhere (CV: 3.2 –19.4%)^155^. Plasma retinol, α-tocopherol, β-carotene, and vitamin C levels were measured as described previously^157,158^.

### Lipidomics analysis

All solvents used for lipidomics analysis were of high performance liquid chromatography (HPLC) or liquid chromatography-mass spectrometry (LC-MS) grade (VWR International GmbH and Merck KGaA, Darmstadt, Germany). In the first step, 2 μL plasma was mixed with 250 mL of a custom extraction solution containing chloroform/methanol (CHCl3/MeOH), mixed with internal standards covering different lipid classes, bath sonicated for 10 s, and centrifuged at 20,000 × g and room temperature (RT) for 2 min. The supernatant was mixed with 200 μL of CHCl3 and 800 μL of 1% acetic acid and centrifuged at 20,000 × g and RT for 2 min. The organic phase was transferred to a new vessel, followed by an evaporation step using a high-speed vacuum concentrator (45°C, 10 min). The lipid extract was then reconstituted in a spray buffer consisting of 2-propanol, methanol, and water in a volumetric ratio of 8:5:1, supplemented with 10 mM ammonium acetate, and sonicated for 5 min to ensure homogeneity. Finally, shotgun lipidomics were measured employing a Thermo Q Exactive Plus mass spectrometer optimized with a heated electrospray ionization II (HESI II). The mass spectrometer was set up to acquire MS1 spectra at a high resolution of 280,000 over the m/z range of 250 to 1200 in positive ionization mode. MS/MS spectra were then acquired at a resolution of 70,000 using Data Independent Acquisition (DIA) in 1 m/z windows from 250 to 1200 m/z, also in positive ionization mode. The raw data was converted to the .mzML files and analyzed using the LipidXplorer software v.1.2.8.1^159^. To ensure accurate identification of lipid species and their associated internal standards, custom Molecular Fragment Query Language (MFQL) files were used. Quantification was performed using internal standard intensities. The resulting data were normalized to the total volume of extracted plasma to facilitate comparative analysis.

### Fecal sample collection/ 16S rDNA V3V4 sequencing data

Stool samples were collected by the participants the day before their visit at baseline and after six weeks of intervention. From n=3 patients, no fecal samples were collected. Upon receipt at the study center, fecal samples were immediately stored at −80 °C until further analysis^160,161^. For analysis, samples were transfer from collection tubes to ZR BashinBead lysis tubes (0.1 and 0.5 mm, Zymo Research, Freiburg, Germany) on dry ice. Subsequently, total genomic DNA was extracted from 120 mg fecal material using the chemagic DNA Stool Kit (PerkinElmer chemagen, Baesweiler, Germany) according to the manufacturer’s instructions 133-35^162–164^. A mechanical lysis step was performed after the addition of lysis buffer using a Precellys 24 tissue homogenizer (Bertin Instruments, Frankfurt am Main, Germany). After extraction, DNA was stored at -20°C until further analysis.

Next-generation sequencing for analysis of the fecal content amplicon sequencing of the fecal microbiome was performed at Life & Brain GmbH, Bonn, Germany, as described previously ^165^ with minor modifications. Briefly, the V3V4 region of the 16S rRNA gene was amplified in the first PCR step using the primers Bakt_341F (5’-CCTACGGGNGGCWGCAG-3’) and Bakt_805R (5’-GACTACHVGGGTATCT AATCC-3’) in a 25 µL PCR reaction containing 2.5 µL template (5 ng/µL), 12.5 µL 2x KAPA HiFi HotStart ReadyMix (Roche, Mannheim, Germany), and 5 µL of corresponding primers (1 µM). PCR was performed in a thermal cycler using the following program: initial denaturation step at 95°C for 3 min, followed by 25 cycles of denaturation (30 s at 95°C), annealing (30 s at 55°C), elongation (30 s at 72°C), and a final elongation step at 72°C for 5 min. In the second PCR step, dual indices and Illumina sequencing adapters were added using a Nextera XT v2 index kit (Illumina, San Diego, CA, USA). For the second PCR reaction 25 µl 2x KAPA HiFi HotStart ReadyMix (Roche, Mannheim, Germany), 5 µL of the corresponding Nextera XT index primer, and 10 µL PCR grade water in a total volume of 50 µL was used per sample. Cycling conditions were as follows: initial denaturation at 95°C for 3 min, followed by eight cycles of denaturation (30 s at 95°C), annealing (30 s at 55°C), elongation (30 s at 72°C), and a final elongation step at 72°C for 5 min. After each PCR step, amplicon libraries were spot-checked on an Agilent TapeStation 4200 using D1000 ScreenTape (Santa Clara, CA, USA) and purified using AMPure XP beads (Beckman Coulter, Krefeld, Germany). The samples were normalized to 4 nM and equimolar concentrations were pooled.

The final pool was quantitated using the Qubit dsDNA HS assay kit from Thermo Fisher Scientific (Waltham, MA, USA), and the fragment size was checked on a D1000 ScreenTape. Sequencing was performed on an Illumina MiSeq system using the MiSeq Reagent Kit v3 to produce 2 × 300 bp sequences. Clustering was performed at 4 pM, with a 15% spike in of PhiX. Demultiplexing was performed using a MiSeq system.

In total, 227 samples had more than 10000 high quality counts and were included in the analyses. On average, 112680 (median = 112771) raw read pairs were generated. The average number of high-quality read pairs per sample was 68197 (median = 68024) with a minimum of 13703 read pairs. Of 2517 detected ASVs, 653 had a 100% hit in the reference database, and 1585 ASVs had a sequence identity of at least 99% to a reference amplicon.

### Whole genome metagenome sequencing (WGS)

In a subsample (n = 34), shotgun sequencing was performed for the determination of the whole genome metagenome in fecal samples. The Illumina DNA PCR-Free Library Prep Kit was utilized for library preparation, eliminating the need for amplification. The steps performed included tagging of genomic DNA, post tagging clean-up, ligation of indexes and clean-up of libraries according to the protocol guidelines. Purified libraries were pooled with a target molarity of 1.5 nM. The concentrations of individual libraries were determined using the ssDNA Qubit Kit. Average fragment size of 450 bp was used for every sample. Before proceeding to the next step, the pool was denatured and diluted according to the manufacturer’s instructions. Finally, the library and PhiX control were combined in a single reaction vessel with a PhiX fraction of 1%. Sequencing was performed on the NovaSeq 6000 platform using 300 cycles of S1 chemistry.

The shotgun metagenomic data underwent an initial process of filtering, demultiplexing, and adapter trimming ^166^. Subsequently, the resulting preprocessed FASTQ files were analyzed using MetaPhlAn4, HUMAnN3, and Kraken2.

MetaPhlAn (version 4.0) was used for taxonomic analysis of bacterial and archaeal compositions. MetaPhlAn utilises unique clade-specific marker genes to create a highly specific profile of microbial taxa present in metagenomic samples. For functional profiling of microbial communities, the HUMAnN software package (version 3) was employed. This package incorporates a comprehensive database of microbial genomic and metabolic information to assess the functional potential of microbiomes. Both analyses were conducted using default parameters.

Kraken2 was used to classify DNA viruses based on k-mers, relying on a user-generated database with a confidence threshold of 0.1^167^. The Dustmasker package was also used to construct the database and mask nucleotide sequences with low complexity, reducing false positives^168^. Bracken 2.7, which performs Bayesian re-estimation of abundance after classification with Kraken, was used to estimate the relative abundances of species present in the samples. Bracken uses the taxonomic assignments acquired through read classification by Kraken2 ^169^.

Additional post-hoc filtering was performed based on at least 5 counts with a 10% sample prevalence ^170^.

### Profiling of raw 16S rDNA V3V4 sequencing data

A customized pipeline based on dada2 was used for bioinformatics processing of the sequence data into an ASV (amplicon sequence variant) abundance table^171^. Primer sequences were removed from raw reads using cutadapt. Reads without primer match or with ambiguous bases (e.g. Ns), as well as reads shorter or longer than expected from the number of sequencing cycles and the length of the primers were filtered out. In an additional filtering and trimming step (dada2::filterAndTrim command), reads were trimmed at the three prime end based on sample-specific quality scores. Trimmed reads expected to contain more than one error based on the nucleotide quality scores of the nucleotides were removed. The remaining reads were dereplicated into unique sequences, and forward and reverse reads were then denoised separately for each sample. In this denoising step, a less abundant sequence may be assigned to a closely related, more abundant sequence based on comparison to a data-based error matrix. In that case, the low abundance sequence is considered a sequencing error of the more abundant sequence. Denoised forward and reverse reads were merged, thereby discarding read pairs without sufficient overlapping or with any mismatch in the overlap region. Finally, suspected bimeras (also called chimeras) were removed from the generated abundance table through internal abundance and sequence comparisons.

The taxonomic assignment of the detected ASVs was done in two steps. First, the ASV sequences were compared to full-length 16S sequences in an internal reference database (CM_16S_27Fto1492R_v2.0.0) using a naïve Bayesian classifier. The reference database was generated using in-silico amplicon extraction from the GTDB database 07-RS207, the rrnDB v5.8, and the UHGG database v2.0 and subsequent curation. In the second annotation step, the taxonomic assignments were improved using precise sequence identity percentages between the found ASVs and reference amplicons in an internal V3V4 amplicon database (CM_16S_341Fto785R_v2.0.0.rds).

### Fecal SCFAs analysis

Analysis of the fecal concentrations of SCFAs, pyruvic acid, lactic acid, acetic acid, and butyric acid) was performed at baseline and after intervention. The non-frozen samples were immediately processed when delivered to the study center, as described previously ^155^.

### Genetic analysis

DNA for genetic analyses was extracted from human whole blood samples collected at baseline using a PerkinElmer chemagen System (Baesweiler, Germany) and magnetic bead-based extraction protocol. Genotyping was performed using the Illumina Inc. Global Screening Array (GSA) v3.0+MD (San Diego, CA, USA). All laboratory procedures were performed in accordance with the manufacturer’s instructions.

### Multi-color flow cytometry analysis

In a subsample of male participants (n = 20), 1ml of fresh EDTA whole blood was treated with 10ml of RBC lysis buffer for 10 minutes (Biolegend). After RBC lysis, cells were washed with DPBS and 2 million cells were used for flow cytometry analysis. Cells were stained for surface markers in MACS Buffer with BD Horizon Brilliant Stain Buffer (Becton Dickinson) for 30 min at 4°C. To distinguish live from dead cells, the cells were incubated with LIVE/DEAD Fixable Yellow Dead Cell Stain Kit (1:1000 – Thermo Scientific). Flow cytometry analysis was performed on a BD Symphony A5 instrument (Becton Dickinson) configured with 5 lasers (UV, violet, blue, yellow-green, red). The cleanup of the flow cytometry data was done using the software FlowJoTM (version 10.10.0). The gating strategy is based on the signals for forward and site scatter as well as the live/dead staining to separate single cells events from debris and cell doublets. Leucocytes were isolated based on the CD45 expression (**Supplemental Figure S4a**) and the gated data was exported in fcs. files with compensated value and imported into the cyCONDOR ecosystem ^172^ for downstream analysis.

The cyCONDOR ecosystem (version 1.1.5)^172^ was used for cell clustering, manual annotation and statistical analysis of cell frequencies. From each sample file 50.000 random cells were imported into date specific condor objects, transformed separately using the autologicle transformation144 ^173^ to minimalize technical batch variations and merged subsequently. Principle component analysis (PCA) was performed on the expression data of the merged condor object and harmonized in respect to processing batches (harmonize_PCA functions). On the basis of the harmonized PCA, UMAP dimensionality reduction and cell clustering were performed using the integrated functions runUMAP (top_PCA = 16) and runPhenograh (top_PCA = 16, k=20), respectively, and cell types were annotated manually according to key marker expressions (**Supplemental Figure S4 b**). The donor wise cell frequencies were calculated using the implemented df_frequency function and statistical testing was performed with a Kruskal-Wallis test with the stat_means_function from the r package ggpubr using default settings.

### Outcomes and sample size calculation

The primary outcome of this study was the diet-induced change in plasma GLP-1 levels. The secondary outcomes of this study were the blood lipid profile (serum and plasma concentrations of triglycerides, NEFA, total-, LDL-, HDL cholesterol, lipidome) and liver enzyme levels of participants with metabolic syndrome traits. Further analyses of genetic predisposition, immunologic signature and gut microbiome composition were considered exploratory.

The sample size was calculated *a priori* on the base of changes of the GLP-1 data from previous intervention trials and anticipated changes in GLP-1. We expected the change in the area under the curve to be approximately normally distributed with a standard deviation of 1200 [(pmol/l)*time]. Under these assumptions, a two-sided t-test at a level of 5% would have power of at least 80% to detect a difference of 800 [(pmol/l)*time] GLP-1 between at least two of the diet groups given a total sample size of 36 participants per group. The intended test procedure with the previous three group comparison reduces the power in comparison to the simple t-test situation. Simulations showed that this loss of power could be compensated by increasing the sample size to 40 participants per group.

Further, a sample size of 120 participants (40 participants per group) would have the power to detect a difference in lipid metabolism between at least two of the diet groups (overall test) using an analysis of variance (ANOVA) at a significance level of 5% of at least 80% (expected mean changes in total cholesterol of -0.14 mmol/l in the ND group, -0.35 mmol/l in the VD group, and 0 mmol/l in the HD group compared to baseline based on previous literature ^77,174^).

### Statistical analysis

All statistical analyses were performed using SPSS (v.28.0, IBM Corp., Chicago, IL, USA), R (v.4.3.3, Boston, MA, USA) or GraphPad Prism for Windows (v.9.5.1, Boston, MA, USA). Unless otherwise described, descriptive data are presented as arithmetic mean ± standard error of the mean (SEM). For all analyses, the significance level was set at p < 0.05.

### Baseline characteristics and metabolic markers

The baseline characteristics within the three study groups were compared using a 1-factor ANOVA. Intervention-specific changes in all fasting and postprandial parameters (anthropometric, blood, and stool parameters) were tested using linear mixed-effect models (restricted maximum likelihood estimation), with group (study diet type; HD, ND, and VD), visit (baseline visit and endline visit), and group-visit interaction as fixed effects. The subject identifier was set as the random factor. Analyses of postprandial parameters additionally included the time of postprandial measures (15, 30, 45, 60, and 120 min after MMTT ingestion) and the three-way interactions (group × visit, group × time, visit × time, and group × visit × time). Diet-induced changes in all parameters were calculated using HD as the reference group. Moreover, in all regression analyses, the residuals were checked for relevant deviations from normal distribution and homoscedasticity. The results obtained from the linear mixed models are presented in terms of coefficient estimates (β) with standard errors (SE). Postprandial metabolites during MMTT were summarized by the total (incremental) area under the curve ([i]AUC) using the trapezoidal rule and analyzed using linear mixed models (effect of group, visit, and group × visit interaction). Moreover, the time course of postprandial metabolites was analyzed using linear mixed-effects models (the effect of group, visit, and group × visit interaction at each time point for time-course analysis). The fasting values of the postprandial parameters were included as covariates. Triglyceride variables were log10 transformed. For cholesterol, triglycerides and liver enzymes, p-values were corrected for multiple testing using block-wise Bonferroni correction. The change in lipidomic variables was examined using linear mixed-effects models (group × visit interaction) for lipid classes, normalized average number of double bonds and acyl chain length per lipid class (data were log10 transformed in advance). For the analysis of single lipid species linear models or zero-inflated gaussian models (adjusted for baseline values) were used, according to the presence of zero-inflation in the data and pairwise post-hoc comparisons were conducted and Bonferroni-Holm corrected. Species with less than three non-zero values for each diet x visit interaction were not analyzed. For illustration of lipidomic analyses, changes between baseline and endline values were calculated as Log2FoldChanges for each individual. P-values were adjusted for the Benjamini-Hochberg false discovery rate (FDR).

### 16S and WGS microbiome data

The beta diversity of the 16S microbiome sample set was measured with pairwise generalized Unifrac distances between all samples on ASV level with the GuniFrac function from the GuniFrac R package, with 𝛼 = 0.5. The beta diversity of the sample set was visualized with non-metric multidimensional scaling using the metaMDS function from the vegan R package. When comparing the beta diversity between intervention groups, a PERMANOVA test was performed using the adonis2 function from the vegan R package with 1000 permutations. Testing for differences in microbiome taxa abundances was performed with a linear regression framework with a compositional bias correction based on LinDA ^175^. The relative taxon abundances were transformed with a centered log ratio transformation (CLR) to account for the compositional structure of the data ^176^. To avoid dividing by zero in the transformation, and due to the sparsity of the data, only the abundances of taxa observed in a given subject were used in this model. After fitting a linear regression model with each taxon as the outcome, a bias in the coefficients arising from the compositional structure of the data is identified and adjusted for^175^. The reported effect sizes are the bias corrected coefficients from the linear models. Due to the log transformation of the data, the coefficients can be interpreted as log2 fold changes. Pairwise comparisons between the intervention groups were performed by contrasting the estimated marginal means for each intervention group with the R package emmeans. To be robust against outliers in the data, Winsorization was performed, capping the abundances at their 97th percentile. To be robust against heteroscedastic errors of the residuals, Huber-White sandwich estimator was used to estimate robust standard errors^177^. Taxa present in less than 10% of the samples were not tested. Testing for differences in microbiome taxa prevalence, i.e., whether the taxon is present or absent, was performed with a binomial generalized linear regression model. To account for differences in sequencing depth between samples, the sequencing depth per sample was included as a covariate in the model. Alpha diversity of the WGS sample subset was calculated at species level as richness and by the Shannon index. Intervention-specific changes in alpha diversity were analyzed using linear mixed effects model analogous to the clinical parameters additionally adjusted for sequencing depth. Aitchison distances were calculated as measure of beta diversity at species level using the vegdist function of the vegan R package and were compared with PERMANOVA at baseline and endline between the diet groups using the adonis2 function (999 permutations), with diet group and sequencing depth as predictors. Differential abundance analysis for bacterial species, virus species and pathway abundances were performed with linear mixed models using the LinDA framework, with baseline visit and habitual diet control group as reference groups. The prevalence and mean abundance filter options were set to 0.1 and 0.0001, respectively. P-values were adjusted using Benjamini-Hochberg with a significance threshold of FDR < 0.05.

### Genetic analyses

For genetic analysis, data were quality-controlled and imputed (Haplotype Reference Consortium^178^. After imputation, polygenic risk scores (PGS) were constructed to assess the individual genetic risk for hyperlipidemia using PRSice^179^ based on genome-wide association studies (GWAS) for total, LDL, HDL cholesterol, and total triglycerides (log-transformed) from the Global Lipids Genetics Consortium^180^. For the analysis, we used the PGS including all single-nucleotide polymorphisms (SNPs) with p-values < 0.0015 (total cholesterol), < 0.0001 (LDL cholesterol), < 0.0005 (HDL cholesterol), and < 0.005 (triglycerides), based on the default settings of the PRSice algorithm. Next, PGS was z-standardized and partitioned into three groups: low (first quintile), intermediate (second to fourth quintile), and high (fifth quintile). Linear mixed-effects regression models were conducted in the PGS subgroups to evaluate the effect of the intervention on the changes in lipid levels. P-values were corrected for multiple testing using the block-wise Bonferroni correction. No differences were observed in the mean score of all four PGS scores (total cholesterol, LDL cholesterol, HDL cholesterol, and triglycerides) across the study groups (all p > 0.05).

### Immuno-phenotyping

Differences in baseline immune cell frequencies between the respective high and low PGS groups have been tested using Wilcoxon signed-rank tests.

### Multi-omics analysis

To find the multi-omic signature associated with the ND and VD, integrative multivariate analysis was performed using the Data Integration Analysis for Biomarker discovery using Latent variable approaches for Omics studies (DIABLO) model in the R package mixOmics^39^.

Omics features associated with the diets were selected using sparse Partial Least Squares Discriminant Analysis (sPLS-DA) based on five subsets of input data: PGS for total, LDL and HDL cholesterol and triglycerides, changes in lipids species identified through lipidomics and changes in metabolic markers (fasting and postprandial NEFA and triglycerides, fasting total cholesterol, LDL cholesterol, and HDL cholesterol, and liver enzymes), changes in fecal short chain fatty acids and microbiome features. Lipidomic data was filtered (including only species with <5% zeros) and the log10 fold change was computed. The models differed in their input of microbiome variables as follows: for the 16S data, model 1 was fitted with baseline clr-transformed bacteria species abundance; for the WGS data, model 2 was fitted with changes in clr-transformed pathway counts and model 3 was fitted with baseline clr-transformed bacteria and virus species. For all microbiome-associated features, low counts were removed by a filter of 0.01% for 16S and 0.001% for WGS data before clr-transformation, respectively. Missing values for metabolic markers and fecal short chain fatty acids were imputed using mice package^181^. Four values were imputed for fasting NEFA, five for postprandial AUC of NEFA and one each for pyruvic, lactic, acetic, and butyric acid. The optimal number of omic features were selected using M-fold cross validation with 50 repeats based on balanced classification error rate (BER) and the maximum distance. The performance of the final DIABLO model was assessed on the classification error rate obtained, the area under the curve of the receiver operating characteristic (AUC) and p-value. A pairwise similarity matrix is directly obtained from the DIABLO model. The similarity measure is analogous to a correlation coefficient^182^. The results were visualized using circos plot, heatmap and loadings plot. An overview of all DIABLO models and detailed information on these models can be found in **Supplementary Table S13.** The workflow is provided in http://mixomics.org/mixdiablo/diablo-tcga-case-study/" \o "http://mixomics.org/mixdiablo/diablo-tcga-case-study/.

## Data availability

Anonymized data that support the findings of this study are available from the corresponding author upon reasonable request. The raw 16S sequencing data, raw lipidomics data and PGS data have been deposited on the Zenodo platform (https://zenodo.org/; DOI: 10.5281/zenodo.10869143). Raw WGS sequencing data has been uploaded at die BioProject database under SubmissionID SUB14339193 and BioProject ID PRJNA1093259 (http://www.ncbi.nlm.nih.gov/bioproject/1093259). Access can be requested.

## Code availability

For statistical analysis of this study, publically available R packages have been used: mixOmics^183^, vegan^184^, MicrobiomeStat^175^, lme4^185^, car^186^, tidyverse^187^, mice, glmmTMB, emmeans, pheatmap, and GUniFrac. The data used for illustration are available within the Source Data file.

