## Supplemental Material for "Differential effects of Nordic and Vegetarian diets on lipid metabolism, gut microbiome and cardiometabolic risk factors: A multi-omic perspective from a randomized clinical intervention trial"

**Supplementary Table S3**: Compliance biomarkers before and after dietary intervention.14

### Supplementary note

*Detailed description of the intervention diets*

Participants received a booklet containing recipes developed by a team of nutritionists and dietitians specifically for the purpose of this study, incorporating ≥2 intervention-group specific key foods (see Table 1) per recipe. Recipes were developed for breakfast, lunch and dinner and included the ingredients with quantities and detailed descriptions, for example, the fat content, if the item is peeled, drained, cooked, etc., as well as a step-by-step cooking guide and the recipe's difficulty level. Portion sizes were adjusted to individual energy requirements to provide an isoenergetic diet. The participants were free to choose which recipe to eat and were instructed to finish the entire portion to ensure the isoenergetic design of the diet. Lunch recipes could be used for dinner and vice versa. All selections were recorded in a 6-week food log. Deviations from the recipe and leftover foods were also documented in the food log. For special occasions such as social or business meals outside, participants were provided with a list of the key foods of their respective intervention group (see Table 1) to help align their food choices with the assigned dietary pattern. Details of meals consumed outside the home were also recorded. In addition to the three main meals per day, participants were instructed to consume two "special items," chosen from a selection of snacks such as fruits, nuts, caloric/alcoholic beverages or salty or sweet treats. Calorie-free drinks were allowed ad libitum, while caloric beverages were counted as special items.

In the following, the recipes choices for both intervention groups are reported.

**VD INTERVENTION**

**VD breakfast options (1 per day)**

1. Brown bread with quark, gouda cheese, cucumber and tomatoes (number of key items:
2. Plain yoghurt with shredded coconut, flaxseed, banana and unsweetened corn flakes
3. Amaranth (unsweetened) with sunflower seeds, pumpkin seeds, raisins, plain yoghurt and mango
4. Rice flakes (unsweetened) with sunflower seeds, pumpkin seeds, raisins, plain yoghurt and mango
5. Quark with flaxseed, banana, mango and unsweetened wheat pops
6. Rice flakes (unsweetened) with flaxseeds, sesame, dried and fresh apricots and plain yoghurt
7. Scrambled eggs with tomatoes and chives, served with whole grain wheat bread
8. Whole grain wheat bread role with quark and jam, served with kiwi and orange juice
9. Omelet with mushrooms, paprika and onions, served with whole grain wheat bread role with butter and gouda cheese
10. Home-made Banana pancakes
11. Whole grain wheat bread role with butter, gouda cheese, one fried egg, served with cucumber and tomatoes
12. Smoothie from banana, orange, mango and flaxseeds, served with whole grain wheat bread with cream cheese and jam
13. Fruit salad from peach, pineapple, grapes and oranges with plain yoghurt and flaxseeds
14. Whole grain wheat bread role with butter, gouda cheese, cream cheese and kiwi or clementine

**VD lunch options (1 per day)**

1. Curry from paprika, zucchini, green beans, chickpeas and onions with fried tofu
2. Pasta with ratatouille with paprika, zucchini, eggplant, tomatoes and mixed herbs

3. Stuffed paprika with pine nuts, olives, feta cheese, paprika and tomatoes, served with rice and a side salad with vinaigrette
4. Spätzle pasta with creamy mushrooms
5. Pizza with olives, onions and corn or paprika, tomatoes and mushrooms
6. Chili sin carne from red lentils, tomatoes, beans, corn, onions and herbs, served with flatbread
7. Roasted potatoes with feta cheese and mixed vegetables (onions, paprika, zucchini), topped with herbs and pumpkin seeds and served with herbed crème fraiche dip
8. Roasted pumpkin served with coconut rice and mango dip, topped with sesame
9. Goulash from mushrooms, paprika and onions with fried bread dumplings and parsley
10. Semolina or rice pudding with plums or cherries
11. Pan-fried potato noodles with peas and green beans in a creamy sauce, topped with grated cheese
12. Mixed salad with cucumber, tomatoes, olives, pine nuts and yoghurt dressing and potato gratin
13. Green asparagus risotto with parmesan and pine nuts
14. Sweet potato stuffed with spinach, onions, tomatoes and feta cheese

**VD dinner options (1 per day)**

1. Lamb's lettuce with oranges and feta in vinaigrette, served with whole wheat bread
2. Whole grain wheat bread with cream cheese, cress, cucumber and avocado and a small plain yoghurt with flax seeds and grapes
3. Whole grain wheat or spelt bread role with cream cheese, gouda cheese, cucumber and cornichons and one orange
4. Tomato soup with baguette
5. Mixed salad with lettuce, corn, kidney beans, onions, tomatoes, feta cheese with yoghurt dressing and baguette
6. Oriental millet salad with dried apricots, peanuts and mint, served with yoghurt dip
7. Italian noodle salad with olives, cherry tomatoes, arugula, basil pesto and parmesan
8. Pea soup with herbs and whole grain wheat or spelt bread role with butter
9. Chickpea salad with lettuce, tomatoes, cucumber, spring onions, dates and cilantro with an oriental yoghurt dressing and flatbread
10. Rice salad with eggplant, zucchini, avocado and herbs vinaigrette, served with feta or goat cheese
11. Green bean salad with vinaigrette, served with whole grain wheat or spelt bread with butter, gouda cheese and cucumber sticks
12. Lentil soup with onions, carrots, potatoes, celeriac, leek and herbs, served with whole grain wheat bread with butter
13. Brown bread with butter, gouda cheese and fried eggs, served with cucumber sticks
14. Whole grain wheat or spelt bread with butter gouda cheese and vegetable sticks (paprika, cucumber, tomatoes)

**VD special items (2 per day)**

- Alcoholic beverages (wine, beer)
- Non-alcoholic beverages (coke, lemonade, orange juice, apple juice, coffee with milk, alcohol-free beer)
- Sweets (dried fruits, cookies, chocolate, gummy bears, licorice)
- Plain yoghurt with fruits
- Salty snacks (chips, nuts, salt sticks, cheese)

**ND INTERVENTION**

**ND breakfast options (1 per day)**

1. Whole grain rye bread with cream cheese and smoked salmon
2. Oat porridge with raspberries and blueberries, topped with hazelnuts
3. Quark with crispy muesli, apples, raspberries and almonds
4. Whole grain rye bread with quark, jam and one boiled egg
5. Smoothie (strawberries, raspberries, blueberries, apple, banana) topped with almonds and walnuts
6. Muesli with plain yoghurt, apple and pears, topped with hazelnuts
7. Rye crisp bread with cottage cheese, tomatoes, cucumber, carrots, one egg and a small plain yoghurt with blueberries
8. Whole grain rye bread role with margarine, turkey breast cold cuts, cucumber and a small quark with lingonberries
9. Homemade banana pancakes, topped with quark and blueberries
10. Whole grain rye bread role with margarine, salami, gouda cheese, tomatoes, cucumber and one egg
11. Scrambled eggs with whole grain rye bread role, margarine, gouda cheese and radish
12. Whole grain rye bread role with cream cheese, gouda cheese or salami and jam
13. Whole grain rye bread with margarine, cream cheese and ham

**ND lunch options (1 per day)**

1. Potato salad with spring onions, apple, carrots and cornichon in vinaigrette, with mackerel and walnuts
2. Whole grain pasta with salmon and spinach, sour cream and parmesan
3. Potato casserole with kohlrabi, broccoli, one egg, sour cream, onions and garlic, topped with grated cheese and hazelnuts
4. Köttbullar (minced beef, onions, mustard, one egg, rapeseed oil) with cream sauce, boiled potatoes and lingonberries
5. Fish sandwich (herring in oil, mayonnaise, onions, cottage cheese, whole grain rye bread role) with side salad (lettuce, tomatoes, cucumber, vinaigrette)
6. Mashed vegetables (potatoes, carrots, parsnip, margarine, milk, parsley, sour cream) with fried sausages
7. Whole grain pasta a la Bolognese (minced beef, tomatoes, carrots, celeriac, onions, garlic, herbs) with parmesan
8. Venison fillet with roasted vegetables (potatoes, carrots, parsnip, beetroot, kohlrabi) and herbed quark dip
9. Venison stew with paprika, onions and garlic, served with potato dumplings and red cabbage
10. Turnips soup with carrots, onions, celeriac, parsley, sour cream, herbs and bacon, served with Viennese sausages
11. Tarte flambé with smoked salmon and beetroot, topped with walnuts and dill
12. Stuffed cabbage role with minced beef and onions, served with boiled potatoes and cream sauce
13. Beetroot soup with onions and garlic, served with turkey skewer
14. Pollock fillet with rice, broccoli and curry sauce
15. Brussels sprouts and pumpkin skillet with boiled potatoes and herbed quark, topped with hazelnuts
16. Warm whole-grain pasta salad with arugula, rhubarb and goat or feta cheese

**ND dinner options (1 per day)**

1. Lamb's lettuce with pear, bacon and yoghurt dressing, topped with almonds and crispbread
2. Whole grain bread with camembert and lingonberries
3. Coleslaw with rye crispbread, margarine and game salami
4. Spinach salad with carrot, radish, tomatoes, cucumber and yogurt dressing, served with crispbread
5. Potato salad with cornichons, paprika, bacon and walnuts with yoghurt dressing
6. Boiled potatoes with herbed quark and smoked salmon
7. Scrambled eggs with salmon and rye bread
8. Rye bread with margarine, gouda cheese and radishes, served with cream herring
9. Carrot and apple salad with walnuts and vinaigrette, served with crispbread and cream cheese
10. Salmon tartare with rye bread and a horseradish and cream cheese spread
11. Lamb's lettuce with beetroot, apple, goat cheese and walnuts with vinaigrette, served with crispbread
12. Carrot and parsnip soup with hazelnuts, bacon and crispbread with margarine
13. Fried egg with rye bread roll and horseradish herring, served with tomatoes and cucumber
14. Casserole made from quark and semolina, with berries and almond crumble topping
15. Whole grain pasta salad with spinach, radishes and carrots, dressed with mayonnaise
16. Rye bread role with margarine and gouda cheese, served with vegetable sticks (carrots, kohlrabi, radishes) and herbed quark dip

**ND special items (2 per day)**

- Alcoholic beverages (wine, beer)
- Non-alcoholic beverages (coke, lemonade, orange juice, apple juice, coffee with milk, alcohol-free beer)
- Sweets (dried fruits, cookies, chocolate, gummy bears, licorice)
- Plain yoghurt with fruits
- Salty snacks (chips, nuts, salt sticks, cheese)

### Extended Data Figures

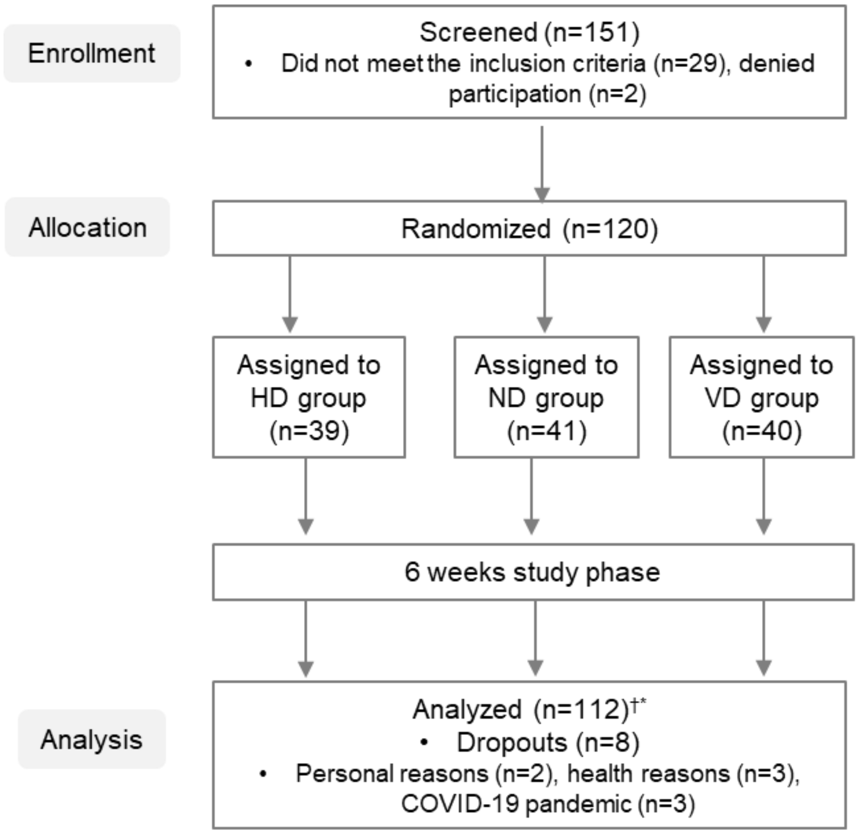

**Extended Data Figure S1:** CONSORT diagram showing participants flow through the trial

A total of 151 individuals were screened and 31 were excluded as they did not meet the inclusion criteria. 120 participants were randomized into one of three groups: ND, VD, or control (HD). Baseline measures were assessed after randomization. At the end of the 6-week intervention period, there were 38 completers in the HD group, 39 completers in the ND group and 35 completers in the VD group

† For n = 3, no stool sample could be obtained, microbiome analysis was performed for n = 117 at baseline and n = 108 at endline; WGS of the gut microbiome was conducted for a subgroup of n = 34 individuals; immuno-phenotyping was performed for a subgroup of n = 21 individuals

*for n = 7, no analysis of fecal SCFA concentrations could be performed, SCFA analysis was performed for n = 113 at baseline and n = 104 at endline

CONSORT, consolidated standards of reporting trials; HD, habitual diet; ND, Nordic diet; SCFA, short-chain fatty acids; VD, vegetarian diet; WGS, whole genome sequencing

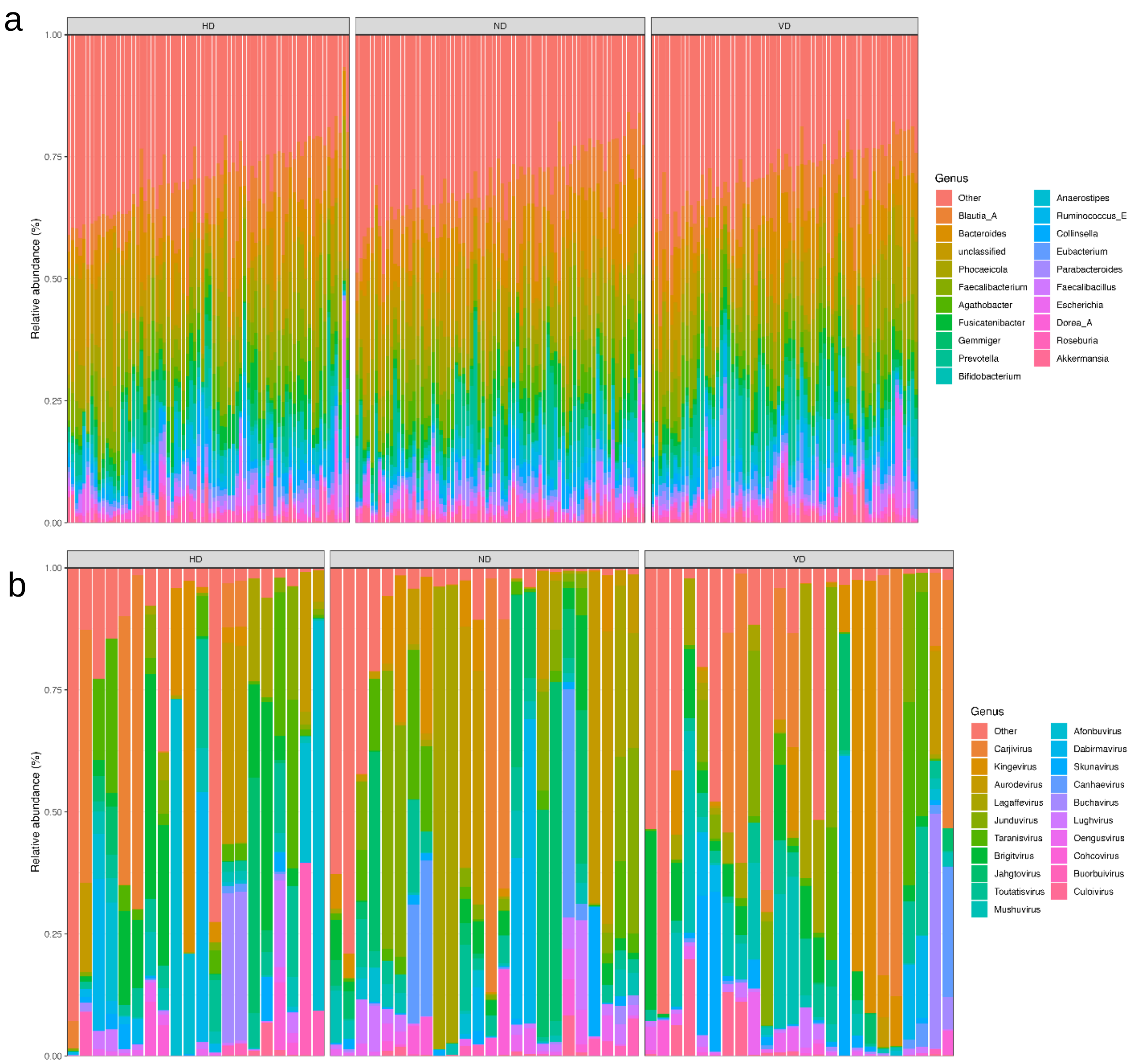

**Extended Data Figure S2**: Individual taxonomy barplots in HD, VD and ND groups

Displayed are the top 20 abundant genera for (a) bacteria derived by 16S sequencing (n = 105) and (b) viruses derived by whole-genome sequencing (n = 31) in the three interventions groups. Presented is the individual relative bacterial abundance before (left bar out of 2 parallel arranged bards for each individual) and after the end of the trial (right bar out of 2 parallel arranged bars for each individual)

ND, Nordic diet; VD, vegetarian diet

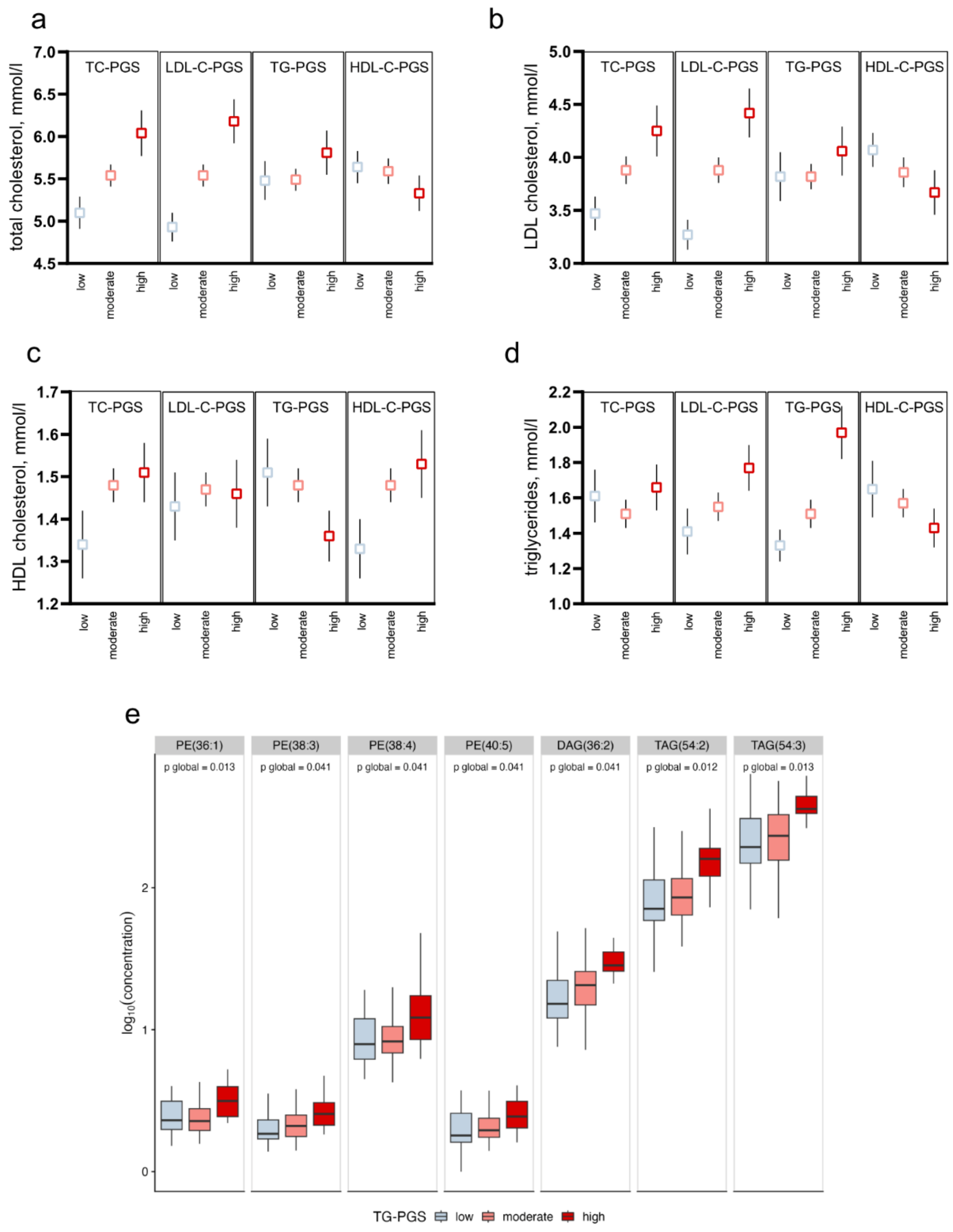

**Extended Data Figure S3**: Lipid profile at baseline in individuals with low, moderate and high risk for hyperlipidemia

Presented are the baseline concentrations of total **(a)**, LDL **(b)**, HDL cholesterol **(c)** and triglycerides **(d)** in low, moderate and high TC-PGS, LDL-PGS. HDL-PGS and TG-PGS groups. In **(e)**, lipidomic species are shown that are significantly different in low, moderate and high TG-PGS groups

In a-d, mean and SEM are presented. In e, boxplots show 25th percentile, median and 75th percentile, whiskers are Q1-1.5*IQR and Q3+1.5*IQR, respectively.

DAG, diacylglycerols; HDL-C, high-density lipoprotein cholesterol; LDL-C, low-density lipoprotein cholesterol; ND, Nordic diet group; NEFA, non-esterified fatty acids; PE, phosphatidylethanolamines; PGS, polygenic risk score; TAG, triacylglycerols; TC, total cholesterol; TG, triglycerides; VD, vegetarian diet group

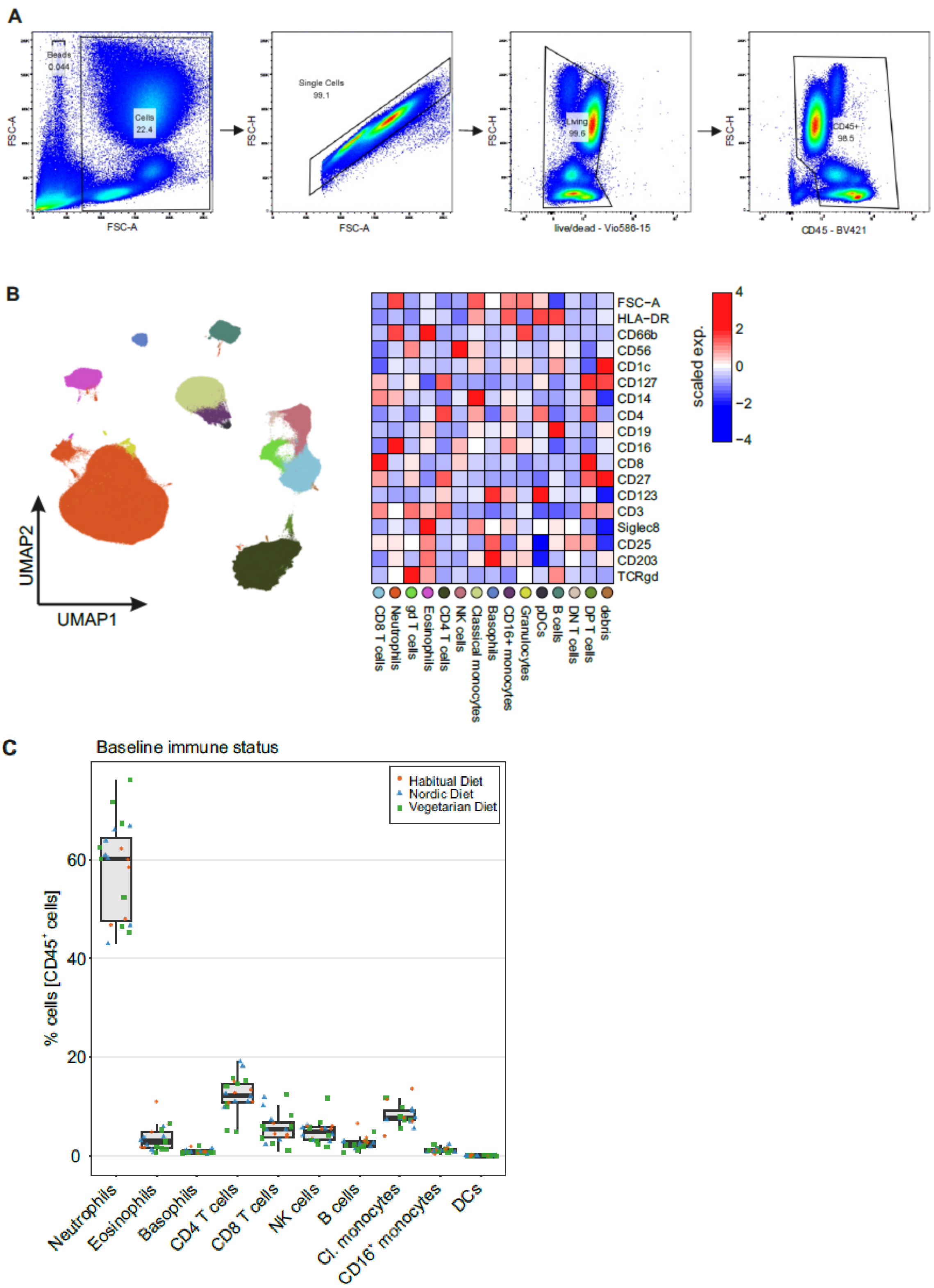

**Extended Data Figure S4**: Processing and analysis of flow cytometry data

Shown are **(a)** the gating strategy for the selection of CD45+ cells; **(b)** the UMAP of annotated cell clusters (left) with corresponding scaled marker expression (right, transformed values); **(c)** boxplot representing the frequency of the main immune cell types, individual dots are colored and shaped according to the diet showing no baseline difference between groups (n=20)

CD, cluster of differentiation; DC, dendritic cells; NK cells, natural killer cells; UMAP, uniform manifold approximation and projection

### Supplementary Tables

**Supplementary Table S1**: Key foods of the study diets

| Food types | Nordic diet | Vegetarian diet |
|---|---|---|
| Beverages | Water and unsweetened tea or coffee | Water and unsweetened tea or coffee |
| Dairy products, eggs | Milk (max. 1.5% fat), plain yoghurt (max. 1.5% fat), quark (max. 10 % fat) cream cheese, cottage cheese, goat cheese, camembert, gouda cheese, parmesan, sour cream, boiled or fried eggs | Milk (3.5-3.8% fat), plain yoghurt (3.5-3.8% fat), Greek yoghurt (10% fat), feta cheese, mozzarella cheese, gouda cheese, parmesan, sour cream, boiled or fried eggs |
| Oil and fat | Rapeseed oil, margarine | Olive oil, butter |
| Fruits | Blueberries, strawberries, blackberries, raspberries, lingonberries (≥ 3 portions/ week), apples, peas | Orange, apricots, banana, watermelon, kaki, clementine, pineapple, grapes, plums, cherries, coconut, mango, kiwi, peaches, dates |
| Legumes | Lentils | Red, brown and black lentils, chickpeas, kidney beans, green peas |
| Nuts and seeds | ≥ 3 portions/ week<br>Walnuts, almonds, hazelnuts | Cashews, peanuts, sesame, pine nuts, pumpkin seeds, sunflower seeds, flaxseeds |
| Starchy food items | Bread and bread rolls from whole grain spelt and rye, crisp bread, (whole grain) pasta, boiled potatoes | Bread and bread rolls from whole grain wheat, (whole grain) rice, (whole grain) pasta, sweet potatoes, millet |
| Cereals | Oat flakes | Wheat bran, rice flakes, unsweetened puffed wheat, unsweetened cornflakes, quinoa, amaranth |
| Vegetables | Spinach, lamb's lettuche, chard, leeks, onions, green onions, savoy cabbage, cauliflower, brussels sprouts, white and red cabbage, broccoli, kohlrabi, pumpkin, turnips, carrots, salsify, beets, parsnip, parsley, celery root, radishes, horseradish | Tomatoes, cucumber, paprika, eggplant, various types of salad greens, mushrooms, zucchini, green beans, corn, avocado, onions, soybeans, pumpkin, olives, asparagus |
| Fish | ≥ 3 portions/ week<br>Salmon, herring, mackerel | NA |
| Meat products and poultry | Chicken, turkey, venison, ham, minced beef | NA |
| Snacks | ≤ 2 small portions/ day<br>caloric and alcoholic beverages, sweet snacks (chocolate, oatmeal cookies, gummy bears), salty snacks (salted nuts, chips), snacks (low-fat yogurt with berries and nuts) | ≤ 2 small portions/ day<br>caloric and alcoholic beverages, sweet snacks (chocolate, cookies, gummy bears), salty snacks (salted nuts, chips), snacks (high-fat yogurt with fruits and nuts) |
| Condiments | Fresh herbs, ginger, garlic | Fresh herbs, garlic, cress |

The key foods were combined into recipes for each meal of the day (including quantities and detailed cooking instructions) and provided to the participants in written form before the start of the study. The participants prepared the daily meals themselves at home according to the recipes

**Supplementary Table S2**: Dietary data from FFQ[a]

| | Total (n=120) | HD (n=39) | ND (n=41) | VD (n=40) | p-value[b] |
|---|---|---|---|---|---|
| Total energy, kJ/d | 10068.2 ± 338.4 | 10800.9 ± 697.0 | 9922.0 ± 575.4 | 9503.6 ± 467.0 | 0.287 |
| Total protein, g/d | 83.4 ± 2.8 | 88.8 ± 5.8 | 82.4 ± 4.4 | 79.2 ± 4.1 | 0.363 |
| Total fat, g/d | 114.6 ± 4.0 | 123.2 ± 8.2 | 111.7 ± 6.8 | 109.2 ± 5.6 | 0.320 |
| Total carbohydrates, g/d | 228.3 ± 8.6 | 241.4 ± 18.0 | 225.8 ± 13.5 | 218.1 ± 13.3 | 0.543 |
| Total fibre, g/d | 21.0 ± 0.7 | 21.1 ± 1.2 | 21.1 ± 1.2 | 20.7 ± 1.1 | 0.962 |
| Total alcohol, g/d | 15.8 ± 1.5 | 19.1 ± 2.5 | 16.6 ± 3.2 | 11.7 ± 2.0 | 0.132 |

[a] Data are shown as mean ± SEM, n = 120

[b] Compared using one factor ANOVA

FFQ, food frequency questionnaire; HD, habitual diet group; ND, Nordic diet group; VD, vegetarian diet group

**Supplementary Table S3**: Dietary data from short-form FFQ before and after the intervention[a]

| | Habitual diet | | | Nordic diet | | | Vegetarian diet | | | |
|---|---|---|---|---|---|---|---|---|---|---|
| **Food items** | **before** | **after** | **Δ** | **before** | **after** | **Δ** | **before** | **after** | **Δ** | **p global[b]** |
| Olive oil (portion/d) | 0.8 ± 0.1 | 0.8 ± 0.1 | 0.0 ± 0.1 | 0.8 ± 0.1 | 0.2 ± 0.1 | -0.6 ± 0.1 | 0.8 ± 0.1 | 1.5 ± 0.1 | 0.7 ± 0.2 | <0.001 |
| Rapeseed oil (portions/d) | 0.5 ± 0.1 | 0.6 ± 0.1 | 0.1 ± 0.1 | 0.6 ± 0.1 | 1.3 ± 0.1 | 0.7 ± 0.1 | 0.5 ± 0.1 | 0.1 ± 0.0 | -0.4 ± 0.1 | <0.001 |
| Oil other than olive oil or rapeseed oil (portions/d) | 0.2 ± 0.0 | 0.4 ± 0.1 | 0.2 ± 0.1 | 0.3 ± 0.1 | 0.1 ± 0.1 | -0.2 ± 0.1 | 0.4 ± 0.1 | 0.1 ± 0.1 | -0.28 ± 0.1 | <0.001 |
| Butter (portions/d) | 0.8 ± 0.1 | 0.8 ± 0.1 | 0.0 ± 0.1 | 0.7 ± 0.1 | 0.1 ± 0.0 | -0.7 ± 0.1 | 0.8 ± 0.1 | 1.1 ± 0.1 | 0.3 ± 0.2 | <0.001 |
| Margarine (portions/d) | 0.3 ± 0.1 | 0.3 ± 0.1 | 0.0 ± 0.0 | 0.2 ± 0.1 | 1.0 ± 0.1 | 0.7 ± 0.2 | 0.2 ± 0.1 | 0.1 ± 0.0 | -0.2 ± 0.1 | <0.001 |
| Tomato, paprika, eggplant, zucchini, cucumber, corn, or mushrooms (portions/d) | 1.0 ± 0.1 | 1.1 ± 0.1 | 0.1 ± 0.1 | 1.1 ± 0.1 | 0.7 ± 0.1 | -0.5 ± 0.1 | 0.9 ± 0.1 | 1.5 ± 0.1 | 0.6 ± 0.2 | <0.001 |
| Root vegetables (portions/d) | 0.7 ± 0.1 | 0.7 ± 0.1 | 0.0 ± 0.1 | 0.7 ± 0.1 | 0.9 ± 0.0 | 0.2 ± 0.1 | 0.7 ± 0.1 | 0.3 ± 0.1 | -0.4 ± 0.1 | <0.001 |
| Green leafy vegetables (portions/d) | 0.6 ± 0.1 | 0.6 ± 0.1 | -0.0 ± 0.1 | 0.6 ± 0.1 | 0.6 ± 0.0 | -0.0 ± 0.1 | 0.7 ± 0.0 | 0.5 ± 0.0 | -0.1 ± 0.1 | 0.558 |
| Exotic fruits (portions/d) | 0.9 ± 0.1 | 0.9 ± 0.1 | 0.0 ± 0.1 | 0.8 ± 0.1 | 0.5 ± 0.1 | -0.3 ± 0.1 | 0.8 ± 0.1 | 0.9 ± 0.1 | 0.1 ± 0.1 | 0.026 |
| Apples, pears (portions/d) | 0.6 ± 0.1 | 0.7 ± 0.1 | 0.0 ± 0.1 | 0.7 ± 0.1 | 0.8 ± 0.1 | 0.1 ± 0.1 | 0.7 ± 0.1 | 0.4 ± 0.1 | -0.3 ± 0.1 | 0.004 |
| Berries (portions/d) | 0.4 ± 0.1 | 0.4 ± 0.1 | 0.0 ± 0.1 | 0.6 ± 0.1 | 1.1 ± 0.1 | 0.6 ± 0.1 | 0.5 ± 0.1 | 0.1 ± 0.0 | -0.4 ± 0.1 | <0.001 |
| Legumes (portions/d) | 0.2 ± 0.0 | 0.2 ± 0.0 | 0.0 ± 0.0 | 0.2 ± 0.0 | 0.1 ± 0.0 | -0.1 ± 0.0 | 0.2 ± 0.0 | 0.8 ± 0.3 | 0.6 ± 0.3 | 0.008 |
| Nuts (not salted) (portions/d) | 0.9 ± 0.3 | 0.5 ± 0.2 | -0.4 ± 0.4 | 0.8 ± 0.3 | 1.9 ± 0.5 | 1.1 ± 0.5 | 0.7 ± 0.3 | 0.4 ± 0.2 | -0.3 ± 0.4 | 0.027 |
| Pork, beef or lamb (portions/d) | 0.5 ± 0.0 | 0.5 ± 0.0 | 0.0 ± 0.0 | 0.4 ± 0.0 | 0.3 ± 0.0 | -0.2 ± 0.0 | 0.4 ± 0.0 | 0.0 ± 0.0 | -0.4 ± 0.0 | 0.232 |
| Game meat (portions/d) | 0.0 ± 0.0 | 0.1 ± 0.0 | 0.0 ± 0.0 | 0.1 ± 0.0 | 0.1 ± 0.0 | 0.0 ± 0.0 | 0.1 ± 0.0 | 0.0 ± 0.0 | -0.1 ± 0.0 | 0.049 |
| Cold cuts/ processed meat (portions/d) | 0.5 ± 0.0 | 0.4 ± 0.0 | -0.0 ± 0.0 | 1.1 ± 0.4 | 0.3 ± 0.0 | -0.8 ± 0.4 | 0.4 ± 0.1 | 0.0 ± 0.0 | -0.4 ± 0.1 | 0.061 |
| Poultry or chicken (portions/d) | 0.3 ± 0.0 | 0.3 ± 0.0 | 0.1 ± 0.0 | 0.4 ± 0.0 | 0.2 ± 0.0 | -0.1 ± 0.0 | 0.3 ± 0.0 | 0.0 ± 0.0 | -0.3 ± 0.0 | <0.001 |
| Fish (portions/d) | 0.2 ± 0.0 | 0.2 ± 0.0 | 0.0 ± 0.0 | 0.4 ± 0.2 | 1.6 ± 0.4 | 1.2 ± 0.5 | 0.2 ± 0.0 | 0.0 ± 0.0 | -0.2 ± 0.0 | 0.001 |
| Sea food (portions/d) | 0.1 ± 0.0 | 0.1 ± 0.0 | -0.0 ± 0.0 | 0.1 ± 0.0 | 0.0 ± 0.0 | -0.0 ± 0.0 | 0.1 ± 0.0 | 0.0 ± 0.0 | -0.1 ± 0.0 | 0.178 |
| Sugar sweetened beverages (portions/d) | 0.4 ± 0.1 | 0.4 ± 0.1 | 0.0 ± 0.1 | 0.2 ± 0.0 | 0.1 ± 0.0 | -0.1 ± 0.0 | 0.4 ± 0.1 | 0.3 ± 0.1 | -0.1 ± 0.1 | 0.646 |
| Wine (portions/d) | 0.4 ± 0.1 | 0.4 ± 0.1 | -0.1 ± 0.0 | 0.4 ± 0.1 | 0.3 ± 0.0 | -0.2 ± 0.0 | 0.3 ± 0.1 | 0.3 ± 0.1 | -0.0 ± 0.0 | 0.336 |
| Sweets (portions/d) | 1.5 ± 0.4 | 1.4 ± 0.4 | -0.1 ± 0.5 | 1.6 ± 0.4 | 0.7 ± 0.2 | -0.9 ± 0.5 | 1.5 ± 0.5 | 0.8 ± 0.3 | -0.7 ± 0.5 | 0.503 |
| Whole grain rye, spelt, barley, or oats (portions/d) | 1.8 ± 0.5 | 1.7 ± 0.5 | -0.2 ± 0.4 | 1.9 ± 0.5 | 3.2 ± 0.6 | 1.3 ± 0.8 | 1.8 ± 0.5 | 0.3 ± 0.1 | -1.6 ± 0.5 | 0.004 |
| Whole grain wheat (portions/d) | 0.6 ± 0.2 | 1.1 ± 0.4 | 0.5 ± 0.5 | 1.1 ± 0.4 | 0.4 ± 0.1 | -0.7 ± 0.4 | 1.0 ± 0.3 | 2.8 ± 0.6 | 1.8 ± 0.7 | 0.004 |
| Potatoes (portions/d) | 0.4 ± 0.0 | 0.4 ± 0.0 | -0.0 ± 0.0 | 0.4 ± 0.0 | 0.5 ± 0.0 | 0.1 ± 0.0 | 0.5 ± 0.0 | 0.4 ± 0.0 | -0.0 ± 0.0 | 0.027 |
| Other sources of starch (e.g. noodles, rice) (portions/d) | 0.2 ± 0.0 | 0.1 ± 0.0 | -0.1 ± 0.0 | 0.1 ± 0.0 | 0.0 ± 0.0 | -0.1 ± 0.0 | 0.2 ± 0.0 | 0.2 ± 0.0 | 0.0 ± 0.1 | 0.161 |
| Eggs (portions/d) | 0.5 ± 0.0 | 0.5 ± 0.0 | -1.3 ± 0.0 | 0.5 ± 0.0 | 0.5 ± 0.1 | 0.1 ± 0.1 | 0.5 ± 0.0 | 0.7 ± 0.1 | 0.19 ± 0.1 | 0.156 |
| Low-fat dairy products (portions/d) | 1.9 ± 0.5 | 1.5 ± 0.5 | -0.4 ± 0.3 | 1.5 ± 0.4 | 1.9 ± 0.5 | 0.4 ± 0.6 | 1.8 ± 0.5 | 0.2 ± 0.1 | -1.5 ± 0.6 | <0.001 |
| Full-fat dairy products (portions/d) | 1.1 ± 0.4 | 0.7 ± 0.3 | -0.4 ± 0.5 | 0.4 ± 0.1 | 0.1 ± 0.0 | -0.3 ± 0.1 | 0.5 ± 0.1 | 2.8 ± 0.7 | 2.4 ± 0.8 | 0.026 |

[a] Data are shown as mean ± SEM, n = 120

[b] Compared using linear mixed models

FFQ, food frequency questionnaire

**Supplementary Table S3**: Compliance biomarkers before and after dietary intervention[a]

| | **Habitual diet** | | **Nordic diet** | | **Vegetarian diet** | | **p-values** | |
|---|---|---|---|---|---|---|---|---|
| | before | after | before | after | before | after | p-value global (interaction)[b] | p-value ND vs VD[c] |
| **Fatty acid composition of serum phospholipids** | | | | | | | | |
| Myristic acid (C 14:0), µM/l | 25.26±1.33 | 27.05±2.89 | 27.13±1.43 | 24.67±1.86 | 27.11±1.61 | 29.60±1.48 | 0.142 | |
| Oleic acid (C 18:1n9c), µM/l | 308.34±10.70 | 329.88±35.63 | 328.28±13.63 | 271.69±11.60 | 304.92±10.64 | 331.20±12.35 | 0.008* | 0.010* |
| Alpha-linolenic acid (C 18:3n3c), µM/l | 9.09±0.71 | 10.02±1.66 | 9.62±0.91 | 10.17±0.86 | 9.15±0.96 | 9.67±0.63 | 0.958 | |
| EPA (C 20:5n3c), µM/l | 49.73±3.06 | 52.49±4.42 | 61.72±4.78 | 98.14±6.32 | 51.14±3.78 | 47.61±3.78 | <0.001*** | <0.001*** |
| DHA (C 22:6n3c), µM/l | 136.47±6.93 | 135.98±6.53 | 147.13±6.24 | 197.12±9.63 | 132.71±7.15 | 121.05±7.25 | <0.001*** | <0.001*** |
| **Plasma vitamin concentrations** | | | | | | | | |
| Plasma retinol, ng/ml | 632.67±24.94 | 648.38±25.12 | 664.51±25.31 | 647.93±22.53 | 600.46±20.39 | 607.67±21.51 | 0.345 | |
| Plasma α-tocopherol, ng/ml | 16626.73±539.93 | 16965.52±546.60 | 19482.70±777.05 | 18015.90±715.94 | 17927.04±567.02 | 18198.07±609.55 | 0.003** | 0.003** |
| Plasma β-carotene, ng/ml | 496.39±48.31 | 561.95±53.40 | 583.28±47.53 | 736.01±58.12 | 540.18±43.31 | 614.46±48.26 | 0.272 | |
| Plasma vitamin C, mg/L | 9.54±0.43 | 10.24±0.40 | 10.20±0.40 | 10.53±0.38 | 9.97±0.41 | 10.24±0.45 | 0.798 | |

[a] Data are shown as mean ± SEM; baseline: HD, n = 39; ND, n = 41; VD, n = 40; endline: HD, n = 38; ND, n = 39; VD, n = 35

Compared using linear mixed effect models, [b] p-value global refers to the null hypothesis that the change in outcome parameters over time (visits) is identical across all groups (interaction term of group × visit); [c] shows the p-value referring to the two-group comparison VD vs. ND (only available if p global was <0.05).

DHA, Docosahexaenoic acid; EPA, Eicosapentaenoic acid; HD, Habitual diet group; ND, Nordic diet group; VD, Vegetarian diet group

**Supplementary Table S4**: Anthropometrics and resting energy expenditure at baseline and at the end of the trial[a]

| | **Habitual diet** | | **Nordic diet** | | **Vegetarian diet** | | **p-value global[b]** |
|---|---|---|---|---|---|---|---|
| | before | after | before | after | before | after | |
| Body weight, kg | 93.6 ± 2.1 | 93.5 ± 2.3 | 89.5 ± 2.2 | 87.7 ± 2.1 | 91.9 ± 2.5 | 89.8 ± 2.8 | 0.082 |
| BMI, kg/m² | 30.9 ± 0.5 | 30.8 ± 0.6 | 31.0 ± 0.5 | 30.7 ± 0.6 | 31.5 ± 0.6 | 30.6 ± 0.6 | 0.065 |
| Waist circumference, cm | 105.8 ± 1.7 | 105.1 ± 1.7 | 105.0 ± 1.4 | 103.7 ± 1.5 | 108.0 ± 1.5 | 106.4 ± 1.7 | 0.584 |
| REE, kcal/ d | 1811.7 ± 47.3 | 1773.0 ± 45.6 | 1722.2 ± 44.6 | 1616.1 ± 39.8 | 1628.6 ± 45.3 | 1637.7 ± 51.8 | 0.043* |
| Body fat, % | 38.9 ± 1.2 | 38.7 ± 1.2 | 41.2 ± 1.2 | 40.6 ± 1.3 | 44.1 ± 1.3 | 42.5 ± 1.3 | 0.094 |

[a] Data are shown as mean ± SEM; baseline: HD, n = 39; ND, n = 41; VD, n = 40; endline: HD, n = 38; ND, n = 39; VD, n = 35. Before refers to the respective measurement at baseline, after represents the respective measurement at the end of the 6-weeks trial

[b] Compared using linear mixed effect models, p-value global refers to the null hypothesis that the change in outcome parameters over time (visits) is identical across all groups (interaction term of group × visit)

BMI, body mass index; HD, habitual diet group; ND, Nordic diet group; REE, resting energy expenditure; VD, vegetarian diet group

**Supplementary Table S5**: Time course of postprandial parameters before and after intervention[a]

| | | | | | | | | | | | | | | | p values[b] | | | | | | |
|---|---|---|---|---|---|---|---|---|---|---|---|---|---|---|---|---|---|---|---|---|---|
| **Plasma GLP-1, pmol/l** | | | | | | | | | | | | | | | | | | | | | |
| | before | | | | | | | after | | | | | | | | | | | | | |
| Time | 0 min | 15 min | 30 min | 45 min | 60 min | 120 min | 180 min | 0 min | 15 min | 30 min | 45 min | 60 min | 120 min | 180min | visit | diet | time | visit*diet | visit*time | diet*time | visit*diet*time |
| HD | 4.16±0.36 | 8.96±0.75 | 11.59±0.59 | 11.14±0.64 | 10.31±0.66 | 9.38±0.49 | 6.89±0.42 | 3.83±0.24 | 11.78±0.94 | 12.32±0.76 | 11.88±0.64 | 11.11±0.60 | 9.89±0.61 | 7.18±0.40 | | | | | | | |
| ND | 4.81±0.29 | 10.28±0.98 | 12.68±1.01 | 11.16±0.70 | 10.74±0.67 | 10.62±0.77 | 8.20±0.60 | 4.27±0.29 | 11.18±1.12 | 13.24±1.01 | 12.36±0.91 | 11.29±0.80 | 10.61±0.73 | 8.22±0.51 | 0.002 | 1 | <0.001 | 1 | 1 | 1 | 1 |
| VD | 4.26±0.26 | 10.84±1.22 | 12.80±0.97 | 11.63±0.76 | 10.78±0.63 | 9.74±0.53 | 7.57±0.52 | 4.29±0.28 | 10.50±0.86 | 13.15±0.77 | 11.98±0.53 | 11.70±0.57 | 9.96±0.49 | 8.88±1.17 | | | | | | | |
| **Plasma NEFA, mmol/l** | | | | | | | | | | | | | | | | | | | | | |
| | before | | | | | | | after | | | | | | | | | | | | | |
| HD | 0.52±0.03 | 0.48±0.03 | 0.37±0.02 | 0.25±0.02 | 0.20±0.01 | 0.21±0.01 | | 0.53±0.03 | 0.47±0.02 | 0.37±0.03 | 0.23±0.02 | 0.19±0.01 | 0.20±0.01 | | | | | | | | |
| ND | 0.51±0.03 | 0.48±0.03 | 0.38±0.02 | 0.26±0.01 | 0.20±0.01 | 0.19±0.01 | | 0.49±0.03 | 0.43±0.02 | 0.36±0.02 | 0.26±0.02 | 0.19±0.01 | 0.20±0.02 | | 0.150 | 1 | <0.001 | 1 | 1 | 1 | 1 |
| VD | 0.57±0.03 | 0.48±0.02 | 0.40±0.03 | 0.28±0.02 | 0.21±0.02 | 0.19±0.01 | | 0.50±0.04 | 0.47±0.03 | 0.37±0.03 | 0.26±0.02 | 0.21±0.02 | 0.21±0.02 | | | | | | | | |
| **Serum triglycerides, mmol/l** | | | | | | | | | | | | | | | | | | | | | |
| | before | | | | | | | after | | | | | | | | | | | | | |
| HD | 1.42±0.11 | 1.38±0.11 | 1.41±0.11 | | 1.54±0.11 | 1.71±0.12 | | 1.33±0.12 | 1.28±0.11 | 1.33±0.11 | | 1.49±0.12 | 1.62±0.13 | | | | | | | | |
| ND | 1.55±0.08 | 1.50±0.08 | 1.57±0.09 | | 1.75±0.10 | 1.87±0.11 | | 1.36±0.10 | 1.30±0.10 | 1.37±0.10 | | 1.50±0.10 | 1.62±0.12 | | <0.001 | 1 | <0.001 | <0.001 | 1 | 1 | 1 |
| VD | 1.47±0.10 | 1.43±0.10 | 1.47±0.10 | | 1.63±0.11 | 1.75±0.11 | | 1.56±0.11 | 1.50±0.11 | 1.51±0.11 | | 1.69±0.11 | 1.86±0.12 | | | | | | | | |

[a] Data are shown as mean ± SEM. Before refers to the postprandial parameters at baseline, after represents the postprandial parameters at the end of the 6-weeks trial

[b] Compared using linear mixed effect models, p-values were adjusted for multiple testing by Bonferroni correction

GLP-1, glucagon like peptide 1; HD, habitual diet group; ND, Nordic diet group; NEFA, non-esterified fatty acids; VD, vegetarian diet group

**Supplementary Table S6**: Fasting and postprandial parameters and risk scores before and after intervention[a]

| | Habitual diet | | Nordic diet | | Vegetarian diet | | |
|---|---|---|---|---|---|---|---|
| | before | after | before | after | before | after | p global[b] |
| LDL cholesterol, mmol/l | 3.5 ± 0.2 | 3.4 ± 0.1 | 4.1 ± 0.2 | 3.8 ± 0.2 | 3.9 ± 0.1 | 3.9 ± 0.1 | 0.007** |
| HDL cholesterol, mmol/l | 1.5 ± 0.1 | 1.4 ± 0.1 | 1.5 ± 0.1 | 1.4 ± 0.1 | 1.5 ± 0.1 | 1.4 ± 0.1 | 1 |
| Total cholesterol, mmol/l | 5.1 ± 0.2 | 5.1 ± 0.2 | 5.9 ± 0.2 | 5.5 ± 0.2 | 5.6 ± 0.2 | 5.6 ± 0.2 | 0.001** |
| y-GT, U/l | 28.4 ± 1.8 | 25.3 ± 1.5 | 33.5 ± 3.7 | 25.0 ± 2.0 | 33.5 ± 4.7 | 34.4 ± 5.8 | 0.045* |
| Triglycerides, mmol/l | 1.4 ± 0.1 | 1.4 ± 0.1 | 1.6 ± 0.1 | 1.3 ± 0.1 | 1.6 ± 0.1 | 1.8 ± 0.1 | <0.001*** |
| NEFA, mmol/l | 0.51 ± 0.03 | 0.47 ± 0.03 | 0.57 ± 0.03 | 0.48 ± 0.03 | 0.56 ± 0.03 | 0.49 ± 0.03 | 1 |
| Triglycerides AUC, mmol/l*min | 214.61 ± 15.57 | 196.57 ± 16.41 | 227.07 ± 12.68 | 198.59 ± 13.97 | 224.78 ± 15.58 | 234.96 ± 15.54 | 0.15 |
| NEFA AUC, mmol/l*min | 34.05 ± 1.76 | 31.98 ± 1.38 | 34.72 ± 1.60 | 32.07 ± 1.78 | 34.32 ± 1.90 | 33.70 ± 2.08 | 1 |
| MSSS | -3.34 ± 0.07 | -3.39 ± 0.07 | -3.24 ± 0.06 | -3.40 ± 0.05 | -3.16 ± 0.06 | -3.15 ± 0.07 | 0.001** |
| CMRI | -1.13 ± 0.52 | -1.05 ± 0.57 | 0.18 ± 0.46 | -0.31 ± 0.40 | 1.03 ± 0.55 | 1.50 ± 0.47 | 0.002** |

[a] Data are shown as mean ± SEM; baseline: HD, n=39; ND, n=41; VD, n=40; endline: HD, n=38; ND, n=39; VD, n=35. Before refers to the respective measurement at baseline, after represents the respective measurement at the end of the 6-weeks trial.

[b] Compared using linear mixed effect models, p-value global refers to the null hypothesis that the change in outcome parameters over time (visits) is identical across all groups (interaction term of group × visit). $P_{global}$ values have been adjusted for multiple testing using Bonferroni correction.

AUC, area under the curve; CMRI, cardiometabolic risk index; GGT, γ-glutamyl transferase; HD, Habitual diet group; HDL, high-density lipoprotein; LDL, low-density lipoprotein; MSSS, Metabolic syndrome severity score; ND, Nordic diet group; NEFA, non-esterified fatty acids; VD, vegetarian diet group

**Supplementary Table S7**: Shotgun lipidomics before and after intervention

*See separate file Suppl_TblS7.xlsx*

Data are shown as mean (pmol/2mL) ± SEM; baseline: HD, n=36; ND, n=37; VD, n=33; endline: HD, n=36; ND, n=37; VD, n=35

**(a)** Compared using linear mixed effect models, p-value global refers to the null hypothesis that the change in outcome parameters over time (visits) is identical across all groups (interaction term of group × visit). P-value FDR corrected.

**(b)** Compared using linear models or zero-inflated gaussian models adjusted for baseline values, p-value global refers to the null hypothesis that the change in outcome parameters over time (visits) is identical across all groups, p-values for pairwise group comparisons were FDR corrected.

Car, acylcarnitines; CarOH, hydroxylated acylcarnitines; CE, cholesteryl esters; Cer, ceramides; DAG, diacylglycerols; DiHexCer, dihexosylceramides; FDR, false discovery rate; HD, habitual diet group; HexCer, hexosylceramides; lm, linear model; LPC, lysophosphatidylcholines; LPC-O, ether-linked lysophosphatidylcholines; LPE, lysophosphatidylethanolamines; ND, Nordic diet group; PC, phosphatidylcholines; PC-O, ether-linked phosphatidylcholines; PE, phosphatidylethanolamines; PE-O, ether-linked phosphatidylethanolamine; PG, phosphatidylglycerols; PS, phosphatidylserines; SEM, standard error of the mean; SM, sphingomyelins; TAG, triacylglycerols; VD, vegetarian diet group; zinf, zero-inflated gaussian model.

**Supplementary Table S8**: Partial correlation of changes in lipidomic variables und changes in cardiometabolic risk scores

*See separate file Suppl_TblS8.xlsx*

Car, acylcarnitines; CarOH, hydroxylated acylcarnitines; CE, cholesteryl esters; Cer, ceramides; CMRI, cardiometabolic risk index; DAG, diacylglycerols; DiHexCer, dihexosylceramides; FDR, false discovery rate; FHS, framingham heart study score; HD, habitual diet group; HexCer, hexosylceramides; lm, linear model; LPC, lysophosphatidylcholines; LPC-O, ether-linked lysophosphatidylcholines; LPE, lysophosphatidylethanolamines; MSSS, metabolic syndrome severity score; ND, Nordic diet group; PC, phosphatidylcholines; PC-O, ether-linked phosphatidylcholines; PE, phosphatidylethanolamines; PE-O, ether-linked phosphatidylethanolamine; PG, phosphatidylglycerols; PS, phosphatidylserines; SEM, standard error of the mean; SM, sphingomyelins; TAG, triacylglycerols; VD, vegetarian diet group; zinf, zero-inflated gaussian model.

**Supplementary Table S9**: Gut microbial diversity before and after intervention[a]

| | Habitual diet | | | Nordic diet | | | Vegetarian diet | | | | |
|---|---|---|---|---|---|---|---|---|---|---|---|
| | before | after | Δ | before | after | Δ | before | after | Δ | p-value[b] ND vs. HD | p-value[b] VD vs. HD |
| **16-S sequencing** | | | | | | | | | | | |
| Richness | 141.62 ± 3.47 | 142.89 ± 9.96 | 1.27 ± 3.57 | 146.58 ± 4.08 | 146.29 ± 3.80 | -0.29 ± 1.80 | 145.51 ± 3.31 | 143 ± 3.10 | -1.89 ± 2.17 | 0.92 | 0.86 |
| Shannon diversity | 3.63 ± 0.06 | 3.66 ± 0.04 | 0.03 ± 0.06 | 3.69 ± 0.04 | 3.70 ± 0.04 | 0.007 ± 0.03 | 3.73 ± 0.03 | 3.70 ± 0.03 | -0.03 ± 0.03 | 0.91 | 0.31 |
| **Whole genome sequencing** | | | | | | | | | | p-value global[c] | |
| Richness (bacteria) | 247.40 ± 37.47 | 226.90 ± 35.01 | -20.50 ± 17.14 | 246.67 ± 20.31 | 224.33 ± 18.60 | -22.33 ± 12.81 | 248.58 ± 23.73 | 241.83 ± 16.85 | -6.75 ± 17.41 | 0.989 | |
| Shannon diversity (bacteria) | 3.86 ± 0.22 | 3.89 ± 0.13 | 0.03 ± 0.15 | 4.06 ± 0.10 | 4.05 ± 0.09 | -0.005 ± 0.05 | 4.03 ± 0.09 | 3.97 ± 0.08 | -0.06 ± 0.08 | 0.912 | |
| Richness (virus) | 87.30 ± 8.37 | 87.30 ± 12.96 | 0.00 ± 10.98 | 95.92 ± 8.23 | 80.00 ± 10.71 | -15.92 ± 9.27 | 81.92 ± 10.48 | 85.00 ± 10.03 | 3.08 ± 12.52 | 0.523 | |
| Shannon diversity (virus) | 2.15 ± 0.16 | 2.20 ± 0.22 | 0.05 ± 0.24 | 2.02 ± 0.17 | 2.03 ± 0.22 | 0.004 + 0.11 | 2.15 ± 0.21 | 2.11 ± 0.23 | -0.04 ± 0.22 | 0.916 | |

[a] Data are shown as mean ± SEM; n = 110 for 16-S samples and n=34 for WGS samples. Before refers to the respective measurement at baseline, after represents the respective measurement at the end of the 6-weeks trial.

[b] Values after intervention were compared between the groups using linear regression, adjusting for baseline value, age, sex, BMI and sequencing depth.

[c] Compared using linear mixed effect models adjusting for sequencing depth, p-value global refers to the null hypothesis that the change in outcome parameters over time (visits) is identical across all groups (interaction term of group × visit).

HD, habitual diet; ND, Nordic diet.

**Supplementary Table S10**: Baseline characteristics of the WGS subgroup[a]

| | Total (n = 34) | HD (n = 10) | ND (n = 12) | VD (n = 12) | p-value[b] |
|---|---|---|---|---|---|
| Sex, m/f | 9 / 25 | 3 / 7 | 3 / 9 | 3 / 9 | 0.956[c] |
| Age, y | 58.7 ± 1.2 | 61.4 ± 1.7 | 54.8 ± 1.7 | 60.3 ± 2.3 | 0.046** |
| BMI, kg/m² | 31.7 ± 0.6 | 31.6 ± 1.1 | 31.6 ± 1.2 | 31.9 ± 0.9 | 0.983 |
| Waist circumference, cm | 106.5 ± 1.6 | 107.0 ± 3.6 | 104.7 ± 2.3 | 107.8 ± 2.6 | 0.790 |
| Hip circumference, cm | 111.9 ± 1.7 | 112.1 ± 2.9 | 110.4 ± 3.1 | 113.3 ± 2.9 | 0.703 |
| Waist-to-height-ratio | 0.6 ± 0.0 | 0.6 ± 0.0 | 0.6 ± 0.0 | 0.6 ± 0.0 | 0.386 |
| BP systolic, mmHg | 133.5 ± 3.0 | 129.7 ± 2.7 | 129.1 ± 4.5 | 141.9 ± 6.7 | 0.151 |
| BP diastolic, mmHg | 87.9 ± 1.8 | 83.9 ± 2.8 | 87.0 ± 2.8 | 92.4 ± 3.6 | 0.180 |
| Heart rate, min-1 | 66.2 ± 1.9 | 63.5 ± 3.0 | 67.9 ± 3.3 | 66.7 ± 3.5 | 0.637 |
| REE, kcal/d | 1673.4 ± 50.6 | 1884.5 ± 114.6 | 1629.0 ± 64.9 | 1541.9 ± 58.7 | 0.016** |
| Body fat, % | 43.6 ± 1.4 | 40.9 ± 2.5 | 43.3 ± 2.6 | 46.2 ± 2.3 | 0.334 |
| Creatinine, µmol/l | 67.6 ± 2.4 | 63.2 ± 3.3 | 68.6 ± 4.2 | 70.4 ± 4.5 | 0.474 |
| Urea, mmol/l | 5.0 ± 0.2 | 5.2 ± 0.6 | 4.9 ± 0.4 | 4.9 ± 0.3 | 0.853 |
| Bilirubin, µmol/l | 9.3 ± 0.6 | 8.1 ± 0.7 | 9.3 ± 0.9 | 10.2 ± 1.4 | 0.392 |
| Uric acid, µmol/l | 313.4 ± 10.1 | 305.2 ± 17.0 | 306.0 ± 17.2 | 326.8 ± 18.9 | 0.637 |
| GGT, U/l | 28.4 ± 2.9 | 25.0 ± 4.1 | 29.8 ± 5.4 | 29.9 ± 5.4 | 0.760 |
| ALT, U/l | 28.8 ± 2.2 | 26.8 ± 3.7 | 29.8 ± 2.9 | 29.4 ± 4.9 | 0.857 |
| AST, U/l | 28.3 ± 1.7 | 27.2 ± 2.5 | 31.4 ± 4.0 | 26.2 ± 1.7 | 0.402 |
| Serum triglycerides, mmol/l | 1.6 ± 0.1 | 1.7 ± 0.2 | 1.8 ± 0.2 | 1.4 ± 0.1 | 0.252 |
| Serum total cholesterol, mmol/l | 5.8 ± 0.2 | 5.6 ± 0.3 | 6.1 ± 0.3 | 5.7 ± 0.3 | 0.385 |
| Serum HDL cholesterol, mmol/l | 1.5 ± 0.1 | 1.7 ± 0.1 | 1.5 ± 0.1 | 1.5 ± 0.1 | 0.506 |
| Serum LDL cholesterol, mmol/l | 4.0 ± 0.2 | 3.8 ± 0.4 | 4.3 ± 0.3 | 4.0 ± 0.3 | 0.542 |
| Plasma glucose, mmol/l | 5.1 ± 0.1 | 5.0 ± 0.1 | 5.1 ± 0.1 | 5,2 ± 0.2 | 0.734 |
| Serum insulin, pmol/l | 94.1 ± 10.7 | 98.9 ± 29.2 | 92.1 ± 11.7 | 92.0 ± 15.9 | 0.961 |
| HOMA | 3.0 ± 0.4 | 3.1 ± 0.9 | 2.9 ± 0.4 | 3.0 ± 0.6 | 0.980 |
| HbA1c, % | 5.4 ± 0.1 | 5.5 ± 0.1 | 5.2 ± 0.1 | 5.5 ± 0.1 | 0.105 |
| Serum hsCRP, mg/l | 2.6 ± 0.4 | 3.4 ± 1.0 | 2.4 ± 0.5 | 2.0 ± 0.3 | 0.287 |
| Plasma GLP-1, pmol/l | 4.0 ± 0.3 | 3.7 ± 0.6 | 4.0 ± 0.5 | 4.2 ± 0.5 | 0.778 |

[a] Data are shown as mean ± SEM; n = 34

[b] Compared using one factor ANOVA

[c] Compared using Pearson chi square test

ALT, alanine aminotransferase; AST, aspartate aminotransferase; BMI, body mass index; BP, blood pressure; f, female; GGT, γ-glutamyl transferase; GLP-1; glucagon-like peptide 1; HD, habitual diet group; HDL, high-density lipoprotein; HOMA, homeostasis model assessment; hsCRP, high-sensitivity C-reactive protein; LDL, low-density lipoprotein; m, male; ND, Nordic diet group; REE, resting energy expenditure; VD, vegetarian diet group

**Supplementary Table S11**: Anthropometric and metabolic characteristics before and after intervention of the WGS subsample[a]

| | **Habitual diet** (n = 10) | | | **Nordic diet** (n = 12) | | | **Vegetarian diet** (n = 12) | | | **p-value global**[b] |
|---|---|---|---|---|---|---|---|---|---|---|
| | before | after | Δ | before | after | Δ | before | after | Δ | |
| Body weight, kg | 96.9 ± 4.9 | 96.8 ± 5.1 | -0.12 ± 0.34 | 89.3 ± 3.5 | 88.0 ± 3.5 | -1.23 ± 0.35 | 89.2 ± 3.0 | 88.7 ± 3.1 | -0.48 ± 0.48 | 1 |
| BMI, kg/m$^2$ | 31.6 ± 1.1 | 31.6 ± 1.1 | -0.06 ± 0.11 | 31.6 ± 1.2 | 31.2 ± 1.2 | -0.44 ± 0.12 | 31.9 ± 0.9 | 31.7 ± 1.0 | -0.19 ± 0.18 | 1 |
| Waist circumference, cm | 107.0 ± 3.6 | 107.1 ± 3.5 | 0.04 ± 1.04 | 104.7 ± 2.3 | 103.4 ± 2.7 | -1.25 ± 0.93 | 107.8 ± 2.6 | 106.5 ± 2.9 | -1.31 + 0.86 | 1 |
| REE, kcal/d | 1884.5 ± 114.7 | 1799.7 ± 101.7 | -48.89 ± 36.24 | 1629.0 ± 64.9 | 1574.8 ± 69.6 | -54.25 ± 26.62 | 1541.9 ± 58.7 | 1515.3 ± 52.8 | 8.64 ± 34.97 | 1 |
| Body fat, % | 40.9 ± 2.5 | 41.1 ± 2.1 | 0.23 ± 0.57 | 43.3 ± 2.6 | 42.4 ± 2.8 | -0.91 ± 0.48 | 46.2 ± 2.3 | 45.4 ± 2.4 | -0.73 ± 0.39 | 1 |
| GGT, U/l | 25.0 ± 4.1 | 21.7 ± 3.4 | -3.30 ± 1.25 | 29.8 ± 5.4 | 26.6 ± 4.9 | -3.17 ± 3.20 | 29.9 ± 5.4 | 27.2 ± 5.3 | -2.75 ± 0.63 | 1 |
| ALT, U/l | 26.8 ± 3.7 | 20.6 ± 2.8 | -6.2 ± 1.70 | 29.8 ± 2.9 | 27.4 ± 4.2 | -2.33 ± 3.34 | 29.4 ± 4.9 | 29.3 ± 6.8 | -0.17 ± 2.38 | 1 |
| AST, U/l | 27.2 ± 2.5 | 22.1 ± 2.0 | -5.10 ± 1.40 | 31.4 ± 4.0 | 29.4 ± 3.9 | -2.0 ± 1.42 | 26.2 ± 1.7 | 28.3 ± 5.4 | 2.08 ± 2.97 | 0.868 |
| Serum total-C, mmol/l | 5.6 ± 0.3 | 5.2 ± 0.3 | -0.40 ± 0.17 | 6.1 ± 0.3 | 5.8 ± 0.3 | -0.39 ± 0.16 | 5.7 ± 0.3 | 5.4 ± 0.3 | -0.28 ± 0.10 | 1 |
| Serum HDL-C, mmol/l | 1.7 ± 0.1 | 1.5 ± 0.1 | -0.15 ± 0.07 | 1.5 ± 0.1 | 1.4 ± 0.1 | -0.08 ± 0.05 | 1.5 ± 0.1 | 1.4 ± 0.1 | -0.08 ± 0.06 | 1 |
| Serum LDL-C, mmol/l | 3.8 ± 0.4 | 3.4 ± 0.2 | -0.44 ± 0.19 | 4.3 ± 0.3 | 4.0 ± 0.4 | -0.30 ± 0.17 | 4.0 ± 0.3 | 3.7 ± 0.3 | -0.25 ± 0.11 | 1 |
| Serum TG, mmol/l | 1.7 ± 0.2 | 1.6 ± 0.3 | -0.09 ± 0.14 | 1.8 ± 0.2 | 1.5 ± 0.2 | -0.35 ± 0.12 | 1.4 ± 0.1 | 1.4 ± 0.1 | -0.02 ± 0.1 | 1 |
| Serum TG AUC, mmol/l*min | 242.0 ± 34.5 | 227.0 ± 35.3 | -15.02 ± 18.91 | 277.6 ± 25.2 | 218.1 ± 24.1 | -59.5 ± 16.8 | 210.7 ± 12.2 | 211.5 ± 16.7 | 0.76 ± 8.43 | 0.15 |
| NEFA fasting, mmol/l | 0.48 ± 0.03 | 0.51 ± 0.04 | 0.03 ± 0.02 | 0.48 ± 0.06 | 0.47 ± 0.05 | -0.01 ± 0.08 | 0.60 ± 0.05 | 0.51 ± 0.06 | -0.09 ± 0.08 | 1 |
| NEFA AUC, mmol/l*min | 30.8 ± 2.4 | 31.1 ± 2.2 | 1.13 ± 1.74 | 30.5 ± 2.4 | 29.8 ± 2.0 | -0.69 ± 2.53 | 38.2 ± 4.2 | 35.9 ± 3.8 | -2.26 ± 4.03 | 1 |

[a] Data are shown as mean ± SEM. Before refers to the respective measurement at baseline, after represents the respective measurement at the end of the 6-weeks trial

[b] Compared using linear mixed effect models, p-value global refers to the null hypothesis that the change in outcome parameters over time (visits) is identical across all groups (interaction term of group × visit). P-values are adjusted for multiple testing using Bonferroni correction

ALT, alanine aminotransferase; AST, aspartate aminotransferase; AUC, area under the curve; BMI, body mass index; GGT, γ-glutamyl transferase; HD, habitual diet group; HDL-C, high-density lipoprotein cholesterol; LDL-C, low-density lipoprotein cholesterol; ND, Nordic diet group; NEFA, non-esterified fatty acids; REE, resting energy expenditure; Total-C, total cholesterol; TG, triglycerides; VD, vegetarian diet group

**Supplementary Table S12**: Fecal SCFA concentrations before and after dietary intervention in participants under metabolic risk[a]

| | Habitual diet | | Nordic diet | | Vegetarian diet | | p-value global[b] |
|---|---|---|---|---|---|---|---|
| | before | after | before | after | before | after | |
| Benzenoic acid, µmol/g | 0.036 ± 0.016 | 0.036 ± 0.017 | 0.025 ± 0.015 | 0.046 ± 0.022 | 0.141 ± 0.118 | 0.234 ± 0.213 | 0.469 |
| Lactic acid, µmol/g | 4.354 ± 1.187 | 3.409 ± 1.080 | 2.159 ± 0.719 | 4.587 ± 1.477 | 12.678 ± 5.827 | 4.386 ± 1.534 | 0.078 |
| Acetic acid, µmol/g | 35.310 ± 3.833 | 41.972 ± 4.694 | 35.697 ± 3.552 | 40.549 ± 4.066 | 36.625 ± 4.707 | 36.499 ± 4.653 | 0.684 |
| Butyric acid, µmol/g | 91.081 ± 4.634 | 92.238 ± 6.356 | 80.894 ± 3.767 | 87.177 ± 4.436 | 85.132 ± 5.108 | 78.689 ± 3.164 | 0.329 |

[a] Data are shown as mean ± SEM; baseline: HD, n = 37; ND, n = 38; VD, n = 39; endline: HD, n = 36; ND, n = 36; VD, n = 34. Before refers to the respective measurement at baseline, after represents the respective measurement at the end of the six weeks trial

[b] Compared using linear mixed effect models, p-value global refers to the null hypothesis that the change in outcome parameters over time (visits) is identical across all groups (interaction term of group × visit)

HD, habitual diet group; ND, Nordic diet group; VD, vegetarian diet group; SCFA, short-chain fatty acid

**Supplementary Table S13:** Characteristics and results of the MixOmics models

| | Model performance characteristics | | | Microbiome | | SCFA | | PGS | | Metabolic Parameters | | Lipidomics | |
|---|---|---|---|---|---|---|---|---|---|---|---|---|---|
| | BER | AUC | p-value | Features (number) | Top 2 contributors | Features (number) | Top 2 contributors | Features (number) | Top 2 contributors | Features (number) | Top 2 contributors | Features (number) | Top 2 contributors |
| **Model 1** | 0.115 | 0.796 | 0.028 | 27 | *Dorea formicigenerans*<br>*Sutterella wadsworthensis* | 2 | Δ pyruvic acid<br>Δ lactic acid | 3 | LDL-C<br>HDL-C | 9 | Δ% TG (fasting)<br>Δ% total-C | 22 | Δ TAG (48:1)<br>Δ TAG (56:7) |
| **Model 2** | 0.225 | 0.849 | 0.019 | 2 | Δ LPSSYN-PWY: superpathway of lipopolysacchari de biosynthesis<br>Δ GLUDEG-I-PWY: GABA shunt | 2 | Δ lactic acid<br>Δ butyric acid | 4 | TG<br>LDL-C | 6 | Δ% TG (AUC)<br>Δ% LDL-C | 37 | Δ TAG (58:10)<br>Δ PC (30:1) |
| **Model 3** | 0.225 | 0.854 | 0.019 | 7 | *Streptococcus phage 9874*<br>*Prevotella_sp_885* | 4 | Δ lactic acid<br>Δ acetic acid | 3 | TG<br>LDL-C | 3 | Δ% TG (AUC)<br>Δ% NEFA (fasting) | 17 | Δ TAG (58:9)<br>Δ PE (38:3) |

Shown are the multi-omic signatures derived from the MixOmics analysis associated with the ND and VD group, including microbiome features, SCFA, PGS, metabolic parameters and lipidomics. Presented is the number of selected features and the top 2 contributors per category

AUC, Area under the curve; BER, balanced error rate; HDL-C, High-density lipoprotein cholesterol; LDL-C, Low-density lipoprotein cholesterol; NEFA, Non-esterified fatty acids; PC, Phosphatidylcholines; PE, phosphatidylethanolamines; PGS, Polygenetic risk score; SCFA, short chain fatty acids; TG, Log-transformed triglycerides; total-C, Total cholesterol; TAG, Triacylglycerols; Δ, change; Δ% relative change.

.

**Supplementary Table S14**: Blood lipid concentrations in diet and PGS groups before and after the end of the trial[a]

| | **Habitual diet** | | | | | |
|---|---|---|---|---|---|---|
| | low PGS | | moderate PGS | | high PGS | |
| | before | after | before | after | before | after |
| Triglycerides (mmol/l) | 1,4 ± 0,3 | 1,6 ± 0,3 | 1,3 ± 0,1 | 1,2 ± 0,1 | 1,5 ± 0,2 | 1,7 ± 0,2 |
| Total-C (mmol/l) | 4,6 ± 0,2 | 4,7 ± 0,2 | 5,2 ± 0,3 | 5,0 ± 0,2 | 5,8 ± 0,3 | 5,9 ± 0,5 |
| HDL-C (mmol/l) | 1,4 ± 0,1 | 1,4 ± 0,1 | 1,5 ± 0,1 | 1,4 ± 0,1 | 1,6 ± 0,1 | 1,5 ± 0,1 |
| LDL-C (mmol/l) | 2,9 ± 0,2 | 3,0 ± 0,2 | 3,6 ± 0,2 | 3,4 ± 0,2 | 4,0 ± 0,2 | 4,1 ± 0,4 |

| | **Nordic diet** | | | | | |
|---|---|---|---|---|---|---|
| | low PGS | | moderate PGS | | high PGS | |
| | before | after | before | after | before | after |
| Triglycerides (mmol/l) | 1,6 ± 0,2 | 1,1 ± 0,1 | 1,7 ± 0,2 | 1,3 ± 0,1 | 1,8 ± 0,2 | 1,6 ± 0,2 |
| Total-C (mmol/l) | 5,2 ± 0,3 | 4,8 ± 0,4 | 6,0 ± 0,2 | 5,7 ± 0,3 | 6,2 ± 0,4 | 5,4 ± 0,4 |
| HDL-C (mmol/l) | 1,3 ± 0,1 | 1,3 ± 0,1 | 1,5 ± 0,1 | 1,5 ± 0,1 | 1,4 ± 0,1 | 1,2 ± 0,1 |
| LDL-C (mmol/l) | 3,6 ± 0,3 | 3,4 ± 0,3 | 4,3 ± 0,3 | 4,0 ± 0,3 | 4,5 ± 0,3 | 3,8 ± 0,3 |

| | **Vegetarian diet** | | | | | |
|---|---|---|---|---|---|---|
| | low PGS | | moderate PGS | | high PGS | |
| | before | after | before | after | before | after |
| Triglycerides (mmol/l) | 1,3 ± 0,1 | 1,4 ± 0,1 | 1,6 ± 0,1 | 1,9 ± 0,2 | 2,1 ± 0,3 | 1,9 ± 0,5 |
| Total-C (mmol/l) | 5,2 ± 0,2 | 5,7 ± 0,3 | 5,5 ± 0,2 | 5,5 ± 0,2 | 6,7 ± 0,7 | 6,3 ± 0,8 |
| HDL-C (mmol/l) | 1,5 ± 0,1 | 1,6 ± 0,2 | 1,4 ± 0,1 | 1,4 ± 0,1 | 1,5 ± 0,3 | 1,7 ± 0,3 |
| LDL-C (mmol/l) | 3,6 ± 0,2 | 3,9 ± 0,2 | 3,8 ± 0,2 | 3,8 ± 0,2 | 4,9 ± 0,8 | 4,6 ± 0,7 |

[a] Data are shown as mean ± SEM. Before refers to the respective measurement at baseline, after represents the respective measurement at the end of the 6-weeks trial

Parameters of lipid metabolism before and after the intervention based on low, moderate and high genetic risk (LDL PGS)

HD, habitual diet group; HDL, high-density lipoprotein; LDL, low-density lipoprotein; Total-C, total cholesterol; ND, Nordic diet group; PGS, polygenetic risk score; VD, vegetarian diet group
